# Persistent Health and Cognitive Impairments Up to Four Years Post-COVID-19 in Young Students: The Impact of Virus Variants and Vaccination Timing

**DOI:** 10.1101/2024.11.06.24316832

**Authors:** Ashkan Latifi, Jaroslav Flegr

## Abstract

The long-term consequences of COVID-19 infection are becoming increasingly evident in recent studies. This repeated cross-sectional study aimed to explore the long-term health and cognitive effects of COVID-19, focusing on how virus variants, vaccination, illness severity, and time since infection impact post-COVID-19 outcomes. We examined three cohorts of university students (N=584) and used non-parametric methods to assess correlations of various health and cognitive variables with SARS-CoV-2 infection, COVID-19 severity, vaccination status, time since infection, time since vaccination, and virus variants. Our results show that some health and cognitive impairments persist, with some even progressively worsening especially fatigue in women and memory in men, up to four years post-infection, with the Wuhan variant having the most significant long-term impact and the Omicron variant the least. Interestingly, the severity of the acute illness was not correlated with the variant of SARS-CoV-2. The analysis also revealed that individuals who contracted COVID-19 after vaccination had better health and cognitive outcomes compared to those infected before vaccination. The study also discusses some limitations inherent in cross-sectional studies, particularly those arising from the stronger tendency of individuals with poorer health, compared to healthier individuals, to avoid infection and prioritize vaccination. To mitigate potential bias, the study proposes focusing on factors such as illness severity and time since infection or vaccination when analyzing persistent symptoms.

## 1. Introduction

The start of COVID-19, associated with a new coronavirus (SARS-CoV-2) found in Wuhan, China, occurred in late 2019 resulting in a worldwide pandemic [1,2]. While the acute effects of the virus have been extensively studied in earlier phases of the pandemic, increasing evidence now highlights the potential for long-term impacts even after recovery. These impacts, commonly known as “long COVID” or post-acute sequelae of SARS-CoV-2 infection can manifest as a range of physical, neurological, and psychological symptoms [3,4]. Research has indicated that certain individuals may suffer a decrease in lung function or develop pulmonary fibrosis following recovery from COVID-19 [5]. In this respect, a study, investigating the post-recovery results of 587,330 patients admitted to hospitals in the United States, with 257,075 having COVID-19 and 330,255 without the virus, found that among the patients, there were 10,979 cases of heart failure that within a 367-day period after discharge. COVID-19 hospitalization led to a 45% increased risk of developing heart failure, particularly in younger patients, white individuals, and those with a prior history of heart conditions [6].

In a community-based cross-sectional study in China, researchers presented results for 1000 individuals who survived COVID-19 20 months after being diagnosed. The most commonly reported symptoms were generalized (fatigue, muscle weakness, joint pain, hair loss, sweating, myalgia, skin rash, and chill) (60.7%), mental (48.3%), cardiopulmonary (39.8%), neurological (37.1%, with 15.6% of it representing cognitive impairment), and digestive (19.1%) symptoms [7]. A meta-analysis of 29 peer-reviewed studies and 4 preprints, encompassing a sample of 15,244 hospitalized and 9,011 non-hospitalized patients, in this regard, revealed that 63.2%, 71.9%, and 45.9% of the sample exhibited at least one post-COVID-19 symptom 30, 60, or 90 days after the onset of illness or hospitalization. The most prevalent symptoms were fatigue and dyspnea. Additional reported post-COVID-19 symptoms included cough (20–25%), anosmia (10–20%), ageusia (15–20%), and joint pain (15–20%). Time trend analysis showed a decrease in symptom prevalence measured at day 30, followed by an increase measured at day 60 [8].

Post-COVID symptoms also encompass cognitive impairment and mental health deterioration. For instance, a large online survey of 4,445 participants investigating the impact of SARS-CoV-2 infection, COVID-19 severity, and vaccination on health and cognition found that both the infection and the severity of COVID-19 had negative effects on patients’ health. Additionally, a correlation was observed between COVID-19 severity and various cognitive functions.

Specifically, the severity of COVID-19 negatively impacted health and fatigue at the time of the study and had a significant adverse effect on intelligence, and negative trends with memory, precision, and performance on the Stroop test [9]. A large-scale study of 81,337 participants in Britian reported that individuals recovered from COVID-19, regardless of symptom presence, showed significant cognitive deficits, in domains such as reasoning, problem solving, spatial planning and target detection, compared to controls after adjusting for various demographic and health factors. Hospitalized patients (N = 192) exhibited substantial deficits, as did non-hospitalized individuals with confirmed infections (N = 326). Analysis indicated that these cognitive differences were not pre-existing. The authors of this study suggested COVID-19 affects multiple cognitive domains, aligning with reports of persistent ‘Long Covid’ symptoms [10]. Another study, recruiting 475 participants (mean age 58.26) who were followed 2–3 years post-hospitalization, indicated that cognitive scores were significantly lower than expected across all domains assessed by the Montreal Cognitive Assessment—memory, executive functioning, attention, language, visuospatial skills, and orientation, with most participants reporting mild to severe depression, anxiety, fatigue, or cognitive decline. Symptoms worsened over time, and severity at 6 months strongly predicted outcomes at 2–3 years. Occupational changes were reported by 26.9% of participants, largely due to health issues, and were closely linked to cognitive deficits [11].

The effects of COVID-19 on young individuals after recovery can include physical, cognitive, and psychological symptoms. Research indicates that some young subjects experience lingering symptoms which can include fatigue, cognitive difficulties, and respiratory issues even months after recovery. A study of individuals from the Swedish birth cohort BAMSE (average age: 26.5 years) investigated post-COVID-19 symptoms, defined as those persisting for at least 2 months following confirmed infection. Altered smell and taste were the most frequent post-COVID-19 symptoms, followed by dyspnea and fatigue [12]. Another recent study assessing COVID-19’s effects on the physical and mental health, fatigue, and cognitive skills of a sample of 214 students, averaging 21.8 years old showed that contracting COVID-19 was associated with a higher frequency of fatigue reports [13]. In addition, the disease’s impact on physical health, mental well-being, fatigue, and reaction time was moderated by severity. In the first 24 months following the infection, improvements in both physical and mental well-being and a reduction in the rates of error were observed. However, there was a tendency towards decline in reaction time and fatigue. By the second year, physical health and error rates improved, but fatigue and reaction times continued their ongoing decline, and mental health began to deteriorate.

More importantly, a meta-analysis of 55 studies involving 1,139,299 participants provided a broader view of post-COVID-19 conditions. This study found more than two hundred symptoms linked to this condition, with gastrointestinal issues, headaches, cough, and fever being the most common, respectively. From these studies, 21 symptoms reported in 11 studies were identified as suitable for inclusion in the meta-analysis, and their subsequent analysis revealed significantly higher pooled estimates for symptoms such as altered or lost smell or taste, dyspnea, fatigue, and myalgia in children and young people who had confirmed SARS-CoV-2 infections. The meta-analysis concluded that many children and young people experience persistent symptoms after SARS-CoV-2 infection [14]. Nonetheless, it should be noted that some studies suggest that ongoing symptoms and impairments in post-COVID-19 conditions may be influenced by factors beyond the virus itself, particularly psychosocial aspects. [15].

Many earlier studies have focused on a limited timeframe shortly after recovery from COVID-19 or overlooked the specific variant involved (see [16–18]). Extending the duration of observation, however, allows researchers to better track the potential long-term effects of post-COVID-19 infection on health and performance. The current article aimed to address these gaps along with the other objectives listed below.

### 1.1. Aims and the Scope of the Study

The primary aim of this study was to determine whether the trends observed in a previous, smaller-scale study (which included a sample size three times smaller and covered a shorter duration post-COVID-19 infection, specifically up to 39 months) continued into the fourth year after the initial illness (covering up to 54 months post-infection). The second objective was to rule out the possibility that the observed correlations between certain health and performance-related variables and the time since infection were merely artifacts of the different virus variants, which emerged in distinct waves before the start of the study and had different effects on health. The third aim of this study was to obtain a sufficiently large sample size to allow for the analysis of not only the impact of COVID-19 but also the effects of vaccination against COVID-19 and the duration since vaccination on health and performance-related variables. A large sample size was essential to ensure adequate representation of the unvaccinated minority, comprising only about 5% of the student population, allowing for meaningful comparisons between vaccinated and unvaccinated groups.

To achieve these objectives in our cross-sectional study, we conducted tests on nearly 300 participants enrolled in an undergraduate evolutionary biology course each year for three consecutive years, consistently during the same month (January). The same panel of performance tests was administered, and data on current health status, details about their COVID-19 illness, and vaccination history were collected through a consistent electronic questionnaire. The collected data were analyzed using multivariate nonparametric methods, and the findings, including those that may seem counterintuitive, were discussed considering the specifics of cross-sectional studies.

## 2. Materials and Methods

### 2.1. Study Participants

Over three consecutive years (2022, 2023, and 2024), all students enrolled in the basic course in evolutionary biology were invited to take part in an anonymous online study. Before beginning the survey, participants were informed about the general goals of the study—examining specific hypotheses in evolutionary psychology and investigating the influence of various factors on cognitive performance. They were assured that their participation was voluntary and that the data would be used solely for scientific purposes, with no means of identifying individual participants. They were also informed that they could withdraw from the study at any time simply by closing the survey page.

Both the examination and the subsequent survey were conducted separately on the Qualtrics platform. After completing the exam, students were notified of their scores, specifically the number of correct answers, and were asked to report this number in the anonymous survey.

The project, including the exact content and wording of the electronic questionnaire, was preregistered on the Open Science Framework (OSF) (doi.org/10.17605/OSF.IO/M5FYC). The primary objective of the study was to assess the impact of emotional priming—recalling positive or negative emotions—on students’ performance, mood, and optimism. Additionally, the secondary goal, as mentioned in the preregistration, was to explore the effects of various biological and social factors, including SARS-CoV-2 infection, on health and cognitive performance.

The study adhered to the ethical guidelines relevant to research involving human participants. The project’s protocol, participant recruitment methods, including the scope and generality of information provided to participants before the study, and the method of obtaining informed consent (achieved by clicking a designated button on the screen), received approval from the Institutional Review Board of the Faculty of Science at Charles University (No. 2021/19).

### 2.2 Questionnaires and Tests

In this survey, we assessed participants’ intelligence using the Cattell 16PF test (Variant A, Scale B) [19]. Memory abilities, including free recall and recognition memory, were evaluated using a modified Meili test [20,21]. Psychomotor skills, particularly reaction time and precision, were measured using the Choice Reaction Time Test and the Stroop Test, with both reaction time and accuracy recorded. Detailed descriptions of these tests are available in [13]. The ‘Reading Time’ variable was calculated as the average Z-score of the times taken to read instructions for the included tests. For each individual, the ‘Accuracy Score’ was calculated as the mean of the Z-scores of the number of correct answers in the four tests: Evolutionary Biology, IQ, Choice Reaction Time, and Stroop test. The ‘Reaction Time Score’ was determined as the mean Z-score of Reading Time and reaction times recorded during the Choice Reaction Time test and Stroop test, while the ‘Memory Score’ was calculated as the mean Z-score of the two memory tests.

In the anamnestic section of the questionnaire, participants answered 21 questions regarding their physical health. These questions addressed the frequency of conditions such as allergies, skin disorders, cardiovascular diseases, digestive tract disorders, metabolic disorders, common infectious diseases, orthopedic disorders, neurological disorders, headaches, physical pains, and other chronic physical issues. Participants also provided information on how often they had taken antibiotics in the previous three years, their frequency of visits to a general practitioner, and the number of times they had been hospitalized for more than a week in the past five years.

Responses to these questions were recorded on 6-point ordinal scales ranging from ‘never’ to ‘daily or more frequently.’ Additionally, they reported the number of non-mental health medications currently prescribed by a doctor, with options ranging from 0 to 7, where 7 indicated six or more medications. At the start and end of the questionnaire, participants rated their current and usual physical feelings using 6-point scales. Finally, participants were asked to estimate their life expectancy, with six response options ranging from ‘more than 99 years’ to ‘less than 60 years’. They also provided information on their usual blood pressure, selecting from five options (“very low”, “rather low”, “normal”, “rather high”, “very high”). No participant selected “very high,” so we used these responses to calculate two binary variables: low blood pressure and high blood pressure. For the exact wording of all questions, refer to the questionnaire text attached to the preregistration form (doi.org/10.17605/OSF.IO/M5FYC).

For health-related questions, certain responses were reversed so that higher scores consistently indicated poorer health, allowing for a standardized interpretation of the results. Then, a Physical sickness index score was calculated using the mean Z-scores of all relevant questions [22].

Similarly, a Mental sickness score was derived from questions related to nine variables, including frequencies of depression, anxiety, other mental health issues, the number of prescribed mental health medications, and assessments of the participants’ current and usual mental states. The latter two questions were posed at both the beginning and end of the questionnaire.

Participants were also asked to rate the frequency of fatigue using a 6-point scale ranging from ‘never’ to ‘daily or several times a day,’ resulting in the variable Fatigue. Additionally, the survey collected demographic and medical history data, including participants’ age and official sex as listed on their birth certificates (with men coded as 1 and women as 0). At the end of the questionnaire, participants were queried about their history of SARS-CoV-2 infection. Responses to COVID-19 status were categorized as follows: “not yet”, “no but I was in quarantine”, “yes, I was diagnosed with COVID-19”, “probably yes, but I was not diagnosed with COVID-19”, and “I am waiting for the result of a diagnostic test”. Those who confirmed a COVID-19 diagnosis were asked to rate the severity of their illness on a 6-point scale, with 1 being ‘no symptoms’ and 6 being ‘I was in ICU.’

All participants provided information about their vaccination status, the date of their most severe COVID-19 episode, and the date of their last vaccination. These dates were used to calculate the time since the onset of COVID-19, the time since the last vaccination, and the binary variable COVID-19 after vaccination. Based on the date of infection (see [23], the likely variant of SARS-CoV-2 was estimated for most participants and encoded into four binary dummy variables: Wuhan, Alpha, Delta, and Omicron (for more information on the codes see Supplementary Table S1).

### 2.3. Data Analysis

To mitigate potential complications arising from the irregular distribution of some dependent variables and especially to avoid issues associated with an imbalanced dataset—with a two-to-one ratio of women to men and vaccinated individuals outnumbering unvaccinated ones by more than tenfold—we employed non-parametric multivariate analytical method, specifically the partial Kendall correlation controlled for age, sex, and survey year, to examine the relationships between variables related to COVID-19 (such as infection status, severity, time elapsed since infection, vaccination status, time since the last vaccination, and occurrences of COVID-19 post-vaccination) and health and cognitive performance indicators. In analyses specific to sex, only age and survey year were controlled. The analysis was conducted using the Explorer v. 1.0 R script [24], leveraging the ppcor R package [25]. To correct for multiple comparisons, we applied the Benjamini-Hochberg correction for multiple testing with a false discovery rate (FDR) set at 0.10 [26], this correction was done only for the main seven variables (physical health issues, mental health issues, fatigue, intelligence, memory, reactions, accuracy), not for post hoc tests exploring for the sources of observed associations. In addition, due to the issue of normality, we employed the Wilcoxon rank-sum test, which is the non-parametric equivalent of the independent t-test, to compare the frequency, age, and the time elapsed since infection between men and women in the dataset. The dataset for this study is publicly accessible on Figshare 10.6084/m9.figshare.27618876 [27]

Technical Notes: In this article, the term “effect” is used in its statistical sense, referring to the difference between the true population parameter and the null hypothesis value. Causal interpretations are only discussed in the Discussion section. Given the exploratory nature of a large part of this study, we also consider trends that did not reach statistical significance.

## 3. Results

### 3.1. Descriptive statistics

The initial dataset comprised 724 individuals, 520 women (71.8%, average age = 21.54, SD = 2.36) and 204 men (28.2%, average age = 21.50, SD = 2.13). Among the women, 169 (36.18%) reported not having been diagnosed with COVID-19, 210 (45.0%) had a confirmed laboratory diagnosis of COVID-19, 52 (11.1%) believed they had contracted the virus but lacked laboratory confirmation, and 36 (7.7%) reported not having COVID-19 but were quarantined. Among the men, 61 (32.4%) reported not having been diagnosed with COVID-19, 85 (45.2%) had a confirmed laboratory diagnosis, 19 (10.1%) believed they had contracted the virus but lacked laboratory confirmation, and 23 (12.2%) reported not having COVID-19 but were quarantined. No students reported awaiting test results.

From the final dataset, we excluded 52 individuals who stated they might have had COVID-19 but lacked laboratory confirmation, 156 individuals who did not respond to the COVID-related questions, and 4 individuals who provided unreliable data, such as reporting an age over 90 or giving identical responses to most questions. The final sample included 584 participants: 415 women (49.4% COVID-negative, 50.6% COVID-positive) and 169 men (49.7% COVID-negative, 50.3% COVID-positive). There was no significant difference in the incidence of COVID-19 between men and women (Chi^2^_(1)_ = 0.004, p = 0.946). The ages of the women and men were similar (21.58, SD 2.36 vs. 21.51, SD 2.02; W = 33945, p = 0.624). Similarly, there was no significant age difference between uninfected and infected individuals (21.59, SD 1.99 vs. 21.53 SD 2.51; W = 44699, p = 0.200).

A significant majority of the students were vaccinated against COVID-19, with the last vaccination occurring on average 12.38 months prior, and at most 50.8 months prior. Among 409 women who responded to the relevant question, 34 (8.3%) were not vaccinated; among 165 men, only 7 (4.2%) were unvaccinated, however, this difference was not statistically significant (Chi^2^_(1)_ = 2.93, p = 0.086). Of the 100 vaccinated women who had contracted COVID-19, 29 (29.0%) contracted the virus after their last vaccination; among 49 men, this was the case for 17 (34.7%), (Chi^2^_(1)_ = 0.499, p = 0.479).

On average, women contracted COVID-19 24.39 months prior (SD=14.32, maximum 54.58), and men 21.68 months prior (SD 13.97, maximum = 52.61), (W = 0.5368, p = 0.261). Among the 282 individuals diagnosed with COVID-19, 26 (9.22%) reported “No symptoms,” 94 (33.33%) described it as “Like mild flu,” another 106 (37.58%) as “Like normal flu,” and 52 (18.44%) as “Like severe flu”; 4 (1.41%) were hospitalized. None reported being treated in the Intensive Care Unit (ICU). The distribution of responses regarding the severity of COVID-19 did not differ between men and women (Chi^2^_(4)_ = 1.47, p = 0.831); for a detailed description of the data, see Table 1 and Supplementary Table S1.

**Table 1.**
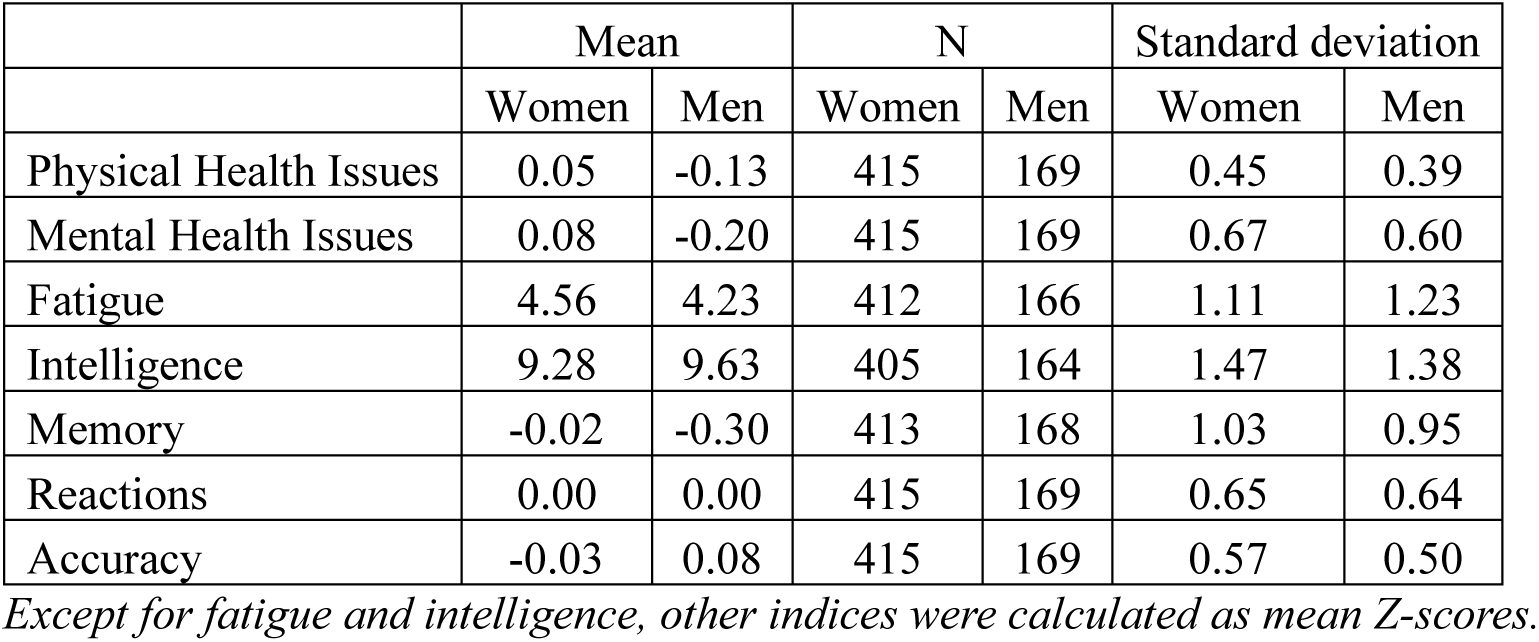
Descriptive statistics of health and cognitive performance-related indices by sex.

|  | Mean |  | N |  | Standard deviation |  |
| --- | --- | --- | --- | --- | --- | --- |
|  | Women | Men | Women | Men | Women | Men |
| Physical Health Issues | 0.05 | -0.13 | 415 | 169 | 0.45 | 0.39 |
| Mental Health Issues | 0.08 | -0.20 | 415 | 169 | 0.67 | 0.60 |
| Fatigue | 4.56 | 4.23 | 412 | 166 | 1.11 | 1.23 |
| Intelligence | 9.28 | 9.63 | 405 | 164 | 1.47 | 1.38 |
| Memory | -0.02 | -0.30 | 413 | 168 | 1.03 | 0.95 |
| Reactions | 0.00 | 0.00 | 415 | 169 | 0.65 | 0.64 |
| Accuracy | -0.03 | 0.08 | 415 | 169 | 0.57 | 0.50 |

*Except for fatigue and intelligence, other indices were calculated as mean Z-scores.*

### 3.2. Confirmation statistics – associations of COVID-related variables with health and cognition issues

In the confirmatory part of the study, we aimed to test the following H_0_ hypotheses:

H1: Trends in health and cognitive performance impairments do not persist into the fourth year after infection.

H2: The observed trends over time are artifacts caused by the gradual replacement of more virulent strains with less and less virulent variants of SARS-CoV-2.

H3: There is no difference in health and cognitive performance between individuals vaccinated before or after contracting COVID-19.

H4: Neither health nor cognitive performance of participants changes with the time elapsed since vaccination.

Age and especially sex significantly impacted some health- and performance-related variables. Consequently, we used partial Kendall correlation, controlling for age, sex, and the survey year, to investigate the effects of COVID-19-related variables on health and performance outcomes (Table 2).

**Table 2.**
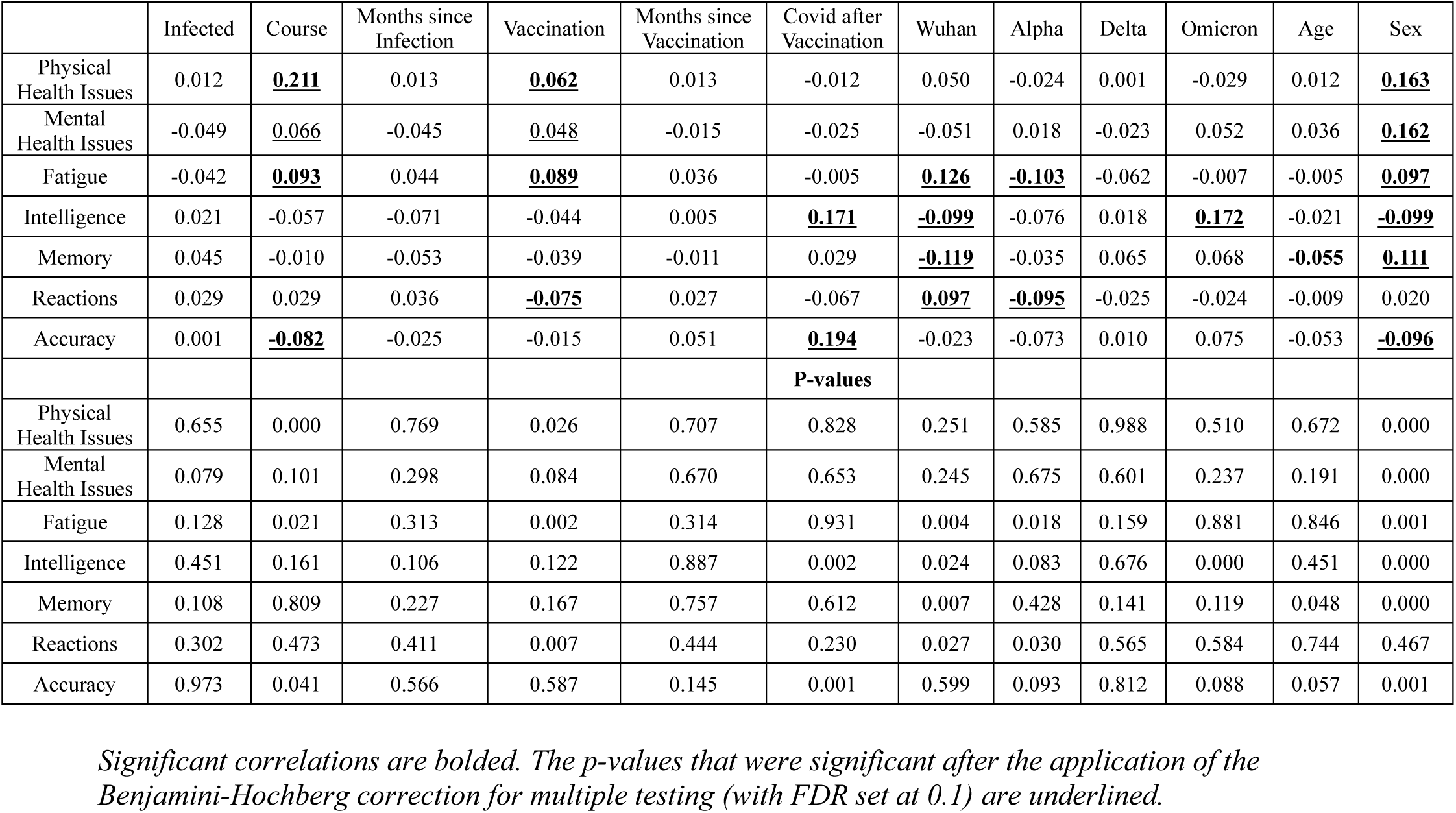
Correlations between health and performance-related variables and COVID-related variables controlled for age, sex, and survey year-All subjects.

|  | Infected | Course | Months since Infection | Vaccination | Months since Vaccination | Covid after Vaccination | Wuhan | Alpha | Delta | Omicron | Age | Sex |
| --- | --- | --- | --- | --- | --- | --- | --- | --- | --- | --- | --- | --- |
| Physical Health Issues | 0.012 | **<u>0.211</u>** | 0.013 | **<u>0.062</u>** | 0.013 | -0.012 | 0.050 | -0.024 | 0.001 | -0.029 | 0.012 | **<u>0.163</u>** |
| Mental Health Issues | -0.049 | <u>0.066</u> | -0.045 | <u>0.048</u> | -0.015 | -0.025 | -0.051 | 0.018 | -0.023 | 0.052 | 0.036 | **<u>0.162</u>** |
| Fatigue | -0.042 | **<u>0.093</u>** | 0.044 | **<u>0.089</u>** | 0.036 | -0.005 | **<u>0.126</u>** | **<u>-0.103</u>** | -0.062 | -0.007 | -0.005 | **<u>0.097</u>** |
| Intelligence | 0.021 | -0.057 | -0.071 | -0.044 | 0.005 | **<u>0.171</u>** | **<u>-0.099</u>** | -0.076 | 0.018 | **<u>0.172</u>** | -0.021 | **<u>-0.099</u>** |
| Memory | 0.045 | -0.010 | -0.053 | -0.039 | -0.011 | 0.029 | **<u>-0.119</u>** | -0.035 | 0.065 | 0.068 | **<u>-0.055</u>** | **<u>0.111</u>** |
| Reactions | 0.029 | 0.029 | 0.036 | **<u>-0.075</u>** | 0.027 | -0.067 | **<u>0.097</u>** | **<u>-0.095</u>** | -0.025 | -0.024 | -0.009 | 0.020 |
| Accuracy | 0.001 | **<u>-0.082</u>** | -0.025 | -0.015 | 0.051 | **<u>0.194</u>** | -0.023 | -0.073 | 0.010 | 0.075 | -0.053 | **<u>-0.096</u>** |
|  |  |  |  |  |  | **P-values** |  |  |  |  |  |  |
| Physical Health Issues | 0.655 | 0.000 | 0.769 | 0.026 | 0.707 | 0.828 | 0.251 | 0.585 | 0.988 | 0.510 | 0.672 | 0.000 |
| Mental Health Issues | 0.079 | 0.101 | 0.298 | 0.084 | 0.670 | 0.653 | 0.245 | 0.675 | 0.601 | 0.237 | 0.191 | 0.000 |
| Fatigue | 0.128 | 0.021 | 0.313 | 0.002 | 0.314 | 0.931 | 0.004 | 0.018 | 0.159 | 0.881 | 0.846 | 0.001 |
| Intelligence | 0.451 | 0.161 | 0.106 | 0.122 | 0.887 | 0.002 | 0.024 | 0.083 | 0.676 | 0.000 | 0.451 | 0.000 |
| Memory | 0.108 | 0.809 | 0.227 | 0.167 | 0.757 | 0.612 | 0.007 | 0.428 | 0.141 | 0.119 | 0.048 | 0.000 |
| Reactions | 0.302 | 0.473 | 0.411 | 0.007 | 0.444 | 0.230 | 0.027 | 0.030 | 0.565 | 0.584 | 0.744 | 0.467 |
| Accuracy | 0.973 | 0.041 | 0.566 | 0.587 | 0.145 | 0.001 | 0.599 | 0.093 | 0.812 | 0.088 | 0.057 | 0.001 |

*Significant correlations are bolded. The p-values that were significant after the application of the Benjamini-Hochberg correction for multiple testing (with FDR set at 0.1) are underlined.*

Our analysis showed that the severity of COVID-19 was significantly associated with worse physical health, fatigue, and a lover cognitive accuracy. After applying the Benjamini-Hochberg (BH) correction for multiple testing, all of these associations remained significant. The COVID-19 vaccine exhibited significant positive associations with physical health issues and fatigue and a negative association with reaction times. These findings indicated a worse physical health and better (shorter) reaction times in vaccinated students. All of these associations remained significant even after the BH correction. Subjects who contracted COVID-19 after receiving the vaccine showed a significantly higher intelligence and accuracy scores than those who contracted COVID-19 before receiving the vaccine. These also remained significant after the BH correction. The analysis of the effects of virus variants revealed that the Wuhan variant had the most adverse effects compared to other variants. This variant correlated with lower scores in the subjects’ intelligence, memory, and higher scores in their reaction times, which mean worse cognitive performance, and resulted in higher levels of fatigue. These associations remained significant even after applying the BH correction. The Alpha variant demonstrated significant negative associations with fatigue and reaction times, indicating lower levels of fatigue and also shorter (better) reaction times compared to other variants. The Omicron variant exhibited a significant positive association with the subjects’ intelligence scores, indicating higher intelligence scores in those infected with this variant. The Delta variant did not have any significantly negative or positive associations with any of these seven variables.

The results indicated that there were no significant associations between the seven index variables and COVID-19 infection, the time elapsed since COVID-19 infection, or the time elapsed since receiving a COVID-19 vaccination (see Table 2). Results of separate analyses for women and men are shown in supplementary Tables S2–S5.

When we included the variable representing the SARS-CoV-2 variant in the model (coded ordinally, with 1 for the earliest variant, Wuhan, and 4 for the most recent variant, Omicron), the results remained unchanged (see Supplementary Tables S6 and S7). This result suggests that the observed patterns of effects, trends, and the absence of trends among the predictors were not simply artifacts caused by the replacement of more virulent variants with progressively less virulent ones throughout the course of the pandemic.

Our analyses of the trajectories of the index variables over the time elapsed since COVID-19 infection (almost 54 months) for all participants, and also separately for men and women, indicated that the first-degree linear models had smaller Bayesian Information Criterion (BIC) coefficients compared to those of higher-degree polynomial models. Accordingly, the following discusses the results based on our first-degree linear models.

The results for all participants showed that a relatively small decline in memory performance over the time elapsed since infection was noticeable. In contrast, trends of marginal improvement in mental health issues, accuracy, and intelligence were observed. Fatigue showed the largest increase over the time elapsed since infection. Marginal trends of deterioration in reaction times and physical health were also present (see Figure 1 and Table 2). Results of separate analyses for women and men are shown in supplementary Figures S1 and S2 and Supplementary Tables S2– S5.

**Figure 1.**
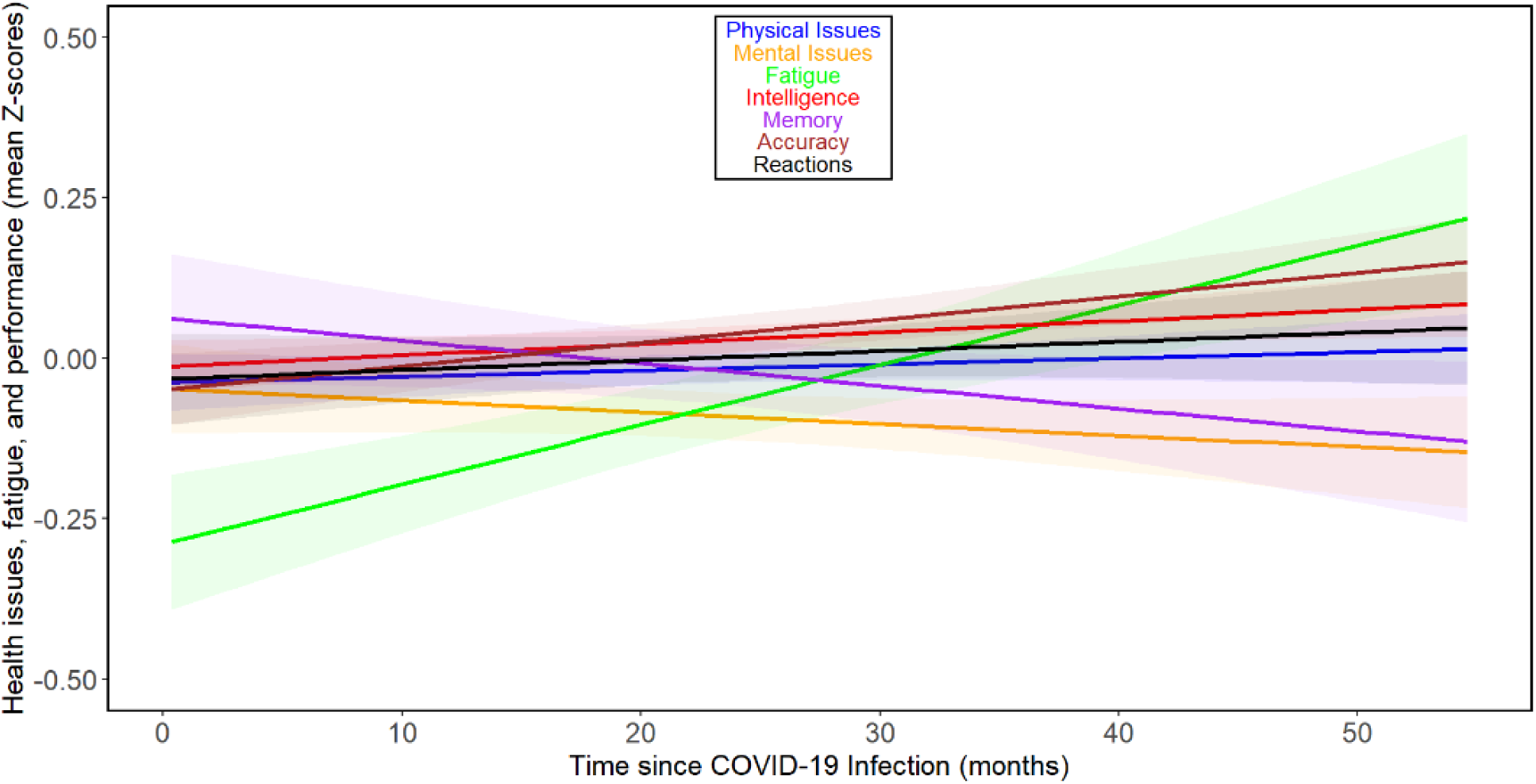
Changes in health and performance-related variables with time since COVID-related variables - All participants *Based on the comparison of BIC values, all observed dependencies were best approximated by a first-degree polynomial (a straight line). The shaded areas around the lines indicate the 60% confidence intervals. It should be noted that the slopes of the lines on the graph cannot be directly compared with the (more accurate) Tau values calculated using the non-parametric partial Kendall test, which is less sensitive to the presence of outliers and additionally controlled for the influence of 3 confounding variables (sex, age, and year of survey)*.

The results of the previous study [13] suggested that the direction and rate of changes over time varied during the first two years after infection but likely increased thereafter. Therefore, we repeated the analyses for 111 subjects from 24 months post-infection onwards. Figure 2 and Supplementary Tables S8–S13 show that there were negative associations between months since infection and physical health, mental health, fatigue, intelligence, memory, reaction times, and accuracy during the fourth year after infection in this group, of which the correlation between intelligence and months since infection was significant. Based on the binomial distribution, the probability of obtaining 7 correlations indicating adverse relationships out of 7 would be 0.008. See Figures S3 and S4 for separate analyses of the trajectories of the seven index variables over the time beyond 24 months post-infection for women and men, respectively. When fifty-one participants who were infected at least 36 months before the study were analyzed, the results were more complex (Supplementary Tables S14 and S15). In the whole population, we saw a relative decrease of intelligence (Tau = -0.072), and worsening of reaction times (Tau = 0.093), however, we also saw clear improvement in accuracy (Tau = 0.125). The analyses split by sex showed that in women all health and cognitive performance related variables except accuracy (Tau = 0.144) and memory (Tau = 0.038) impaired (physical issues: Tau = 0.077, mental issues: Tau = 0.213, fatigue: Tau = 0.083, intelligence: Tau = -0.079, and reactions: Tau = 0.054. In contrast, in men all changes except worsening in reaction times (Tau = 0.350) were improvements– physical issues: Tau = -0.328, mental issues: Tau = -0.414, fatigue: Tau = -0.159, intelligence: Tau = 0.129, memory: Tau = 0.314, and accuracy: Tau = 0.060. It must be noted, however, that none of the observed trends in participants infected more than three years ago were statistically significant, and only 15 participants were included in the subset of men (for the correlational analyses split by sex, see Supplementary Tables S16 and S17 for women, and S18 and S19 for men, respectively).

**Figure 2.**
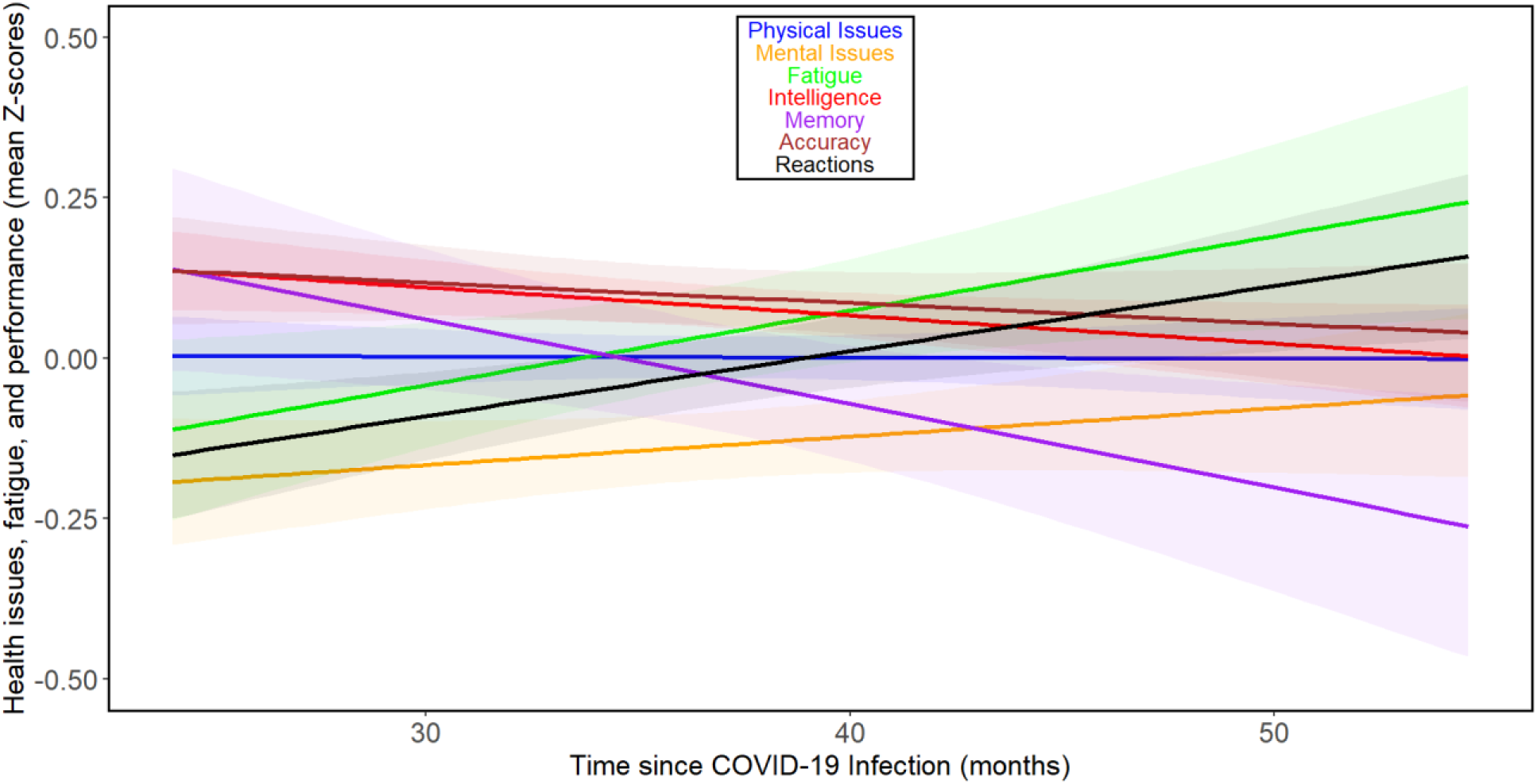
Changes in health and performance-related variables with time since COVID-19 beyond 24 months post-infection-related variables in all participants *For the legend, refer to* Figure 1.

When we analyzed the effect of time since vaccination, as opposed to time since COVID-19 infection, we observed rather distinct results (Figure 3). The comparison of results in columns 3 and 5 of Table 2 shows that correlations with time since vaccination were consistently weaker than those with time since COVID-19. In the case of accuracy and intelligence, the correlations even shifted from negative (indicating worsening performance over time) to positive. None of the correlations came anywhere close to the threshold for significance. See Figures S5 and S6 for separate analyses of the trajectories of the seven index variables over the months following vaccination for women and men, respectively.

**Figure 3.**
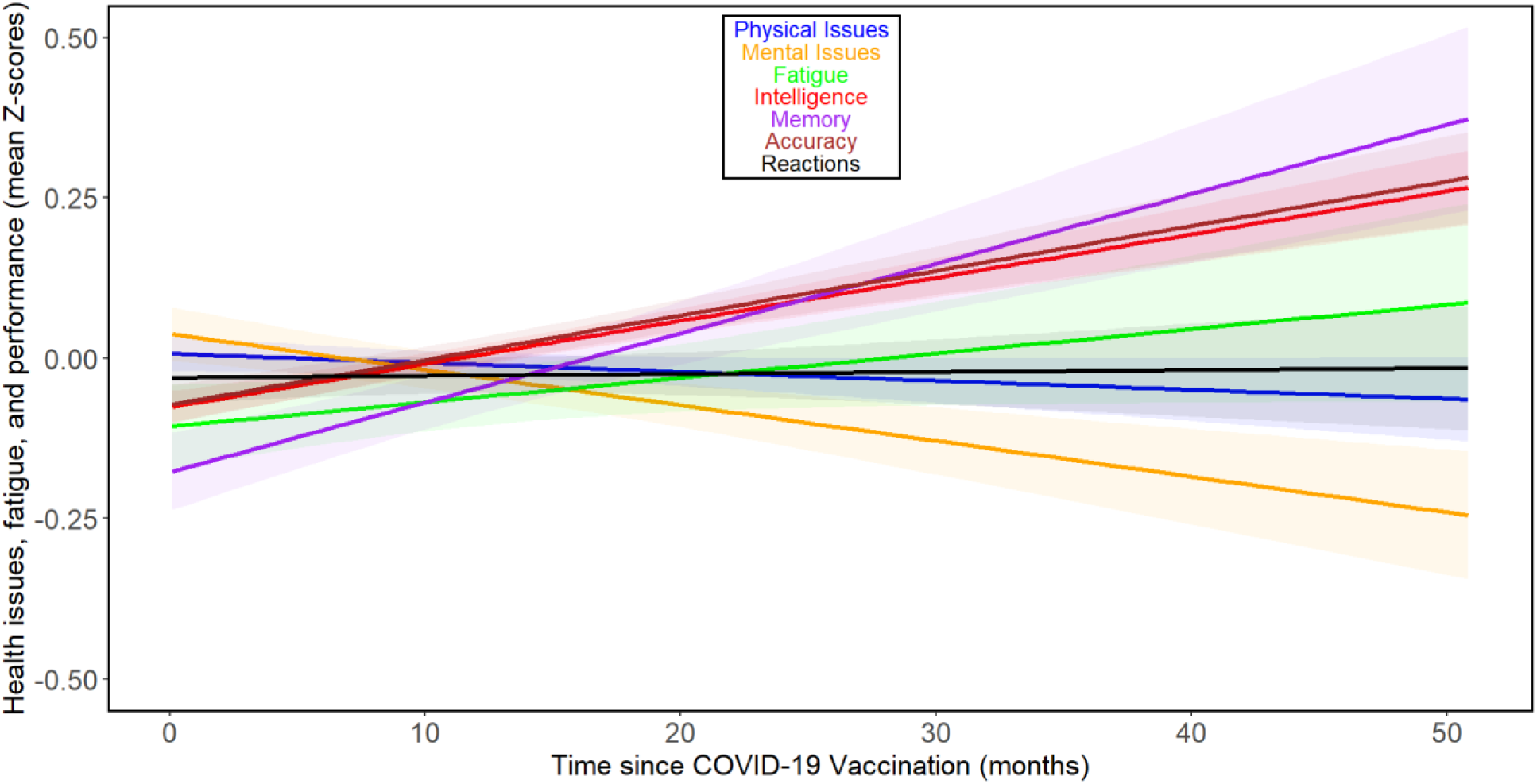
Changes in health and performance-related variables with time since COVID vaccination-related variables in all participants *For the legend, refer to* Figure 1.

To summarize the results of the confirmatory part of the study:

We rejected hypotheses H1 and H3, but we could not reject hypotheses H2 and H4. In other words, our data suggest that:

1. The deterioration of certain health and cognitive functions continues even during the fourth year after COVID-19. However, the direction of the nonsignificant trends in the subset of 15 men more than three years post-COVID suggested that the deteriorations may stop and potentially even reverse except for intelligence and reaction times, at least in men.
2. To test hypothesis H2, we also included the confounding factor ‘Covid variant’ in the model. The results showed that the lack of significant correlations between the outcome variables and the time since infection persisted, even with this factor in the model. However, all nonsignificant trends were markedly weaker when the confounding variable of virus variant was controlled. In several cases, the corresponding trend even reversed direction. For example, the Tau value characterizing memory decline shifted from -0.053 to 0.006, and in the case of intelligence, from -0.071 to 0.012 (see Supplementary Tables S6 and S7, and compare with Supplementary Tables S20 and S21). Thus, based on our data, we cannot refute the H2 hypothesis, which posits that the observed non-significant trends (the intensification of certain symptoms of impaired health or cognition over time since infection) are artifacts caused by the gradual replacement of more virulent strains with progressively less virulent variants of SARS-CoV-2.
3. Additionally, individuals who were vaccinated before contracting COVID-19 are in better health and have better cognitive performance than those who received the vaccination only after recovering from COVID-19.
4. Finally, neither the health nor the cognitive performance of participants changes with the time elapsed since vaccination.

### 3.3. Exploration statistics – association of Covid-related variables with primary health and cognition related variables

In this exploratory part of the study, we did not apply corrections for multiple testing [28].

#### 3.3.1. All participants

Generally, the severity of COVID-19 (course of COVID) showed strong, statistically significant positive associations with worse health outcomes and lower cognitive test performance, see Supplementary Tables S20 and S21. These included lower precision in Stroop test performance, higher levels of allergies, skin conditions, digestive issues, infections, neurological problems, headaches, recurrent health issues, overall physical pain, depression, overall psychological problems, higher consumption of psychotropic medication, higher rates of doctor visits, higher use of other types of medication, greater antibiotic intake over the past three years, more frequent hospitalizations in the past five years, and shorter life expectancies.

In contrast, the associations of health and performance with merely having contracted COVID-19 were much less frequent and generally weaker and non-significant. Exceptions included a significant negative association with usually feeling mentally miserable, and positive associations with recognition memory, neurological disorders, infections, and the frequency of antibiotic use in the past three years. The last two variables included COVID-19 and the treatment of associated bacterial infections, making their correlation with contracting COVID-19 inevitable. Individuals who had contracted COVID-19 reported generally feeling mentally well.

COVID-19 vaccination demonstrated positive associations with allergies, digestive problems, cardiovascular issues, headaches, recurrent health problems, medication intake, and usually feeling mentally miserable. However, it was associated with shorter (better) simple reaction times.

Individuals who contracted COVID-19 after vaccination reported higher scores on the evolutionary biology exam and better Stroop test precision. They also had higher rates of skin conditions, elevated blood pressure, and reported a longer life expectancy compared to those infected before vaccination.

To assess the differences in the effects of various virus variants, we studied the correlations of the output variables with four dummy variables, each indicating whether a person was infected with a specific variant (coded as 1) compared with other variants (coded as 0). Investigating the correlations of the output variables with these dummy variables showed that infection with the Wuhan variant was negatively associated with memory (it had negative associations with both free-recall memory and recognition memory). It was also linked to higher rates of recurrent health problems. Infection with the Alpha variant was associated with worse precision in simple reaction time tests, lower instances of infections, higher rates of orthopedic issues, and a higher tendency to feel mentally miserable. The Delta variant did not show significant associations with the output variables, while the Omicron variant was linked to higher levels of depression and anxiety.

Associations between the studied output variables and the time since infection were weak and sparse, and even weaker were corresponding associations between these variables and the time since vaccination.

Results of all analyses remained nearly the same when we also controlled for the time elapsed since infection in addition to age, sex, and survey year (see Supplementary Tables S22 and S23), showcasing that the associations of virus variants with health and cognition were not confounded by this variable.

#### 3.3.2. Women

Investigating the results for women and men separately revealed several statistically significant associations. The comprehensive set of the results is presented in Supplementary Tables S2 and S4.

In the female group, the long-term effects of COVID-19 infection were evident in higher rates of allergies, higher susceptibility to infections, a higher number of doctor visits, and an elevated rate of antibiotic consumption over the past three years. Conversely, they expressed lower psychological distress, as indicated by a reduced tendency to usually feel mentally miserable, along with higher memory and word recognition scores. More critically, the severity of COVID-19 was associated with a greater incidence of allergies, skin conditions, digestive issues, infections, neurological problems, and headaches. It also correlated with more frequent physical problems, a higher number of doctor visits, and higher rates of antibiotic use during the past three years, as well as greater medication consumption for both mental and physical health concerns. The analysis for this group also showed that there were negative associations between the time elapsed since infection and both the utilization of psychotropic medications and the number of doctor visits.

COVID-19 vaccination was associated with better performance in simple reaction times test, higher rates of allergies, cardiovascular issues, orthopedic problems, headaches, medication consumption, a tendency to feel mentally miserable, and a shorter life expectancy in the female group. Additionally, there was a positive correlation between the time elapsed since COVID-19 vaccination and high precision on the Stroop test. Those who contracted COVID-19 after vaccination had higher scores on the evolutionary biology exam and better precision on the Stroop test. They also reported higher rates of skin conditions, infections, neurological problems, and feeling physically miserable, but reported a longer life expectancy.

We also investigated the associations between the four variants of the SARS-CoV-2 virus and the output variables. The analysis showed that the infection with the Wuhan variant was associated with more frequent orthopedic problems, overall physical pains, and recurrent health issues. The infection with the Alpha variant was associated with lower simple reaction time precision, lower precision on the Stroop test, fewer skin conditions, lower rates of infections, and reduced levels of medication consumption. The infection with the Delta variant was associated with higher rates of metabolic problems, but lower levels of anxiety. And finally, the infection with the Omicron variant was associated with lower scores on the evolutionary biology exam, better performance on the Stroop test, lower orthopedic issues, higher levels of depression, higher levels of anxiety, and higher rates of the utilization of medication for mental issue purposes.

#### 3.3.3. Men

Despite the small sample size of men, COVID-19 was significantly associated with worse reading time and more frequent hospitalization over the past five years, but lower levels of medication use for mental purposes and overall less frequent other psychological problems. More importantly, the severity of COVID-19 was associated with higher rates of allergies, metabolic problems, recurrent health conditions, overall other physical pains, overall other psychological problems, higher utilization of medications for mental health purposes, a larger number of doctor visits, higher overall medication consumption, higher incidents of hospitalization during the past five years, and worse precision on the Stroop test. The time elapsed since infection did not show any significant correlations with the output variables, but the time since vaccination was associated with lower levels of depression and feeling less mentally distressed at the time of the survey. Those who had contracted COVID-19 after vaccination in this group had lower levels of feeling physically miserable now than those who had contracted COVID-19 before vaccination. COVID-19 vaccination was associated with higher rates of infections, headaches, lower incidents of hospitalizations during the past five years, and lower blood pressure.

Investigating the effects of the four variants of the SARS-CoV-2 virus indicated that the infection with Wuhan variant was associated with worse performance in both memory tests, lower orthopedic issues, and lower anxiety. The infection with Alpha was associated with worse performance in the recognition memory task, shorter reading times, higher cardiovascular problems, more orthopedic issues, more headaches, higher rates of doctor visits, and longer life expectancies. The infection with Delta variant was associated with better performance in both memory tests, lower doctor visits, and lower consumption of medications. The infection with Omicron variant was associated with better performance on the evolutionary biology exam, lower levels of feeling mentally miserable now, and a negative association with low blood pressure. The aforementioned results concerning the long-term consequences of COVID-19 infection differ markedly from those related to the acute phase of the illness. Supplementary Tables S6 and S7 show that no effect of any SARS-CoV-2 variant on the reported severity of acute COVID-19 was detected.

## 4. Discussion

The current study employed a design and technical execution similar to our previous research [13]. However, it differed in that the data for this study were collected from participants of a basic-level course in evolutionary biology, which had twice the number of students compared to the advanced course used in the previous study. This larger student population allowed for a more robust sample size. Data collection occurred each time in January instead of May and spanned the years 2022, 2023, and 2024, whereas the previous study was limited to 2022 and 2023.

Additionally, the questionnaire used in this study differed slightly from the previous one, with most changes affecting questions unrelated to the core objectives of either study. The new study aimed to determine whether the health and performance trends observed over 39 months post-infection in the previous study persisted in the following 12 months. Furthermore, the current study sought to determine whether these trends were genuinely due to the increasing time since infection or merely because the time since infection closely correlated with the virus variant a person was infected with. Lastly, the nearly threefold increase in respondents in the new study (584 vs. 214) allowed for the analysis of the impact of vaccination, which only a small number of students had not received (8.3% of women and 4.2% of men).

### 4.1. Trends in health and performance post COVID

The results showed that some trends reversed, with health and performance parameters beginning to improve, although some negative trends persisted even 51 months post-COVID. This was particularly evident in the higher levels of fatigue among both men and women. At least in men, there was a continued decline in physical health. While previous data from a smaller dataset suggested that the direction of ongoing changes varied over time, the new, more extensive data indicated that the trends were likely linear, as the relevant data points were best fitted by a first-degree polynomial, a straight line. Even so, beyond three years post-Covid infection mental health, memory, intelligence, fatigue, reaction times, and accuracy deteriorated further; however, physical health remained relatively constant. When men who had been infected for more than 3 years were analyzed separately, it appeared that the negative trends were finally beginning to reverse. However, given the small number of these men (15), this result should be interpreted with caution, and it will certainly need to be confirmed in a larger sample.

In our study, we interpreted the observed trends of performance decline over time since infection as either a result of the gradual accumulation of the infection’s negative effects or, alternatively, as the replacement of more virulent strains by less virulent ones over the course of the epidemic. However, due to the design of the study, all participants were approximately 21.5 years old. As a result, the time since infection negatively correlated with the age at which each participant contracted COVID. It is therefore possible that the poorer health and cognitive performance in individuals who had COVID longer ago could be related to the fact that they were younger at the time of infection, and their bodies, particularly their brains, were more susceptible to the negative effects of the illness.

### 4.2. Impact of SARS-CoV-2 variants

Our study confirmed that the SARS-CoV-2 variant affected the variables studied, similar to findings by [29,30]. However, these studies focused mainly on physical health variables or case fatality rates, whereas our research examined a broader range of health and cognitive outcomes. Specifically, Torabi et al., almost exclusively focused on physical health variables such as myalgia, cough, taste/smell disorder, diarrhea, etc., investigated personal characteristics, symptoms, and underlying conditions of individuals infected with different variants of SARS-CoV-2 over a period of two years, while Xia et al. concentrated on the case fatality rates of the variants of this virus during the epidemic periods by continents. The observed associations between the studied dependent variables and the dummy variables representing the different virus variants did not change when the time since infection was included as a covariate in the Kendall’s partial correlation tests, save for the removal of the effects of the Wuhan variant on three variables related to intelligence, reactions, and memory. This indicates that the observed differences are primarily driven by the specific virus variant present at the time of infection, rather than the time elapsed since infection. Conversely, the noticeable changes in the model outputs after controlling for the ordinal virus variant variable suggest that the trends—namely, the weakening or intensifying of symptoms over time since infection could be driven by differences in the virulence levels of the virus variants contracted at different times.

Our data do not clarify whether the more pronounced symptoms associated with the Wuhan variant are due to its inherent properties or the fact that, during its spread, there was no vaccine available in the Czech Republic, leading to all patients infected with this variant being unvaccinated. However, the absence of vaccination cannot fully account for the differences, as most students infected with the Alpha variant also had not been vaccinated, given that the vaccination of young, low-risk individuals in the Czech Republic began only in the summer months of 2021, i.e. after the Alpha variant wave had passed. This suggests that factors beyond vaccination status likely contributed to the differences in symptoms between the Wuhan variant and other variants. An unexpected result of the study is that the observed differences between the virus variants were limited to the long-term consequences of COVID-19, and had no measurable effect on the severity of the acute phase of the illness.

When discussing the varying effects of different virus variants, it is important to reiterate that our study was conducted on a young student population, where the illness generally presented mildly, as evidenced by the fact that only four students (0.7%) required hospitalization due to COVID-19. It’s possible, and indeed likely, that the impact of different virus variants on individuals in older age groups, particularly those who experienced severe or very severe illness, would differ significantly from our observations.

### 4.3. Role of vaccination in health and performance

As previously mentioned, individuals who had been vaccinated showed poorer health outcomes in some aspects compared to unvaccinated individuals. As suggested in earlier studies [9,13], the most probable explanation for this phenomenon is that individuals with poorer health were more likely to opt for vaccination as a precaution during the pandemic. This explanation is substantiated by a number of studies showing that the elderly and those at risk held more positive attitudes towards vaccination [31,32]. In our study, we further supported this hypothesis by finding no significant correlations between health and performance-related variables and the time elapsed since vaccination. Even when nonsignificant trends were present, they were generally much weaker than those related to the time since COVID-19 infection. It is also important to note that, in contrast to the frequent adverse correlations with time since COVID-19, nearly all correlations with time since vaccination were beneficial.

Our data corroborate earlier findings that the experience of having had COVID-19 has a relatively weak impact on the health status and cognitive performance of young individuals [9,13,33]. On average, those who had been infected demonstrated slightly better health and performed better on performance tests. The most pronounced and often significant negative symptoms were observed in individuals previously infected with the Wuhan variant, while those infected with the Omicron variant showed the least severe symptoms. However, even in our sample of young individuals, the severity of the illness had a strong impact. The intensity of symptoms associated with worsening health generally change only slowly with time since the COVID-19 illness (mostly showing nonsignificant trends). Vaccinated individuals generally exhibited poorer health compared to unvaccinated individuals. However, the difference in their health and performance did not change significantly with time since vaccination, suggesting a lack of causal relationship. *Most importantly, individuals vaccinated before contracting COVID-19 demonstrated better performance on cognitive tests and often showed significantly or nonsignificantly better health parameters compared to those vaccinated after recovering from COVID-19*.

The relatively weak impact of having had COVID-19 and the observed negative association between vaccination and health status can be attributed to the design of the study. Since this was an observational cross-sectional study, individuals were not randomly assigned to groups such as infected vs. uninfected, vaccinated vs. unvaccinated, or vaccinated before vs. after contracting COVID-19. Previous studies have shown that individuals with better health were more likely to contract COVID-19, as they took fewer precautions against infection, compared to those with chronic health issues [34,35]. Indeed, this inverse relationship between long-term health conditions and the likelihood of contracting the infection was statistically confirmed [9,36]. The study that separately tracked long-term and current health statuses showed a non-significant negative association between prior health issues and contracting COVID-19, and a significant positive association between contracting COVID-19 and current health problems [9]. The same study revealed a non-significant negative association between vaccination and current health issues and a significant positive association with long-term health problems. Unlike the present study, the earlier study included 4,445 individuals from a general internet population. This population was much more age-diverse, allowing for the study of age effects. As expected, age was much more strongly correlated with health and cognitive performance than in our highly age-homogeneous sample. The study showed that younger individuals were more likely to contract COVID-19 and less likely to be vaccinated compared to older individuals, who were generally more at risk. The positive association between long-term poor health and the willingness to get vaccinated persisted even after controlling for age, suggesting that the reported link between vaccination willingness and age is not mediated by age itself, but by age-related health issues.

For a precise examination of the impact of COVID-19 and vaccination on health status and cognitive performance, data from randomized experimental studies would be necessary. However, these studies are not feasible, except in rare cases, due to ethical constraints. A potential solution is to monitor the severity of the illness, the time elapsed since infection, and the time elapsed since vaccination, as we did in this and our previous study [13]. Additionally, prospective observational studies can provide important information regarding the causality of observed associations. However, prospective studies are more demanding to conduct, require large sample sizes, and involve repeated assessments of the same individuals before and after illness or vaccination. A prospective study conducted on 30,000 Czech internet users, of which 5,164 participated repeatedly, showed that having COVID-19 had a strong impact on physical health (partial Tau = 0.25) and a weaker, yet still highly significant impact (partial Tau = 0.04, p < 0.001) on mental health [36]. Data collection for this study was completed by the end of March 2021, so the study could not assess the impact of vaccines on health status.

### 4.4. Strength and limitations

The present study has several strengths and limitations. One of the main strengths, compared to similar studies, is the rigorous implementation of all available measures to limit selection bias and reduce distortions arising from subjective opinions on the topic. More than 80% of course participants volunteered for the study. Given the strong polarization within Czech society regarding the effects of COVID-19 and vaccination on health, steps were taken to avoid potential bias stemming from participants’ personal beliefs. For example, students were informed that the study would investigate the impact of various ‘biological factors’ on health, but they were not explicitly told that COVID-19 infection would be one of these factors. Additionally, relevant questions about COVID-19 and vaccination were placed at the end of the questionnaire, ensuring that participants answered health-related questions and completed performance tests without being given any indication that the study might relate to COVID-19.

However, the study also had some limitations. The first is an inherent limitation of all cross-sectional observational studies: the observed statistical associations do not establish causality and cannot discern what is cause and what is effect. Another limitation was that participants self-reported their health status, which was not verified through medical examinations. A further limitation was technical: the questionnaire asked participants for the date of their last vaccination, which, along with the date of their most severe COVID-19 episode, was used to determine the binary variable “COVID-19 before or after vaccination.” In reality, some participants with a longer time since their last COVID-19 infection than since their last vaccination had their first vaccination before contracting COVID-19. The presence of such misclassified individuals in the dataset likely skewed the results, specifically diminishing the measured effects of the binary variable vaccination before COVID and increasing the risk of Type II errors—failing to detect an existing effect. However, their presence did not increase the risk of Type I errors—detecting a non-existent effect.

## 5. Conclusions

Our study confirmed that contracting COVID-19, especially when accompanied by severe symptoms, can negatively affect health and cognitive performance. It also showed that even among young individuals, who mostly experienced relatively mild cases of COVID-19, some symptoms (such as frequent fatigue, impaired memory, and certain health issues) can persist for up to four years. Notably, while these symptoms tend to worsen for everyone over the first three years, they may continue to worsen in women even during the fourth year after infection. In contrast, neither participants’ health nor cognition correlated significantly with the time elapsed since vaccination, and nearly all nonsignificant trends (except for the levels of fatigue) were beneficial. SARS-CoV-2 variants differ in the severity and nature of their long-term impacts on health and cognitive performance. The Wuhan variant had the most severe long-term consequences, followed by the Alpha variant, with the Delta variant showing intermediate effects. The Omicron variant had the mildest impact on physical health and cognition, though it may have a relatively stronger effect on mental health. These differences are not related to variations in the clinical course of the original acute illness or the time elapsed since COVID-19. Individuals who contracted COVID-19 after being vaccinated are better off in terms of health and cognition compared to those who contracted it before vaccination.

The study also highlighted the need for caution when interpreting the results of cross-sectional studies. Since the likelihood of infection and adherence to preventive measures, including vaccination, often correlate with individuals’ overall health status and their perception of the risks associated with contracting the illness, observational studies may sometimes find that infected and unvaccinated individuals exhibit better health in certain parameters compared to those who are uninfected and vaccinated. Given that randomized experimental studies are rarely ethically permissible in human medicine to assess the impact of infection or vaccination on health, prospective case-control studies should be prioritized. In observational studies, it is essential to consider the Bradford Hill criteria for causality [37], a set of principles that help assess whether observed associations are likely to be causal [38]. In particular, the dose-response criterion highlights the need to examine correlations between health outcomes and both the severity of illness and the time elapsed since illness or vaccination.

## Data Availability

All data produced are available online at Figshare 10.6084/m9.figshare.27618876

https://doi.org/10.6084/m9.figshare.27618876

## Acknowledgments

This research was supported by the Czech Science Foundation, grant number 22-20785S.

## Supplementary materials

**Table S1.**
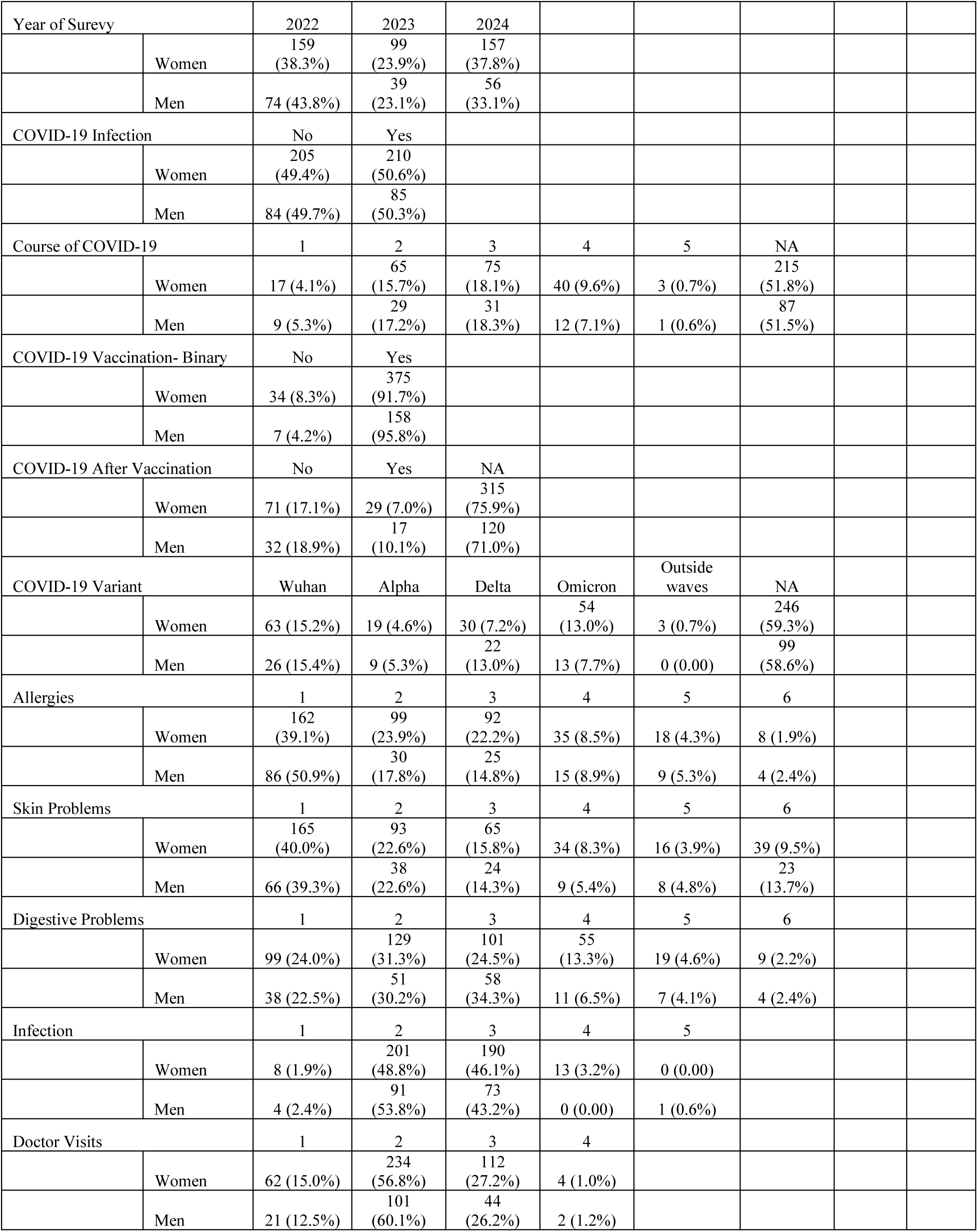
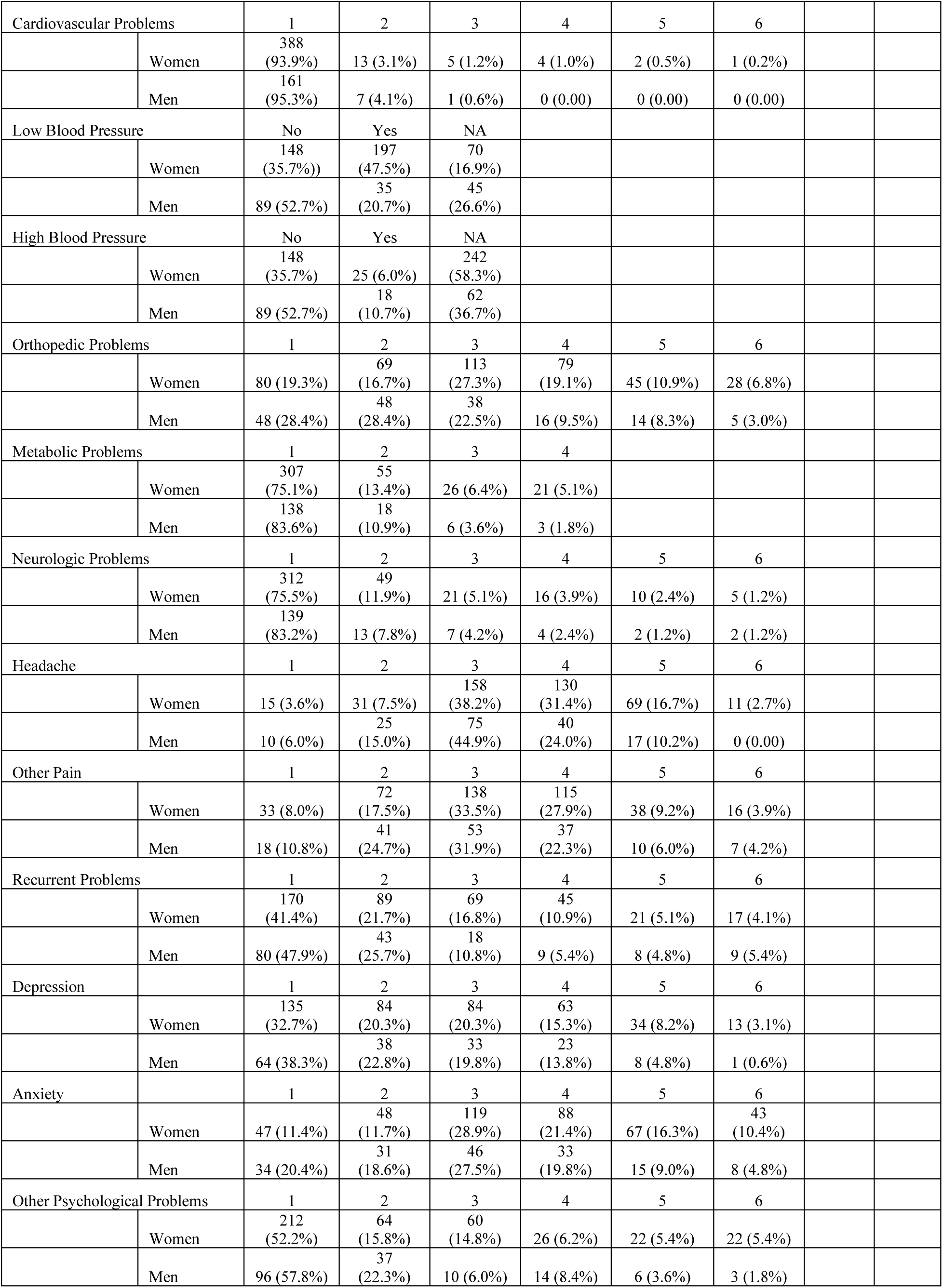
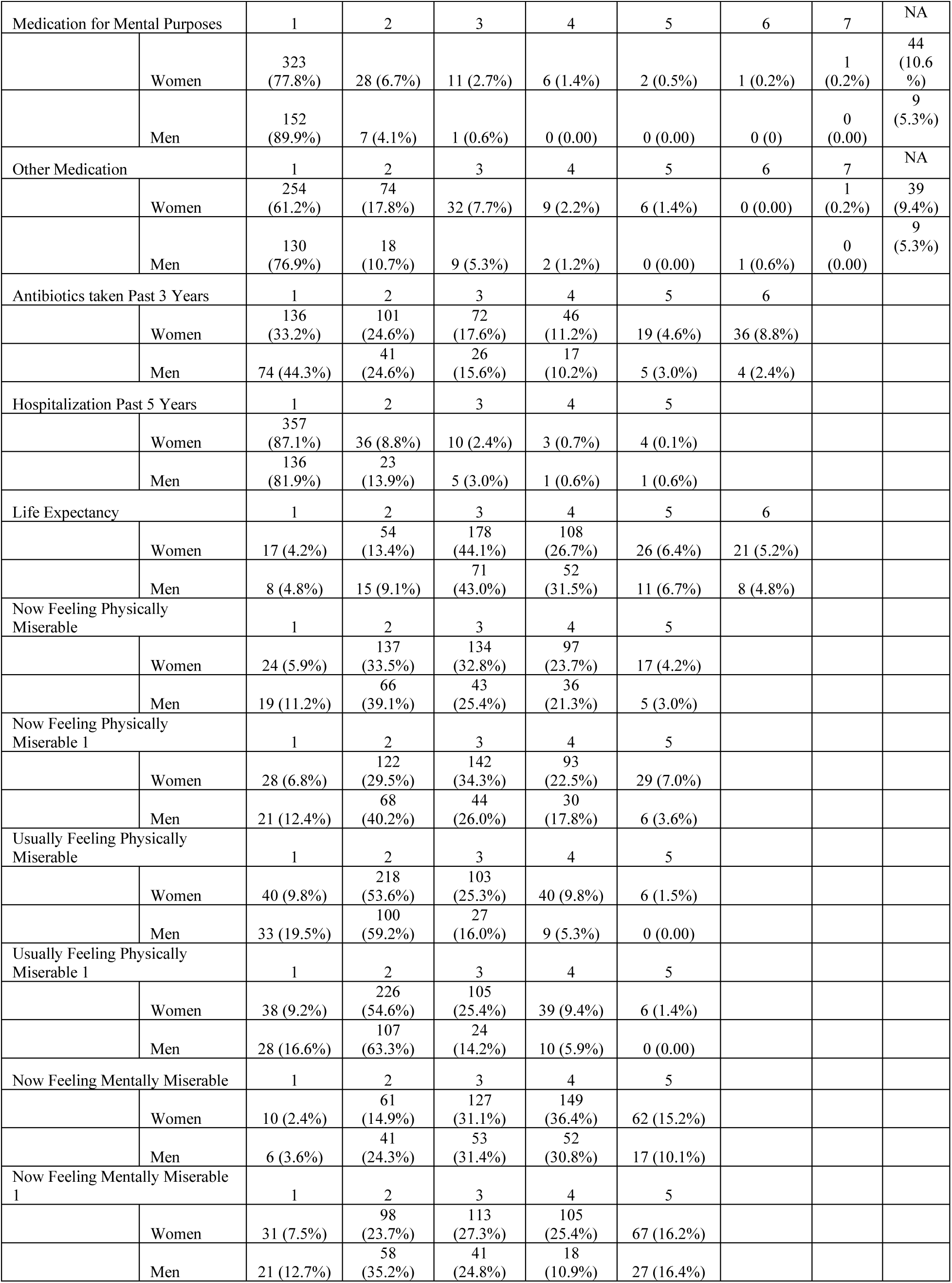
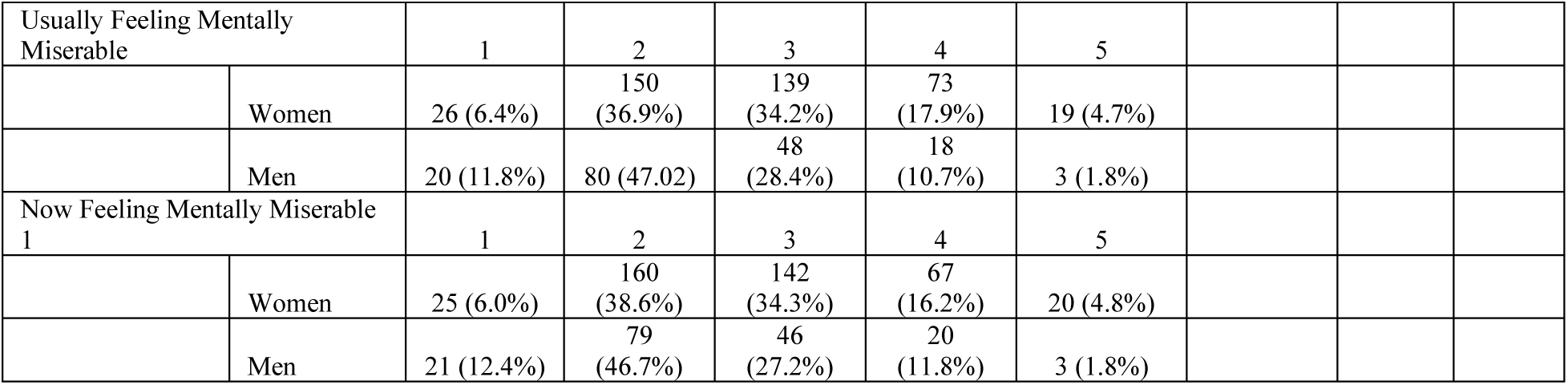
Distribution of data for the levels of ordinal variables.

**Table S2.** Correlations between health and performance-related variables and COVID-related variables controlled for age, sex, and survey year-Women.

| Correlations |  |  |  |  |  |  |  |  |  |  |  |
| --- | --- | --- | --- | --- | --- | --- | --- | --- | --- | --- | --- |
|  | Infected | Course | Months since Infection | Vaccination | Months since Vaccination | Covid after Vaccination | Wuhan | Alpha | Delta | Omicron | Age |
| Physical Health Issues | 0.022 | **0.208** | -0.002 | **0.078** | 0.045 | 0.061 | 0.072 | -0.058 | -0.001 | -0.025 | 0.006 |
| Mental Health Issues | -0.038 | 0.04 | -0.052 | 0.059 | 0.025 | **-0.02** | -0.042 | 0.001 | -0.063 | 0.078 | 0.034 |
| Fatigue | 0.002 | 0.078 | 0.038 | **0.106** | 0.061 | 0.051 | **0.141** | **-0.11** | **-0.108** | 0.019 | 0.013 |
| Intelligence | 0.024 | -0.074 | -0.098 | -0.023 | 0.032 | 0.203 | **-0.149** | -0.044 | 0.033 | **0.189** | -0.02 |
| Memory | **0.065** | -0.015 | -0.027 | -0.032 | 0.002 | 0.066 | -0.089 | 0.006 | 0.003 | 0.049 | -0.036 |
| Reactions | 0.044 | 0.027 | 0.045 | **-0.076** | 0.009 | -0.053 | 0.063 | -0.041 | 0.004 | -0.057 | 0.01 |
| Accuracy | -0.004 | -0.079 | -0.028 | -0.006 | 0.072 | **0.265** | -0.016 | **-0.11** | 0.076 | 0.037 | -0.048 |
| Output Variables |  |  |  |  |  |  |  |  |  |  |  |
| Evolutionary Biology Score | -0.013 | 0.020 | 0.057 | 0.026 | 0.077 | **0.139** | 0.035 | 0.065 | 0.052 | **-0.120** | **-0.089** |
| Free-Recall Memory | **0.088** | -0.032 | -0.040 | 0.007 | -0.000 | 0.120 | -0.045 | -0.003 | -0.053 | 0.055 | -0.048 |
| Recognition Memory | **0.083** | -0.026 | -0.044 | -0.001 | 0.001 | 0.126 | -0.058 | -0.024 | -0.017 | 0.053 | -0.036 |
| Simple RT Duration | 0.016 | -0.073 | 0.049 | **-0.069** | 0.045 | 0.060 | 0.051 | -0.037 | -0.027 | -0.027 | 0.045 |
| Simple RT Precision | 0.000 | -0.010 | -0.017 | -0.056 | 0.038 | 0.009 | 0.020 | **-0.232** | 0.084 | 0.062 | -0.010 |
| Stroop Duration | 0.028 | 0.037 | 0.050 | -0.059 | -0.015 | -0.048 | 0.063 | -0.036 | 0.065 | **-0.104** | 0.015 |
| Stroop Precision | -0.023 | -0.047 | -0.044 | 0.004 | **0.088** | **0.222** | 0.039 | **-0.111** | -0.044 | 0.061 | -0.017 |
| Reading Time | 0.007 | -0.053 | 0.047 | -0.052 | -0.028 | -0.053 | 0.042 | 0.018 | -0.097 | -0.004 | -0.021 |
| Allergy | **0.066** | **0.098** | 0.011 | **0.079** | 0.021 | 0.034 | 0.059 | -0.016 | -0.007 | -0.018 | 0.014 |
| Skin Problem | 0.026 | **0.183** | 0.011 | 0.035 | -0.018 | **0.139** | 0.048 | **-0.134** | 0.047 | 0.006 | -0.016 |
| Digestive Problem | 0.013 | **0.171** | -0.033 | 0.064 | 0.035 | -0.076 | 0.036 | 0.042 | -0.081 | 0.022 | 0.019 |
| Infection | **0.153** | **0.217** | -0.034 | 0.002 | -0.006 | **0.166** | 0.043 | **-0.219** | 0.090 | 0.062 | -0.047 |
| Cardiovascular Problem | 0.032 | -0.016 | 0.045 | **0.085** | -0.031 | -0.078 | 0.095 | -0.079 | -0.047 | 0.004 | -0.036 |
| Low Blood Pressure | -0.045 | 0.031 | 0.070 | 0.017 | 0.066 | -0.100 | 0.083 | 0.008 | -0.071 | -0.043 | -0.018 |
| High Blood Pressure | -0.039 | 0.084 | 0.103 | -0.001 | 0.118 | 0.161 | 0.009 | 0.100 | 0.069 | -0.127 | 0.057 |
| Orthopedic Problem | 0.027 | 0.005 | 0.057 | **0.066** | 0.053 | 0.002 | **0.102** | 0.092 | -0.075 | **-0.118** | 0.026 |
| Metabolic Problem | 0.052 | -0.008 | 0.008 | -0.003 | 0.001 | 0.050 | -0.010 | -0.047 | **0.108** | -0.060 | 0.016 |
| Neurologic Problem | 0.052 | **0.118** | -0.045 | -0.051 | 0.034 | **0.135** | 0.005 | -0.040 | 0.035 | 0.0121 | 0.035 |
| Headache | 0.010 | **0.138** | 0.0177 | **0.104** | -0.034 | 0.109 | 0.079 | -0.022 | -0.030 | -0.060 | -0.049 |
| Other Pain | -0.008 | **0.105** | 0.016 | 0.011 | 0.020 | 0.023 | **0.102** | -0.053 | 0.029 | -0.084 | -0.064 |
| Recurrent Problem | 0.036 | 0.044 | 0.002 | 0.052 | 0.030 | 0.104 | **0.160** | -0.060 | -0.093 | -0.010 | 0.048 |
| Depression | 0.020 | 0.068 | -0.095 | 0.028 | 0.033 | 0.004 | -0.077 | -0.060 | -0.039 | **0.150** | **0.072** |
| Anxiety | 0.004 | 0.055 | -0.031 | 0.057 | 0.034 | 0.086 | 0.060 | -0.091 | **-0.122** | **0.120** | 0.034 |
| Other Psychological Problems | -0.008 | 0.086 | 0.052 | 0.027 | 0.019 | -0.012 | 0.084 | -0.035 | -0.039 | -0.020 | -0.008 |
| Medication for Mental Purposes | 0.003 | **0.167** | **-0.114** | 0.045 | -0.002 | -0.010 | -0.107 | -0.013 | 0.017 | **0.113** | 0.029 |
| Doctor Visits | **0.066** | **0.120** | **-0.127** | 0.038 | 0.002 | 0.078 | -0.071 | -0.019 | 0.050 | 0.088 | -0.026 |
| Other Medication | -0.046 | **0.152** | -0.025 | **0.143** | 0.009 | 0.068 | 0.011 | **-0.131** | 0.016 | 0.043 | 0.015 |
| Antibiotic Taken Past 3 Years | **0.083** | **0.100** | -0.011 | 0.023 | 0.047 | 0.062 | -0.007 | -0.034 | 0.045 | 0.025 | 0.004 |
| Hospitalization Past 5 Years | -0.012 | 0.082 | 0.007 | 0.026 | 0.028 | -0.048 | 0.085 | -0.029 | 0.000 | -0.063 | 0.015 |
| Life Expectancy | -0.040 | 0.092 | 0.072 | **0.075** | -0.013 | **-0.217** | -0.001 | 0.058 | 0.033 | -0.073 | -0.046 |
| Now Feeling Physically Miserable | -0.028 | 0.066 | -0.040 | 0.017 | 0.039 | **0.151** | -0.061 | -0.014 | 0.015 | 0.054 | 0.048 |
| Usually Feeling Physically Miserable | -0.047 | 0.035 | -0.073 | 0.044 | -0.025 | 0.024 | -0.097 | 0.036 | 0.035 | 0.029 | 0.020 |
| Now Feeling Mentally Miserable | 0.000 | 0.024 | -0.012 | -0.004 | 0.027 | -0.058 | -0.034 | 0.015 | -0.031 | 0.006 | 0.042 |
| Usually Feeling Mentally Miserable | **-0.090** | 0.036 | -0.084 | **0.075** | -0.002 | -0.055 | -0.084 | 0.101 | -0.035 | 0.044 | 0.021 |
| Year | **0.394** | 0.076 | **0.591** | **-0.089** | **0.711** | **0.147** | **-0.165** | **-0.142** | 0.013 | **0.222** | 0.021 |
| Course of COVID-19 | NA | NA | -0.034 | -0.033 | -0.003 | 0.129 | 0.012 | -0.100 | 0.053 | 0.026 | 0.092 |
| Months since COVID Infection | NA | -0.034 | NA | 0.024 | **0.175** | **-0.390** | **0.467** | 0.033 | -0.059 | **-0.509** | -0.011 |

*Significant correlations are bolded. The p-values that remained or turned significant after the application of the Benjamini-Hochberg correction for multiple testing (with FDR set at 0.1) are underlined.*

**Table S3.**
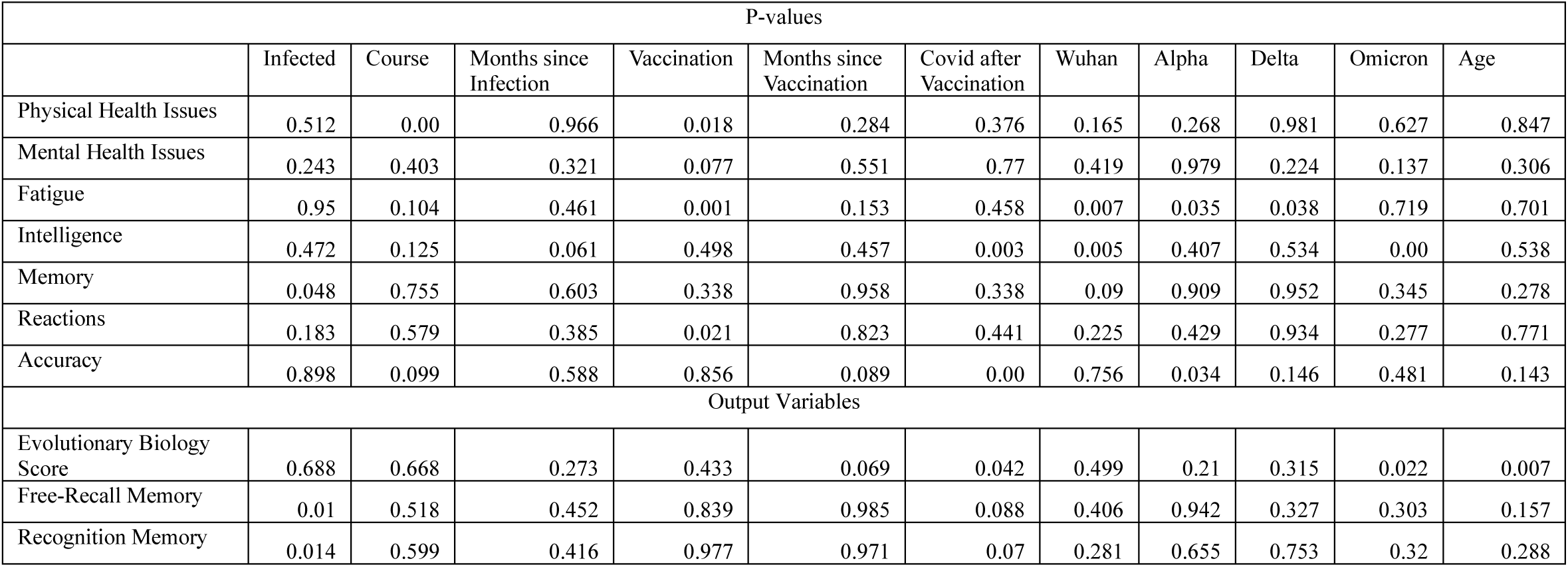
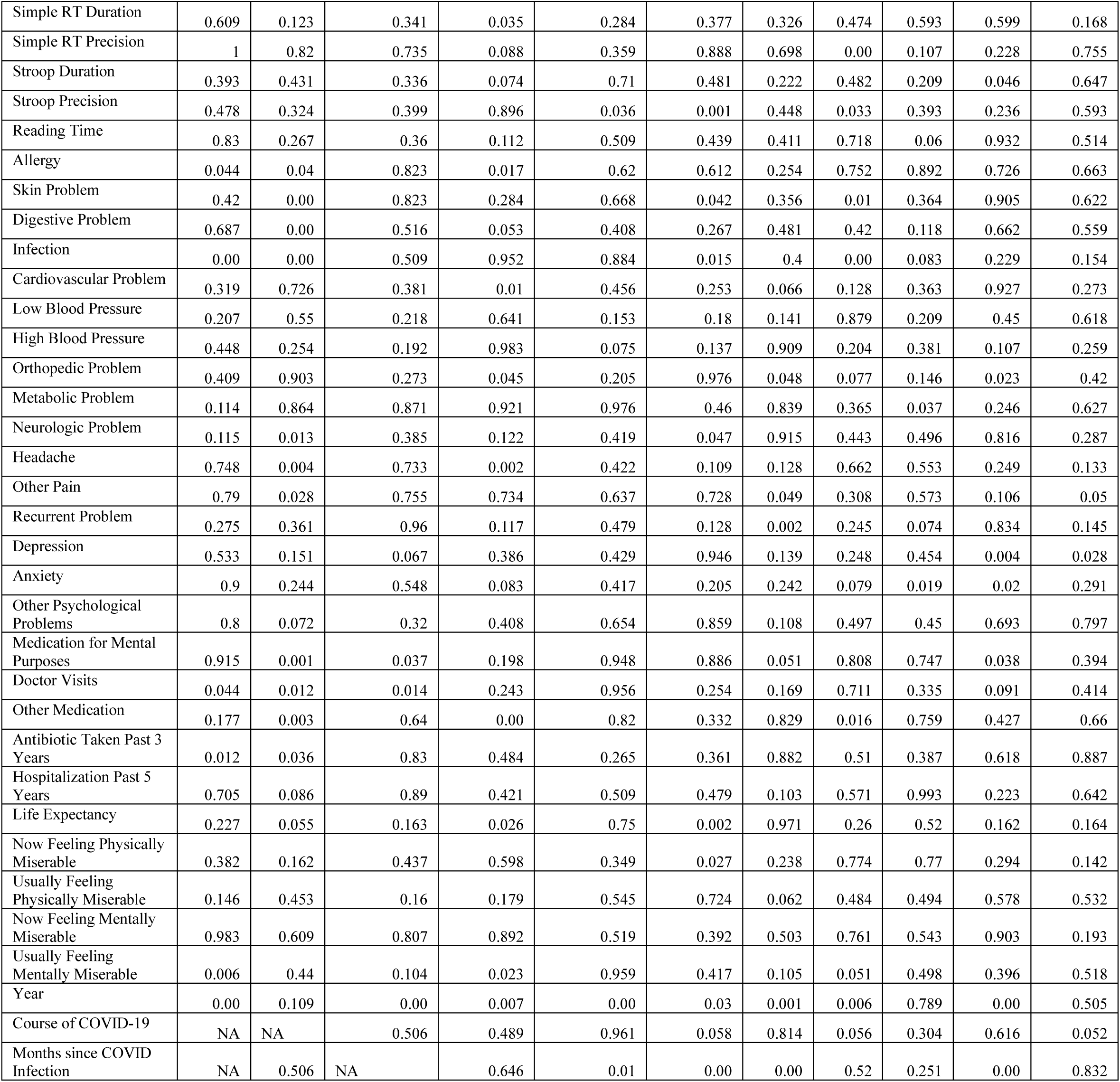
P-values of the correlations between health and performance-related variables and COVID-related variables controlled for age, sex, and survey year-Women.

**Table S4.** Correlations between health and performance-related variables and COVID-related variables controlled for age, sex, and survey year-Men.

| Correlations |  |  |  |  |  |  |  |  |  |  |  |
| --- | --- | --- | --- | --- | --- | --- | --- | --- | --- | --- | --- |
|  | Infected | Course | Months since Infection | Vaccination | Months since Vaccination | Covid after Vaccination | Wuhan | Alpha | Delta | Omicron | Age |
| Physical Health Issues | -0.019 | **0.220** | 0.054 | 0.012 | -0.053 | -0.069 | -0.021 | 0.014 | 0.043 | -0.038 | 0.020 |
| Mental Health Issues | -0.075 | 0.108 | -0.033 | 0.042 | -0.107 | 0.002 | -0.099 | 0.065 | 0.082 | -0.033 | 0.036 |
| Fatigue | **-0.154** | 0.139 | 0.108 | 0.029 | -0.021 | -0.111 | 0.141 | -0.105 | 0.013 | -0.104 | -0.053 |
| Intelligence | 0.009 | -0.007 | 0.034 | **-0.133** | -0.048 | 0.088 | 0.031 | -0.163 | -0.006 | 0.113 | -0.024 |
| Memory | -0.000 | -0.015 | -0.123 | -0.066 | -0.047 | -0.059 | **-0.166** | -0.157 | **0.187** | 0.117 | -0.098 |
| Reactions | -0.003 | 0.028 | 0.008 | -0.073 | 0.048 | -0.073 | **0.176** | **-0.230** | -0.070 | 0.067 | -0.053 |
| Accuracy | -0.00 | -0.090 | -0.008 | -0.052 | -0.006 | 0.048 | -0.041 | 0.008 | -0.137 | **0.215** | -0.089 |
| Output Variables |  |  |  |  |  |  |  |  |  |  |  |
| Evolutionary Biology Score | -0.019 | -0.007 | -0.051 | 0.017 | 0.048 | 0.060 | -0.073 | 0.022 | -0.083 | **0.176** | **-0.130** |
| Free-Recall Memory | -0.038 | 0.029 | -0.134 | -0.054 | -0.052 | -0.058 | **-0.216** | -0.146 | **0.264** | 0.069 | -0.059 |
| Recognition Memory | -0.013 | 0.005 | -0.109 | -0.062 | -0.049 | -0.065 | **-0.177** | **-0.204** | **0.290** | 0.063 | -0.056 |
| Simple RT Duration | -0.011 | -0.057 | 0.027 | -0.026 | 0.040 | -0.085 | 0.088 | -0.051 | -0.057 | 0.004 | 0.003 |
| Simple RT Precision | 0.056 | NA | NA | -0.007 | 0.061 | NA | NA | NA | NA | NA | 0.001 |
| Stroop Duration | -0.013 | 0.044 | 0.010 | -0.025 | 0.076 | -0.028 | 0.114 | -0.118 | -0.071 | 0.047 | -0.021 |
| Stroop Precision | 0.036 | **-0.259** | 0.059 | -0.067 | -0.054 | -0.105 | 0.050 | 0.126 | -0.126 | -0.019 | -0.012 |
| Reading Time | **0.103** | 0.085 | 0.005 | -0.034 | 0.012 | -0.114 | 0.086 | **-0.202** | 0.128 | -0.090 | 0.037 |
| Allergy | -0.090 | **0.201** | 0.020 | 0.026 | 0.017 | 0.031 | 0.005 | 0.031 | 0.079 | -0.133 | -0.064 |
| Skin Problem | -0.006 | 0.119 | 0.003 | -0.048 | 0.119 | 0.131 | -0.037 | 0.090 | 0.003 | -0.037 | -0.054 |
| Digestive Problem | -0.053 | 0.006 | 0.000 | 0.067 | 0.052 | 0.015 | -0.078 | 0.028 | 0.044 | 0.019 | 0.041 |
| Infection | -0.033 | 0.121 | 0.103 | **0.130** | -0.048 | -0.113 | 0.106 | 0.080 | -0.130 | -0.045 | -0.078 |
| Cardiovascular Problem | -0.036 | 0.089 | -0.023 | 0.040 | -0.037 | 0.175 | -0.120 | **0.220** | -0.104 | 0.086 | -0.014 |
| Low Blood Pressure | -0.056 | -0.014 | 0.029 | **0.140** | -0.008 | -0.111 | 0.019 | 0.030 | 0.122 | **-0.221** | 0.006 |
| High Blood Pressure | -0.094 | -0.137 | -0.093 | 0.097 | 0.052 | 0.158 | -0.143 | -0.142 | 0.101 | 0.173 | **0.129** |
| Orthopedic Problem | 0.027 | 0.148 | -0.040 | -0.028 | -0.046 | -0.004 | **-0.225** | **0.168** | 0.077 | 0.040 | **0.105** |
| Metabolic Problem | -0.010 | **0.199** | 0.083 | -0.073 | -0.073 | -0.132 | 0.043 | 0.108 | -0.056 | -0.082 | 0.032 |
| Neurologic Problem | 0.082 | 0.144 | 0.028 | -0.003 | 0.009 | -0.056 | -0.016 | -0.098 | -0.033 | 0.149 | -0.054 |
| Headache | -0.047 | 0.064 | 0.042 | **0.113** | -0.009 | -0.119 | -0.147 | **0.200** | -0.001 | 0.011 | -0.034 |
| Other Pain | -0.053 | **0.191** | 0.044 | -0.012 | -0.033 | -0.001 | -0.019 | 0.089 | -0.053 | 0.011 | 0.047 |
| Recurrent Problem | -0.003 | **0.173** | 0.051 | 0.080 | -0.098 | -0.041 | 0.048 | 0.076 | -0.066 | -0.048 | -0.011 |
| Depression | -0.021 | 0.143 | -0.037 | -0.034 | **-0.128** | -0.174 | -0.070 | 0.042 | -0.005 | 0.058 | 0.052 |
| Anxiety | -0.026 | 0.101 | -0.120 | 0.018 | -0.070 | 0.055 | **-0.182** | 0.017 | 0.089 | 0.106 | 0.036 |
| Other Psychological Problems | **-0.106** | **0.187** | -0.072 | 0.029 | -0.047 | -0.080 | -0.120 | 0.122 | -0.067 | 0.126 | 0.050 |
| Medication for Mental Purposes | **-0.118** | **0.154** | 0.065 | 0.042 | 0.028 | -0.136 | 0.023 | -0.054 | 0.092 | -0.093 | 0.059 |
| Doctor Visits | -0.000 | **0.194** | 0.025 | -0.031 | 0.055 | -0.165 | 0.028 | **0.199** | **-0.206** | 0.042 | **-0.103** |
| Other Medication | -0.068 | **0.273** | 0.001 | 0.019 | -0.013 | 0.125 | 0.021 | 0.088 | **-0.184** | 0.116 | -0.019 |
| Antibiotic Taken Past 3 Years | 0.010 | 0.099 | -0.009 | -0.077 | 0.017 | -0.045 | -0.019 | 0.027 | -0.023 | 0.030 | -0.057 |
| Hospitalization Past 5 Years | **0.112** | **0.176** | 0.045 | **-0.106** | -0.055 | -0.097 | -0.018 | 0.047 | 0.075 | -0.112 | **-0.114** |
| Life Expectancy | 0.040 | 0.090 | 0.017 | -0.091 | -0.044 | 0.039 | 0.038 | **-0.183** | 0.115 | -0.029 | 0.037 |
| Now Feeling Physically Miserable | 0.049 | 0.072 | 0.125 | 0.007 | -0.111 | **-0.233** | 0.103 | 0.002 | -0.058 | -0.062 | **0.130** |
| Usually Feeling Physically Miserable | -0.024 | 0.071 | -0.018 | -0.027 | -0.090 | -0.078 | 0.003 | -0.131 | 0.045 | 0.055 | 0.046 |
| Now Feeling Mentally Miserable | -0.060 | 0.086 | 0.053 | 0.002 | **-0.148** | 0.069 | 0.014 | 0.010 | 0.125 | **-0.183** | 0.043 |
| Usually Feeling Mentally Miserable | -0.084 | 0.018 | -0.020 | 0.084 | -0.039 | -0.1747 | -0.046 | 0.155 | -0.027 | -0.045 | 0.014 |
| Year | **0.315** | **0.178** | **0.684** | **-0.123** | **0.710** | -0.011 | -0.107 | -0.112 | -0.066 | **0.306** | 0.009 |
| Course of COVID-19 | NA | NA | 0.065 | -0.042 | -0.002 | -0.147 | 0.084 | 0.125 | -0.112 | -0.080 | 0.129 |
| Months since COVID Infection | -0.078 | 0.065 | NA | 0.086 | 0.067 | **-0.441** | **0.485** | 0.035 | **-0.223** | **-0.374** | 0.031 |

*Significant correlations are bolded. The p-values that remained or turned significant after the application of the Benjamini-Hochberg correction for multiple testing (with FDR set at 0.1) are underlined.*

**Table S5.** P-values of the correlations between health and performance-related variables and COVID-related variables controlled for age, sex, and survey year-Men.

|  | P-values |  |  |  |  |  |  |  |  |  |  |
| --- | --- | --- | --- | --- | --- | --- | --- | --- | --- | --- | --- |
|  | Infected | Course | Months since Infection | Vaccination | Months since Vaccination | Covid after Vaccination | Wuhan | Alpha | Delta | Omicron | Age |
| Physical Health Issues | 0.713 | 0.004 | 0.51 | 0.819 | 0.404 | 0.49 | 0.797 | 0.866 | 0.603 | 0.639 | 0.687 |
| Mental Health Issues | 0.149 | 0.155 | 0.685 | 0.419 | 0.096 | 0.983 | 0.229 | 0.432 | 0.318 | 0.684 | 0.485 |
| Fatigue | 0.003 | 0.069 | 0.19 | 0.575 | 0.743 | 0.27 | 0.087 | 0.205 | 0.87 | 0.21 | 0.304 |
| Intelligence | 0.851 | 0.924 | 0.678 | 0.013 | 0.454 | 0.381 | 0.706 | 0.05 | 0.939 | 0.176 | 0.648 |
| Memory | 0.994 | 0.843 | 0.14 | 0.206 | 0.467 | 0.56 | 0.047 | 0.06 | 0.025 | 0.16 | 0.06 |
| Reactions | 0.953 | 0.711 | 0.92 | 0.164 | 0.448 | 0.469 | 0.033 | 0.005 | 0.398 | 0.417 | 0.306 |
| Accuracy | 1 | 0.234 | 0.916 | 0.315 | 0.926 | 0.629 | 0.616 | 0.916 | 0.098 | 0.009 | 0.084 |
| Output Variables |  |  |  |  |  |  |  |  |  |  |  |
| Evolutionary Biology Score | 0.705 | 0.92 | 0.535 | 0.737 | 0.449 | 0.551 | 0.378 | 0.787 | 0.315 | 0.033 | 0.012 |
| Free-Recall Memory | 0.482 | 0.713 | 0.123 | 0.332 | 0.435 | 0.584 | 0.013 | 0.092 | 0.002 | 0.423 | 0.278 |
| Recognition Memory | 0.813 | 0.942 | 0.221 | 0.254 | 0.46 | 0.539 | 0.047 | 0.022 | 0.001 | 0.48 | 0.301 |
| Simple RT Duration | 0.823 | 0.454 | 0.745 | 0.616 | 0.527 | 0.396 | 0.287 | 0.532 | 0.49 | 0.955 | 0.939 |
| Simple RT Precision | 0.278 | NA | NA | 0.892 | 0.34 | NA | NA | NA | NA | NA | 0.977 |
| Stroop Duration | 0.79 | 0.558 | 0.898 | 0.626 | 0.237 | 0.778 | 0.168 | 0.154 | 0.392 | 0.566 | 0.685 |
| Stroop Precision | 0.488 | 0.001 | 0.474 | 0.202 | 0.396 | 0.295 | 0.546 | 0.127 | 0.127 | 0.817 | 0.815 |
| Reading Time | 0.047 | 0.26 | 0.944 | 0.513 | 0.846 | 0.258 | 0.295 | 0.015 | 0.12 | 0.274 | 0.47 |
| Allergy | 0.083 | 0.008 | 0.802 | 0.62 | 0.785 | 0.756 | 0.946 | 0.702 | 0.34 | 0.107 | 0.212 |
| Skin Problem | 0.905 | 0.116 | 0.971 | 0.362 | 0.065 | 0.192 | 0.653 | 0.276 | 0.965 | 0.651 | 0.294 |
| Digestive Problem | 0.303 | 0.936 | 0.997 | 0.202 | 0.418 | 0.879 | 0.343 | 0.73 | 0.588 | 0.818 | 0.424 |
| Infection | 0.52 | 0.11 | 0.21 | 0.013 | 0.457 | 0.261 | 0.198 | 0.332 | 0.115 | 0.581 | 0.132 |
| Cardiovascular Problem | 0.482 | 0.242 | 0.779 | 0.438 | 0.566 | 0.082 | 0.147 | 0.008 | 0.208 | 0.296 | 0.783 |
| Low Blood Pressure | 0.355 | 0.87 | 0.751 | 0.023 | 0.909 | 0.34 | 0.839 | 0.75 | 0.195 | 0.019 | 0.92 |
| High Blood Pressure | 0.152 | 0.143 | 0.345 | 0.146 | 0.515 | 0.187 | 0.147 | 0.148 | 0.302 | 0.079 | 0.05 |
| Orthopedic Problem | 0.596 | 0.051 | 0.623 | 0.586 | 0.473 | 0.966 | 0.007 | 0.042 | 0.348 | 0.623 | 0.043 |
| Metabolic Problem | 0.838 | 0.009 | 0.315 | 0.167 | 0.263 | 0.188 | 0.598 | 0.193 | 0.497 | 0.323 | 0.539 |
| Neurologic Problem | 0.115 | 0.059 | 0.736 | 0.954 | 0.883 | 0.574 | 0.847 | 0.238 | 0.692 | 0.074 | 0.296 |
| Headache | 0.36 | 0.398 | 0.609 | 0.032 | 0.885 | 0.236 | 0.078 | 0.017 | 0.985 | 0.893 | 0.514 |
| Other Pain | 0.308 | 0.012 | 0.594 | 0.815 | 0.601 | 0.986 | 0.812 | 0.286 | 0.522 | 0.893 | 0.367 |
| Recurrent Problem | 0.953 | 0.024 | 0.535 | 0.128 | 0.129 | 0.681 | 0.562 | 0.363 | 0.428 | 0.559 | 0.827 |
| Depression | 0.686 | 0.059 | 0.652 | 0.519 | 0.048 | 0.084 | 0.397 | 0.606 | 0.949 | 0.484 | 0.315 |
| Anxiety | 0.615 | 0.184 | 0.146 | 0.727 | 0.275 | 0.582 | 0.028 | 0.837 | 0.281 | 0.2 | 0.48 |
| Other Psychological Problems | 0.044 | 0.014 | 0.385 | 0.582 | 0.467 | 0.424 | 0.149 | 0.141 | 0.421 | 0.129 | 0.332 |
| Medication for Mental Purposes | 0.027 | 0.048 | 0.442 | 0.431 | 0.663 | 0.188 | 0.779 | 0.519 | 0.274 | 0.269 | 0.266 |
| Doctor Visits | 0.99 | 0.011 | 0.758 | 0.551 | 0.392 | 0.1 | 0.732 | 0.016 | 0.013 | 0.61 | 0.047 |
| Other Medication | 0.199 | 0.001 | 0.99 | 0.721 | 0.84 | 0.226 | 0.8 | 0.296 | 0.03 | 0.17 | 0.711 |
| Antibiotic Taken Past 3 Years | 0.841 | 0.191 | 0.912 | 0.144 | 0.781 | 0.65 | 0.813 | 0.743 | 0.776 | 0.716 | 0.275 |
| Hospitalization Past 5 Years | 0.032 | 0.02 | 0.584 | 0.045 | 0.397 | 0.335 | 0.824 | 0.568 | 0.362 | 0.175 | 0.029 |
| Life Expectancy | 0.439 | 0.242 | 0.837 | 0.084 | 0.493 | 0.702 | 0.643 | 0.028 | 0.165 | 0.725 | 0.477 |
| Now Feeling Physically Miserable | 0.346 | 0.338 | 0.13 | 0.887 | 0.084 | 0.021 | 0.21 | 0.98 | 0.483 | 0.454 | 0.012 |
| Usually Feeling Physically Miserable | 0.643 | 0.351 | 0.822 | 0.6 | 0.16 | 0.439 | 0.963 | 0.113 | 0.581 | 0.506 | 0.37 |
| Now Feeling Mentally Miserable | 0.243 | 0.255 | 0.515 | 0.961 | 0.021 | 0.489 | 0.859 | 0.901 | 0.131 | 0.027 | 0.4 |
| Usually Feeling Mentally Miserable | 0.103 | 0.805 | 0.807 | 0.111 | 0.539 | 0.083 | 0.578 | 0.061 | 0.741 | 0.582 | 0.783 |
| Year | 0.00 | 0.019 | 0.00 | 0.018 | 0.00 | 0.904 | 0.191 | 0.174 | 0.418 | 0.00 | 0.848 |
| Course of COVID-19 | NA | NA | 0.439 | 0.585 | 0.983 | 0.148 | 0.316 | 0.137 | 0.181 | 0.337 | 0.088 |
| Months since COVID Infection | 0.347 | 0.439 | NA | 0.311 | 0.503 | 0.00 | 0.00 | 0.668 | 0.007 | 0.00 | 0.7 |

**Table S6.** Correlations between health and performance-related variables and COVID-related variables controlled for age, sex, survey year, and SARS-CoV-2 variants-All subjects.

| Correlations |  |  |  |  |  |  |  |  |  |  |  |  |
| --- | --- | --- | --- | --- | --- | --- | --- | --- | --- | --- | --- | --- |
|  | Infected | Course | Months since Infection | Vaccination | Months since Vaccination | Covid after Vaccination | Wuhan | Alpha | Delta | Omicron | Age | Sex |
| Physical Health Issues | 0.011 | **<u>0.210</u>** | -0.009 | **0.060** | 0.012 | 0.010 | 0.031 | -0.026 | 0.009 | 0.000 | 0.011 | **<u>0.162</u>** |
| Mental Health Issues | -0.047 | 0.066 | -0.021 | 0.050 | -0.014 | -0.042 | -0.017 | 0.021 | -0.034 | 0.021 | 0.036 | **<u>0.162</u>** |
| Fatigue | -0.043 | **<u>0.092</u>** | 0.009 | **<u>0.086</u>** | 0.034 | 0.047 | **0.119** | **-0.107** | -0.048 | 0.065 | -0.005 | **<u>0.096</u>** |
| Intelligence | 0.025 | -0.056 | 0.011 | -0.035 | 0.009 | 0.099 | 0.039 | -0.068 | -0.014 | **0.088** | -0.020 | **<u>-0.099</u>** |
| Memory | 0.047 | -0.008 | 0.006 | -0.031 | -0.007 | -0.053 | -0.052 | -0.028 | 0.042 | -0.018 | -0.054 | **<u>0.111</u>** |
| Reactions | 0.027 | 0.028 | 0.005 | **<u>-0.079</u>** | 0.025 | -0.018 | 0.082 | **-0.098** | -0.012 | 0.029 | -0.009 | 0.020 |
| Accuracy | 0.003 | **<u>-0.081</u>** | 0.004 | -0.010 | 0.054 | **<u>0.174</u>** | 0.033 | -0.070 | -0.001 | 0.051 | -0.052 | **<u>-0.095</u>** |
| Output Variables |  |  |  |  |  |  |  |  |  |  |  |  |
| Evolutionary Biology Score | -0.014 | 0.008 | 0.015 | 0.023 | 0.066 | **0.126** | -0.036 | 0.050 | 0.017 | -0.039 | **-0.099** | **-0.073** |
| Free-Recall Memory | 0.053 | -0.017 | -0.018 | 0.001 | -0.011 | -0.004 | -0.038 | -0.036 | 0.036 | -0.016 | -0.051 | **0.107** |
| Recognition Memory | **0.059** | -0.018 | -0.013 | -0.005 | -0.007 | -0.004 | -0.030 | -0.068 | 0.064 | -0.025 | -0.041 | **0.115** |
| Simple RT Duration | 0.003 | -0.069 | 0.020 | **-0.060** | 0.041 | 0.065 | 0.049 | -0.051 | -0.028 | 0.022 | 0.034 | 0.045 |
| Simple RT Precision | 0.009 | -0.008 | 0.006 | -0.044 | 0.045 | 0.059 | **0.090** | **-0.191** | 0.059 | 0.032 | -0.006 | **-0.074** |
| Stroop Duration | 0.013 | 0.039 | 0.000 | -0.054 | 0.010 | 0.009 | 0.048 | -0.072 | 0.0311 | -0.017 | 0.000 | -0.002 |
| Stroop Precision | -0.006 | **-0.094** | -0.026 | -0.009 | 0.045 | **0.110** | 0.060 | -0.042 | -0.070 | 0.070 | -0.015 | -0.007 |
| Reading Time | 0.039 | -0.011 | 0.025 | -0.053 | -0.011 | -0.049 | 0.033 | -0.055 | -0.008 | 0.006 | -0.002 | 0.053 |
| Allergy | 0.016 | **0.127** | -0.014 | **0.066** | 0.018 | 0.074 | 0.014 | -0.001 | 0.031 | -0.022 | -0.006 | **0.069** |
| Skin Problem | 0.016 | **0.163** | 0.004 | 0.017 | 0.029 | 0.094 | 0.025 | -0.055 | 0.031 | -0.002 | -0.029 | -0.017 |
| Digestive Problem | -0.003 | **0.125** | -0.022 | **0.063** | 0.039 | -0.059 | 0.005 | 0.040 | -0.041 | 0.032 | 0.025 | 0.004 |
| Infection | **0.102** | **0.189** | 0.00 | 0.031 | -0.013 | 0.039 | **0.102** | **-0.127** | 0.017 | 0.052 | **-0.055** | **0.055** |
| Cardiovascular Problem | 0.015 | 0.003 | 0.019 | **0.075** | -0.032 | -0.017 | 0.052 | -0.011 | -0.058 | 0.055 | -0.029 | 0.026 |
| Low Blood Pressure | -0.048 | 0.020 | 0.027 | 0.039 | 0.048 | -0.090 | 0.005 | 0.009 | 0.004 | -0.030 | -0.014 | **0.255** |
| High Blood Pressure | -0.059 | 0.001 | 0.049 | 0.037 | 0.090 | **0.156** | -0.039 | -0.003 | 0.077 | -0.052 | **0.088** | -0.034 |
| Orthopedic Problem | 0.023 | 0.045 | 0.001 | 0.042 | 0.020 | 0.038 | -0.057 | **0.104** | -0.017 | -0.044 | 0.045 | **0.159** |
| Metabolic Problem | 0.039 | 0.042 | 0.015 | -0.017 | -0.019 | -0.001 | -0.029 | -0.011 | 0.070 | -0.068 | 0.021 | **0.095** |
| Neurologic Problem | **0.061** | **0.125** | -0.020 | -0.041 | 0.025 | 0.104 | 0.039 | -0.056 | 0.008 | 0.033 | 0.012 | **0.078** |
| Headache | -0.006 | **0.130** | -0.003 | **0.108** | -0.025 | -0.007 | -0.026 | 0.044 | -0.014 | -0.024 | -0.043 | **0.160** |
| Other Pain | -0.021 | **0.140** | -0.016 | 0.001 | 0.002 | 0.066 | 0.031 | -0.018 | 0.010 | -0.010 | -0.028 | **0.088** |
| Recurrent Problem | 0.024 | 0.079 | -0.031 | 0.050 | -0.013 | 0.103 | **0.094** | -0.022 | -0.068 | 0.068 | 0.029 | **0.070** |
| Depression | 0.010 | **0.089** | -0.034 | 0.019 | -0.013 | -0.071 | 0.015 | -0.023 | -0.053 | 0.076 | **0.069** | **0.081** |
| Anxiety | -0.006 | 0.070 | -0.023 | 0.050 | 0.004 | 0.060 | 0.071 | -0.056 | -0.069 | **0.108** | 0.035 | **0.155** |
| Other Psychological Problems | -0.034 | **0.114** | 0.019 | 0.024 | -0.001 | -0.025 | 0.034 | 0.002 | -0.047 | 0.040 | 0.008 | **0.072** |
| Medication for Mental Purposes | -0.020 | **0.155** | -0.033 | 0.048 | 0.005 | **-0.129** | -0.013 | -0.021 | 0.020 | 0.017 | 0.036 | **0.117** |
| Doctor Visits | 0.046 | **0.144** | -0.065 | 0.024 | 0.015 | -0.050 | 0.002 | 0.051 | -0.050 | 0.058 | -0.049 | -0.009 |
| Other Medication | -0.051 | **0.185** | -0.014 | **0.117** | 0.001 | 0.075 | 0.059 | -0.067 | -0.049 | 0.064 | 0.004 | **0.136** |
| Antibiotic Taken Past 3 Years | **0.064** | **0.099** | 0.004 | 0.004 | 0.034 | 0.012 | 0.012 | -0.012 | 0.017 | 0.016 | -0.015 | **0.117** |
| Hospitalization Past 5 Years | 0.018 | **0.124** | -0.0171 | -0.008 | -0.000 | -0.042 | 0.003 | -0.001 | 0.035 | -0.038 | -0.028 | **-0.058** |
| Life Expectancy | -0.015 | **0.095** | 0.050 | 0.035 | -0.025 | -0.103 | -0.019 | -0.014 | 0.067 | -0.061 | -0.020 | -0.037 |
| Now Feeling Physically Miserable | -0.006 | 0.073 | 0.008 | 0.017 | -0.007 | 0.024 | 0.009 | -0.009 | -0.020 | 0.021 | **0.073** | **0.102** |
| Usually Feeling Physically Miserable | -0.037 | 0.050 | -0.033 | 0.033 | -0.045 | -0.048 | -0.034 | -0.006 | 0.022 | -0.010 | 0.029 | **0.169** |
| Now Feeling Mentally Miserable | -0.021 | 0.053 | -0.006 | -0.006 | -0.028 | -0.022 | -0.042 | 0.009 | 0.026 | -0.051 | 0.043 | **0.128** |
| Usually Feeling Mentally Miserable | **-0.087** | 0.034 | -0.058 | **0.070** | -0.015 | **-0.136** | -0.073 | **0.118** | -0.045 | 0.001 | 0.021 | **0.149** |
| Year | **0.372** | **0.106** | **0.720** | **-0.081** | **0.706** | -0.108 | 0.045 | **-0.117** | -0.064 | **0.133** | 0.019 | 0.048 |
| Course of COVID-19 | NA | NA | -0.020 | -0.037 | -0.003 | 0.018 | 0.037 | -0.034 | 0.002 | 0.017 | **0.101** | 0.050 |
| Months since COVID Infection | -0.012 | -0.020 | NA | -0.015 | 0.097 | **-0.163** | **0.086** | 0.002 | 0.013 | **-0.153** | -0.002 | -0.028 |

*Significant correlations are bolded. The p-values that remained or turned significant after the application of the Benjamini-Hochberg correction for multiple testing (with FDR set at 0.1) are underlined.*

**Table S7.** P-values of the correlations between health and performance-related variables and COVID-related variables controlled for age, sex, survey year, and SARS-CoV-2 variants-All subjects.

|  | P-values |  |  |  |  |  |  |  |  |  |  |  |
| --- | --- | --- | --- | --- | --- | --- | --- | --- | --- | --- | --- | --- |
|  | Infected | Course | Months since Infection | Vaccination | Months since Vaccination | Covid after Vaccination | Wuhan | Alpha | Delta | Omicron | Age | Sex |
| Physical Health Issues | 0.673 | 0.00 | 0.83 | 0.03 | 0.715 | 0.85 | 0.474 | 0.551 | 0.831 | 0.997 | 0.678 | 0.00 |
| Mental Health Issues | 0.084 | 0.1 | 0.616 | 0.07 | 0.682 | 0.446 | 0.697 | 0.628 | 0.429 | 0.628 | 0.188 | 0.00 |
| Fatigue | 0.116 | 0.022 | 0.834 | 0.002 | 0.326 | 0.395 | 0.006 | 0.014 | 0.273 | 0.136 | 0.835 | 0.001 |
| Intelligence | 0.374 | 0.163 | 0.786 | 0.205 | 0.795 | 0.077 | 0.372 | 0.122 | 0.747 | 0.044 | 0.474 | 0.00 |
| Memory | 0.089 | 0.827 | 0.891 | 0.26 | 0.831 | 0.347 | 0.235 | 0.519 | 0.332 | 0.67 | 0.052 | 0.00 |
| Reactions | 0.326 | 0.483 | 0.901 | 0.005 | 0.477 | 0.739 | 0.059 | 0.024 | 0.771 | 0.508 | 0.733 | 0.463 |
| Accuracy | 0.912 | 0.043 | 0.926 | 0.711 | 0.121 | 0.002 | 0.439 | 0.107 | 0.978 | 0.236 | 0.06 | 0.001 |
| Output Variables |  |  |  |  |  |  |  |  |  |  |  |  |
| Evolutionary Biology Score | 0.602 | 0.835 | 0.719 | 0.394 | 0.062 | 0.024 | 0.409 | 0.252 | 0.692 | 0.37 | 0 | 0.009 |
| Free-Recall Memory | 0.065 | 0.683 | 0.685 | 0.972 | 0.75 | 0.942 | 0.402 | 0.422 | 0.421 | 0.714 | 0.079 | 0.00 |
| Recognition Memory | 0.041 | 0.657 | 0.765 | 0.839 | 0.828 | 0.933 | 0.51 | 0.135 | 0.163 | 0.586 | 0.149 | 0.00 |
| Simple RT Duration | 0.889 | 0.086 | 0.643 | 0.03 | 0.24 | 0.24 | 0.262 | 0.244 | 0.519 | 0.613 | 0.217 | 0.1 |
| Simple RT Precision | 0.72 | 0.835 | 0.88 | 0.113 | 0.195 | 0.286 | 0.039 | 0.00 | 0.177 | 0.464 | 0.804 | 0.007 |
| Stroop Duration | 0.622 | 0.326 | 0.994 | 0.05 | 0.755 | 0.86 | 0.267 | 0.096 | 0.477 | 0.692 | 0.983 | 0.938 |
| Stroop Precision | 0.822 | 0.019 | 0.545 | 0.73 | 0.201 | 0.049 | 0.165 | 0.338 | 0.11 | 0.109 | 0.568 | 0.774 |
| Reading Time | 0.157 | 0.766 | 0.566 | 0.055 | 0.755 | 0.373 | 0.442 | 0.205 | 0.852 | 0.877 | 0.931 | 0.055 |
| Allergy | 0.56 | 0.002 | 0.738 | 0.018 | 0.595 | 0.186 | 0.746 | 0.967 | 0.477 | 0.607 | 0.817 | 0.012 |
| Skin Problem | 0.566 | 0.00 | 0.919 | 0.536 | 0.405 | 0.092 | 0.566 | 0.208 | 0.469 | 0.963 | 0.282 | 0.521 |
| Digestive Problem | 0.89 | 0.002 | 0.615 | 0.024 | 0.261 | 0.285 | 0.899 | 0.356 | 0.343 | 0.465 | 0.353 | 0.874 |
| Infection | 0.00 | 0.00 | 0.999 | 0.257 | 0.701 | 0.482 | 0.019 | 0.004 | 0.697 | 0.229 | 0.048 | 0.047 |
| Cardiovascular Problem | 0.567 | 0.922 | 0.651 | 0.007 | 0.358 | 0.759 | 0.232 | 0.795 | 0.181 | 0.204 | 0.291 | 0.349 |
| Low Blood Pressure | 0.116 | 0.647 | 0.571 | 0.211 | 0.219 | 0.148 | 0.903 | 0.845 | 0.933 | 0.524 | 0.642 | 0.00 |
| High Blood Pressure | 0.139 | 0.975 | 0.42 | 0.366 | 0.075 | 0.047 | 0.516 | 0.956 | 0.201 | 0.388 | 0.028 | 0.396 |
| Orthopedic Problem | 0.396 | 0.256 | 0.965 | 0.127 | 0.554 | 0.49 | 0.191 | 0.017 | 0.696 | 0.306 | 0.099 | 0.00 |
| Metabolic Problem | 0.162 | 0.291 | 0.715 | 0.531 | 0.589 | 0.982 | 0.505 | 0.799 | 0.106 | 0.116 | 0.446 | 0.001 |
| Neurologic Problem | 0.027 | 0.002 | 0.638 | 0.143 | 0.471 | 0.063 | 0.365 | 0.195 | 0.843 | 0.449 | 0.66 | 0.005 |
| Headache | 0.825 | 0.001 | 0.941 | 0.00 | 0.469 | 0.895 | 0.553 | 0.305 | 0.742 | 0.583 | 0.12 | 0.00 |
| Other Pain | 0.447 | 0.00 | 0.711 | 0.945 | 0.94 | 0.237 | 0.473 | 0.676 | 0.812 | 0.815 | 0.303 | 0.001 |
| Recurrent Problem | 0.377 | 0.05 | 0.469 | 0.072 | 0.704 | 0.066 | 0.031 | 0.616 | 0.122 | 0.119 | 0.283 | 0.012 |
| Depression | 0.705 | 0.027 | 0.432 | 0.489 | 0.707 | 0.201 | 0.72 | 0.595 | 0.223 | 0.079 | 0.012 | 0.004 |
| Anxiety | 0.821 | 0.081 | 0.594 | 0.071 | 0.908 | 0.284 | 0.101 | 0.196 | 0.112 | 0.013 | 0.207 | 0.00 |
| Other Psychological Problems | 0.224 | 0.005 | 0.652 | 0.395 | 0.962 | 0.655 | 0.435 | 0.949 | 0.282 | 0.363 | 0.767 | 0.01 |
| Medication for Mental Purposes | 0.475 | 0.00 | 0.47 | 0.096 | 0.879 | 0.026 | 0.777 | 0.644 | 0.661 | 0.708 | 0.208 | 0.00 |
| Doctor Visits | 0.095 | 0.00 | 0.133 | 0.391 | 0.652 | 0.367 | 0.956 | 0.236 | 0.248 | 0.183 | 0.075 | 0.74 |
| Other Medication | 0.074 | 0.00 | 0.742 | 0.00 | 0.963 | 0.188 | 0.191 | 0.138 | 0.28 | 0.155 | 0.88 | 0.00 |
| Antibiotic Taken Past 3 Years | 0.022 | 0.013 | 0.91 | 0.863 | 0.325 | 0.821 | 0.782 | 0.773 | 0.696 | 0.705 | 0.582 | 0.00 |
| Hospitalization Past 5 Years | 0.518 | 0.002 | 0.696 | 0.768 | 0.993 | 0.445 | 0.932 | 0.969 | 0.415 | 0.375 | 0.309 | 0.035 |
| Life Expectancy | 0.571 | 0.019 | 0.249 | 0.212 | 0.471 | 0.065 | 0.652 | 0.737 | 0.122 | 0.159 | 0.463 | 0.177 |
| Now Feeling Physically Miserable | 0.813 | 0.066 | 0.843 | 0.54 | 0.838 | 0.657 | 0.833 | 0.835 | 0.64 | 0.631 | 0.008 | 0.00 |
| Usually Feeling Physically Miserable | 0.182 | 0.207 | 0.445 | 0.23 | 0.2 | 0.385 | 0.429 | 0.891 | 0.6 | 0.818 | 0.29 | 0.00 |
| Now Feeling Mentally Miserable | 0.43 | 0.186 | 0.889 | 0.82 | 0.423 | 0.695 | 0.328 | 0.829 | 0.539 | 0.236 | 0.119 | 0.00 |
| Usually Feeling Mentally Miserable | 0.002 | 0.392 | 0.182 | 0.011 | 0.669 | 0.015 | 0.094 | 0.007 | 0.299 | 0.981 | 0.441 | 0.00 |
| Year | 0 | 0.008 | 0.00 | 0.004 | 0.00 | 0.052 | 0.3 | 0.007 | 0.137 | 0.002 | 0.474 | 0.08 |
| Course of COVID-19 | NA | NA | 0.644 | 0.356 | 0.948 | 0.737 | 0.4 | 0.433 | 0.962 | 0.697 | 0.012 | 0.21 |
| Months since COVID Infection | 0.767 | 0.644 | NA | 0.72 | 0.083 | 0.004 | 0.05 | 0.952 | 0.758 | 0.00 | 0.946 | 0.52 |

**Table S8.** Correlations between health and performance-related variables and COVID-related variables controlled for age, sex, and survey year-All Subjects beyond 24 months post-infection.

|  | Infected | Course | Months since Infection | Vaccination | Months since Vaccination | Covid after Vaccination | Wuhan | Alpha | Delta | Omicron | Age | Sex |
| --- | --- | --- | --- | --- | --- | --- | --- | --- | --- | --- | --- | --- |
| Physical Health Issues | NA | **0.234** | 0.045 | -0.021 | -0.009 | **-0.218** | 0.067 | 0.048 | 0.014 | -0.101 | 0.015 | 0.057 |
| Mental Health Issues | NA | 0.047 | 0.028 | 0.019 | 0.016 | **-0.173** | -0.018 | 0.071 | -0.001 | -0.062 | 0.023 | 0.069 |
| Fatigue | NA | 0.097 | 0.041 | 0.106 | -0.003 | -0.13 | 0.12 | -0.044 | -0.095 | -0.016 | **0.13** | **0.248** |
| Intelligence | NA | 0.007 | **-0.193** | **-0.172** | -0.009 | 0.183 | **-0.144** | -0.071 | 0.086 | **0.176** | -0.024 | **-0.136** |
| Memory | NA | 0.028 | -0.079 | **-0.13** | 0.002 | 0.097 | -0.109 | -0.113 | 0.079 | 0.065 | -0.043 | **0.171** |
| Reactions | NA | 0.021 | 0.091 | **-0.164** | 0.079 | -0.09 | 0.103 | -0.092 | -0.064 | -0.016 | 0.024 | 0.073 |
| Accuracy | NA | -0.034 | -0.09 | **-0.141** | -0.04 | 0.166 | -0.099 | -0.073 | 0.066 | 0.109 | -0.097 | -0.052 |
| Output Variables |  |  |  |  |  |  |  |  |  |  |  |  |
| Evolutionary Biology Score | NA | -0.002 | 0.02 | 0.025 | -0.014 | **0.19** | -0.053 | 0.069 | 0.057 | -0.034 | **-0.161** | 0.012 |
| Free-Recall Memory | NA | 0.029 | -0.071 | -0.075 | 0.042 | 0.14 | -0.089 | -0.067 | 0.031 | 0.047 | -0.028 | **0.189** |
| Recognition Memory | NA | 0.027 | -0.076 | -0.098 | 0.03 | 0.144 | -0.077 | **-0.151** | 0.074 | 0.056 | -0.042 | **0.201** |
| Simple RT Duration | NA | -0.036 | 0.109 | **-0.172** | 0.166 | 0.004 | 0.097 | -0.069 | -0.119 | 0.013 | 0.007 | 0.011 |
| Simple RT Precision | NA | 0.022 | -0.01 | -0.078 | 0.067 | **-0.245** | -0.109 | 0.056 | 0.09 | -0.017 | -0.012 | -0.09 |
| Stroop Duration | NA | 0.041 | 0.049 | -0.112 | -0.005 | -0.079 | 0.029 | -0.009 | -0.004 | -0.032 | -0.011 | 0.059 |
| Stroop Precision | NA | -0.069 | 0.03 | -0.128 | -0.014 | 0.047 | **0.165** | **-0.256** | -0.092 | 0.051 | -0.058 | -0.095 |
| Reading Time | NA | -0.013 | 0.086 | -0.059 | 0.12 | 0.031 | 0.088 | -0.077 | -0.014 | -0.08 | 0.045 | 0.091 |
| Allergy | NA | **0.182** | 0.009 | **0.16** | -0.014 | -0.098 | 0.055 | **0.175** | -0.054 | -0.079 | 0.016 | 0.037 |
| Skin Problem | NA | **0.201** | 0.037 | -0.008 | -0.138 | -0.018 | 0.004 | 0.013 | 0.04 | -0.045 | 0.012 | -0.067 |
| Digestive Problem | NA | 0.098 | -0.056 | -0.046 | -0.043 | -0.098 | -0.014 | -0.007 | -0.023 | 0.075 | 0.107 | -0.017 |
| Infection | NA | **0.137** | 0.01 | **-0.137** | -0.005 | 0.165 | 0.066 | -0.083 | 0.029 | 0.007 | -0.055 | 0.094 |
| Cardiovascular Problem | NA | -0.001 | -0.03 | 0.112 | **-0.189** | -0.175 | -0.019 | 0.055 | -0.03 | 0.032 | -0.073 | 0.049 |
| Low Blood Pressure | NA | -0.051 | 0.008 | **-0.183** | 0.023 | -0.154 | -0.032 | 0.047 | -0.018 | 0.009 | 0.041 | **0.289** |
| High Blood Pressure | NA | 0.109 | -0.108 | -0.113 | 0.012 | 0.202 | **-0.234** | 0.096 | 0.159 | 0.073 | **0.214** | 0.089 |
| Orthopedic Problem | NA | 0.009 | **0.203** | 0.1 | 0.136 | -0.176 | **0.175** | **0.145** | -0.056 | **-0.254** | 0.06 | 0.032 |
| Metabolic Problem | NA | 0.104 | -0.12 | -0.096 | -0.049 | 0.064 | -0.117 | -0.108 | **0.175** | 0.019 | 0.023 | 0.107 |
| Neurologic Problem | NA | **0.235** | -0.062 | -0.083 | -0.005 | -0.002 | -0.025 | -0.071 | 0.076 | 0.038 | 0.001 | 0.093 |
| Headache | NA | **0.15** | 0.029 | 0.128 | 0.01 | -0.077 | -0.036 | -0.013 | **0.141** | -0.114 | -0.098 | **0.133** |
| Other Pain | NA | **0.257** | 0.085 | -0.073 | 0.079 | **-0.267** | **0.133** | -0.009 | 0.002 | -0.121 | -0.084 | 0.022 |
| Recurrent Problem | NA | 0.065 | 0.043 | 0.124 | -0.022 | -0.133 | **0.157** | 0.061 | **-0.135** | -0.023 | 0.025 | 0.03 |
| Depression | NA | 0.042 | 0.044 | -0.104 | -0.033 | **-0.243** | 0.002 | 0.012 | 0.018 | -0.046 | 0.066 | 0.003 |
| Anxiety | NA | 0.111 | 0.02 | -0.071 | -0.038 | -0.073 | 0.03 | 0.029 | 0.01 | -0.032 | 0.072 | 0.098 |
| Other Psychological Problems | NA | 0.095 | -0.017 | -0.002 | 0.033 | 0.018 | 0.026 | 0.011 | -0.044 | 0.023 | -0.019 | **0.131** |
| Medication for Mental Purposes | NA | 0.026 | **-0.198** | 0.071 | -0.155 | 0.036 | -0.169 | -0.058 | 0.045 | **0.189** | 0.102 | 0.021 |
| Doctor Visits | NA | 0.062 | -0.012 | -0.106 | -0.1 | **-0.169** | 0.061 | 0.025 | -0.012 | -0.01 | 0.09 | -0.099 |
| Other Medication | NA | 0.127 | 0.033 | 0.133 | 0.037 | -0.082 | -0.023 | 0.071 | 0.04 | -0.082 | 0.067 | 0.117 |
| Antibiotic Taken Past 3 Years | NA | 0.09 | -0.003 | -0.044 | 0.012 | 0.004 | 0.006 | -0.022 | 0.082 | -0.014 | **-0.161** | 0.055 |
| Hospitalization Past 5 Years | NA | 0.097 | -0.015 | -0.088 | 0.073 | 0.038 | 0.055 | 0.009 | -0.042 | -0.017 | 0.043 | **-0.329** |
| Life Expectancy | NA | -0.005 | 0.003 | -0.007 | 0.049 | -0.142 | 0.002 | -0.009 | 0.066 | -0.062 | -0.097 | -0.116 |
| Now Feeling Physically Miserable | NA | **0.14** | -0.019 | -0.021 | -0.01 | -0.023 | -0.01 | -0.017 | -0.01 | 0.02 | -0.026 | -0.011 |
| Usually Feeling Physically Miserable | NA | 0.004 | -0.008 | 0.045 | -0.094 | **-0.24** | -0.051 | 0.095 | 0.061 | -0.092 | 0.028 | 0.125 |
| Now Feeling Mentally Miserable | NA | 0.019 | 0.048 | 0.024 | 0.028 | -0.103 | -0.016 | 0.002 | -0.009 | -0.047 | 0.026 | 0.016 |
| Usually Feeling Mentally Miserable | NA | 0.032 | 0.077 | 0.044 | -0.007 | **-0.246** | 0.018 | **0.167** | -0.006 | **-0.138** | 0.012 | 0.098 |
| Year | NA | -0.093 | **0.374** | -0.04 | **0.436** | **0.207** | **-0.395** | 0.107 | **0.181** | **0.203** | -0.018 | 0.004 |
| Course of COVID-19 | NA | NA | 0.036 | **-0.205** | 0.105 | **0.197** | 0.023 | -0.045 | **0.18** | **-0.145** | **0.136** | -0.051 |
| Months since COVID Infection | NA | 0.036 | NA | **0.144** | **0.248** | **-0.424** | **0.582** | 0.039 | **-0.182** | **-0.587** | -0.002 | -0.062 |

Significant correlations are bolded. The *p*-values that remained or turned significant after the application of the Benjamini-Hochberg correction for multiple testing (with FDR set at 0.1) are underlined.

**Table S9.**
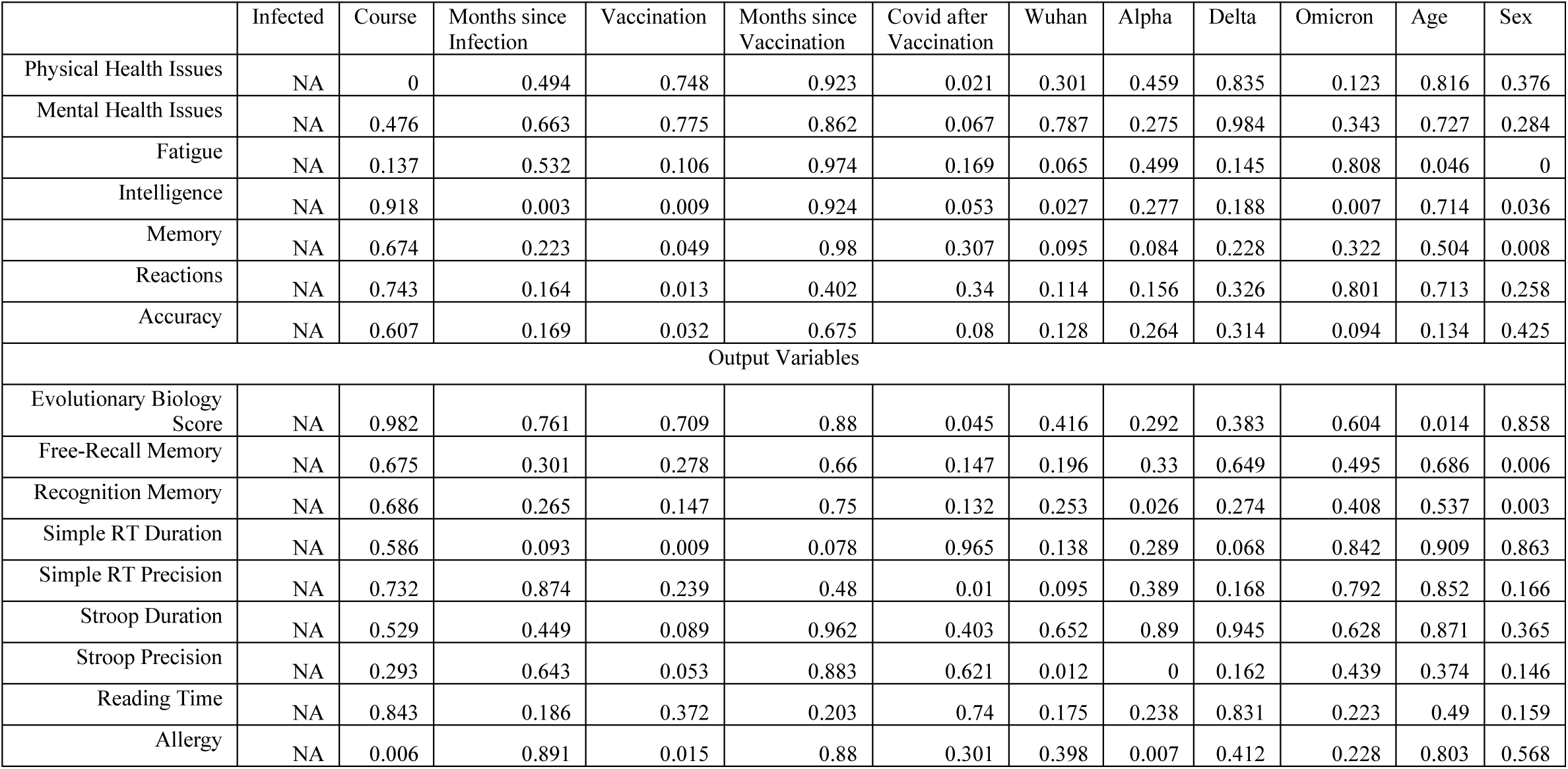
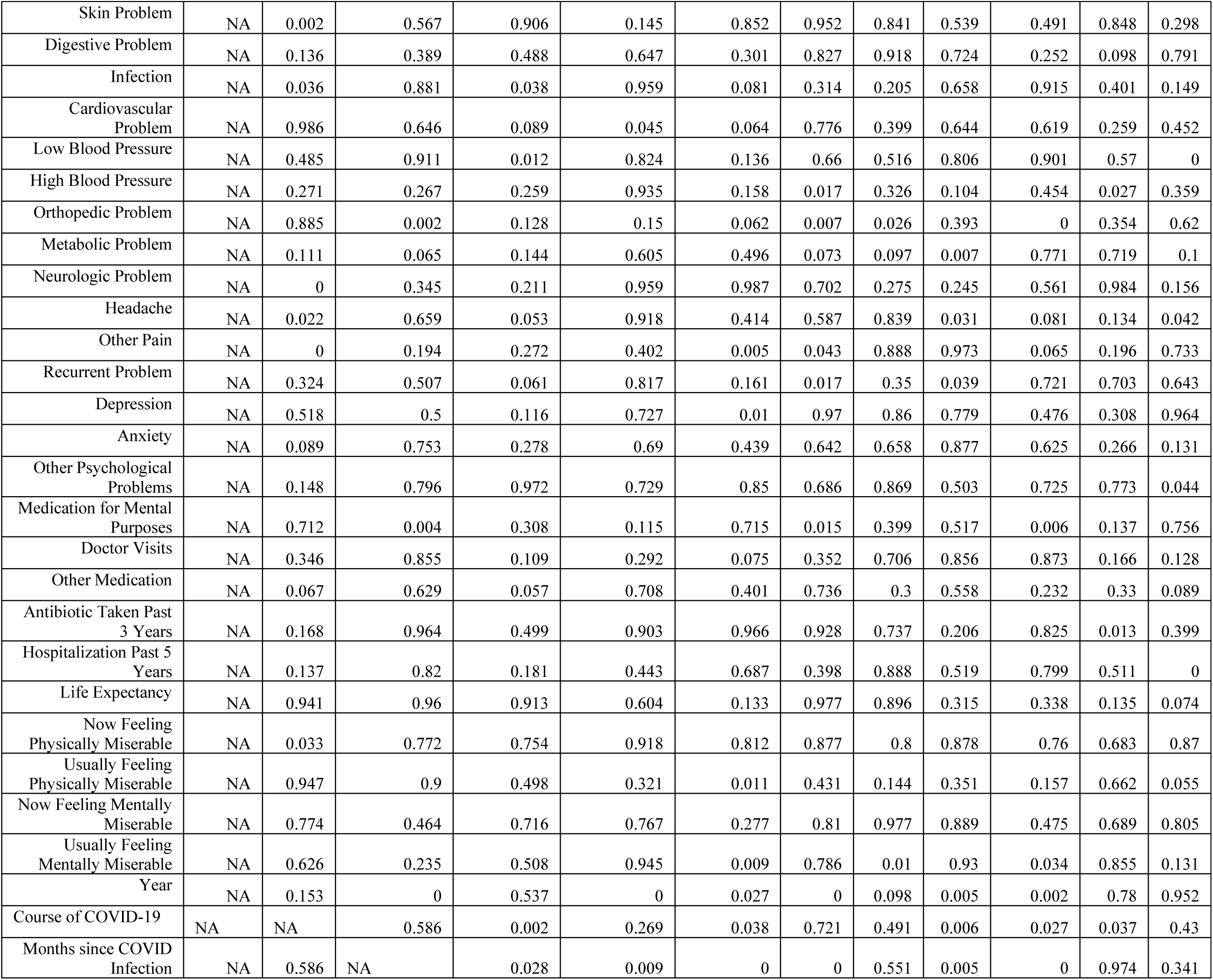
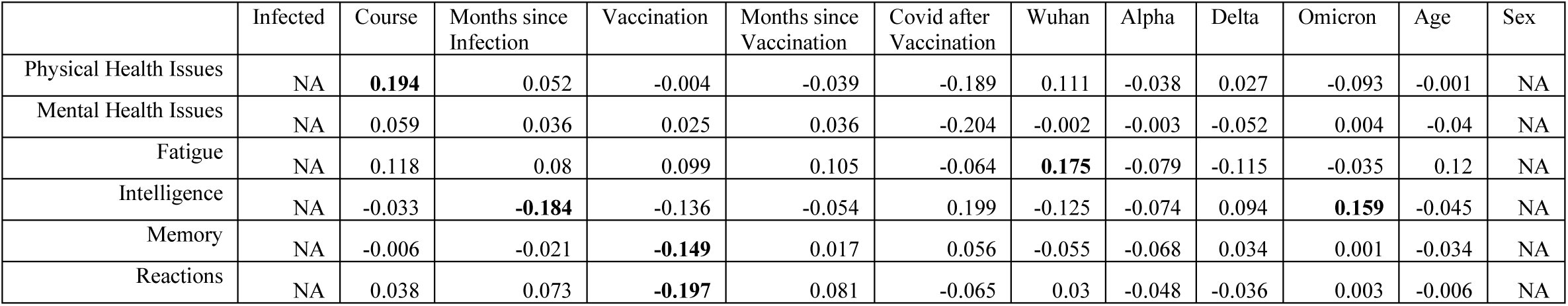
P-values of the correlations between health and performance-related variables and COVID-related variables controlled for age, sex, and survey year-All Subjects beyond 24 months post-infection.

**Table S10.**
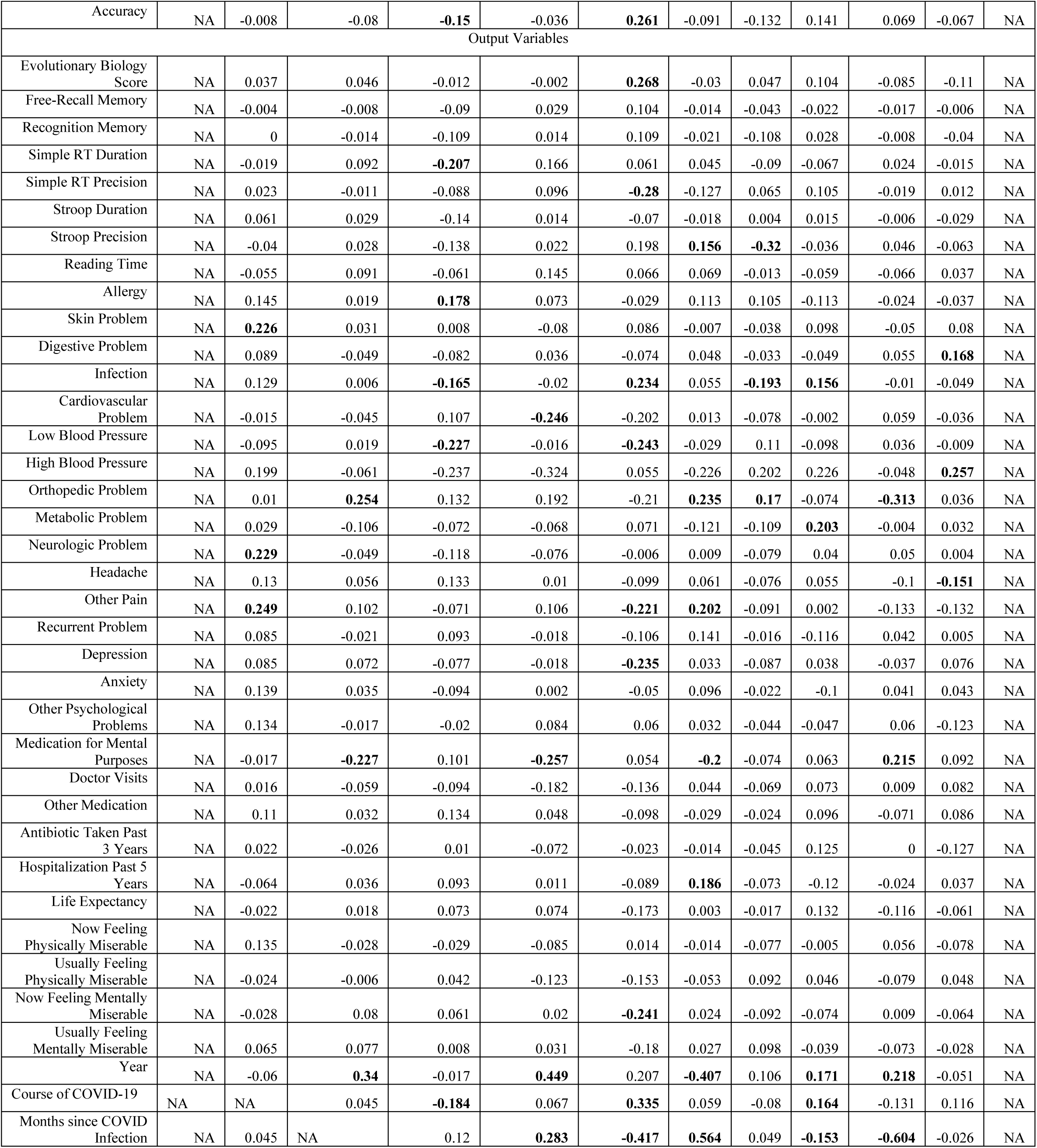
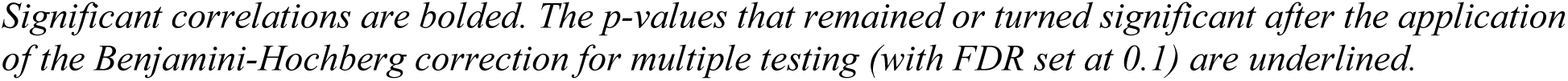
Correlations between health and performance-related variables and COVID-related variables controlled for age, sex, and survey year-Women beyond 24 months post-infection.

**Table S11.** P-values of the correlations between health and performance-related variables and COVID-related variables controlled for age, sex, and survey year-Women beyond 24 months post-infection.

|  | Infected | Course | Months since Infection | Vaccination | Months since Vaccination | Covid after Vaccination | Wuhan | Alpha | Delta | Omicron | Age | Sex |
| --- | --- | --- | --- | --- | --- | --- | --- | --- | --- | --- | --- | --- |
| Physical Health Issues | NA | 0.01 | 0.483 | 0.962 | 0.722 | 0.086 | 0.135 | 0.608 | 0.714 | 0.211 | 0.985 | NA |
| Mental Health Issues | NA | 0.428 | 0.625 | 0.738 | 0.74 | 0.064 | 0.979 | 0.971 | 0.481 | 0.955 | 0.588 | NA |
| Fatigue | NA | 0.114 | 0.281 | 0.19 | 0.341 | 0.563 | 0.018 | 0.287 | 0.122 | 0.64 | 0.104 | NA |
| Intelligence | NA | 0.663 | 0.013 | 0.071 | 0.622 | 0.071 | 0.093 | 0.322 | 0.205 | 0.033 | 0.54 | NA |
| Memory | NA | 0.932 | 0.78 | 0.048 | 0.877 | 0.611 | 0.459 | 0.356 | 0.649 | 0.995 | 0.649 | NA |
| Reactions | NA | 0.607 | 0.329 | 0.009 | 0.46 | 0.557 | 0.689 | 0.521 | 0.625 | 0.969 | 0.937 | NA |
| Accuracy | NA | 0.92 | 0.282 | 0.046 | 0.745 | 0.018 | 0.22 | 0.076 | 0.057 | 0.353 | 0.362 | NA |
| Output Variables |  |  |  |  |  |  |  |  |  |  |  |  |
| Evolutionary Biology Score | NA | 0.622 | 0.536 | 0.878 | 0.989 | 0.015 | 0.688 | 0.531 | 0.162 | 0.253 | 0.139 | NA |
| Free-Recall Memory | NA | 0.96 | 0.918 | 0.256 | 0.794 | 0.352 | 0.854 | 0.585 | 0.779 | 0.831 | 0.934 | NA |
| Recognition Memory | NA | 1 | 0.86 | 0.157 | 0.898 | 0.32 | 0.784 | 0.16 | 0.715 | 0.917 | 0.602 | NA |
| Simple RT Duration | NA | 0.796 | 0.213 | 0.006 | 0.131 | 0.578 | 0.543 | 0.225 | 0.37 | 0.749 | 0.836 | NA |
| Simple RT Precision | NA | 0.761 | 0.877 | 0.242 | 0.385 | 0.011 | 0.088 | 0.382 | 0.157 | 0.799 | 0.875 | NA |
| Stroop Duration | NA | 0.411 | 0.696 | 0.062 | 0.898 | 0.524 | 0.806 | 0.953 | 0.839 | 0.933 | 0.699 | NA |
| Stroop Precision | NA | 0.59 | 0.712 | 0.069 | 0.838 | 0.072 | 0.037 | 0 | 0.627 | 0.539 | 0.398 | NA |
| Reading Time | NA | 0.465 | 0.222 | 0.418 | 0.189 | 0.55 | 0.354 | 0.86 | 0.426 | 0.375 | 0.615 | NA |
| Allergy | NA | 0.053 | 0.797 | 0.018 | 0.509 | 0.794 | 0.129 | 0.158 | 0.127 | 0.751 | 0.611 | NA |
| Skin Problem | NA | 0.003 | 0.674 | 0.914 | 0.469 | 0.433 | 0.93 | 0.611 | 0.189 | 0.504 | 0.278 | NA |
| Digestive Problem | NA | 0.235 | 0.51 | 0.273 | 0.74 | 0.5 | 0.52 | 0.658 | 0.509 | 0.462 | 0.023 | NA |
| Infection | NA | 0.084 | 0.941 | 0.028 | 0.858 | 0.033 | 0.456 | 0.009 | 0.035 | 0.895 | 0.509 | NA |
| Cardiovascular Problem | NA | 0.844 | 0.542 | 0.154 | 0.025 | 0.067 | 0.856 | 0.294 | 0.982 | 0.429 | 0.628 | NA |
| Low Blood Pressure | NA | 0.249 | 0.816 | 0.006 | 0.89 | 0.04 | 0.721 | 0.177 | 0.231 | 0.656 | 0.915 | NA |
| High Blood Pressure | NA | 0.103 | 0.614 | 0.057 | 0.08 | 0.765 | 0.06 | 0.093 | 0.06 | 0.688 | 0.03 | NA |
| Orthopedic Problem | NA | 0.894 | 0.001 | 0.08 | 0.081 | 0.056 | 0.002 | 0.022 | 0.318 | 0 | 0.623 | NA |
| Metabolic Problem | NA | 0.697 | 0.152 | 0.337 | 0.536 | 0.517 | 0.104 | 0.143 | 0.006 | 0.959 | 0.666 | NA |
| Neurologic Problem | NA | 0.002 | 0.508 | 0.115 | 0.488 | 0.957 | 0.9 | 0.289 | 0.589 | 0.505 | 0.959 | NA |
| Headache | NA | 0.082 | 0.449 | 0.076 | 0.929 | 0.367 | 0.413 | 0.303 | 0.463 | 0.177 | 0.041 | NA |
| Other Pain | NA | 0.001 | 0.17 | 0.344 | 0.335 | 0.045 | 0.006 | 0.221 | 0.976 | 0.073 | 0.073 | NA |
| Recurrent Problem | NA | 0.257 | 0.773 | 0.215 | 0.872 | 0.335 | 0.058 | 0.831 | 0.119 | 0.569 | 0.951 | NA |
| Depression | NA | 0.258 | 0.333 | 0.308 | 0.872 | 0.033 | 0.659 | 0.242 | 0.611 | 0.615 | 0.301 | NA |
| Anxiety | NA | 0.064 | 0.638 | 0.209 | 0.983 | 0.652 | 0.197 | 0.77 | 0.179 | 0.582 | 0.564 | NA |
| Other Psychological Problems | NA | 0.074 | 0.824 | 0.791 | 0.443 | 0.587 | 0.662 | 0.557 | 0.531 | 0.422 | 0.095 | NA |
| Medication for Mental Purposes | NA | 0.834 | 0.004 | 0.211 | 0.025 | 0.637 | 0.012 | 0.35 | 0.427 | 0.007 | 0.242 | NA |
| Doctor Visits | NA | 0.83 | 0.427 | 0.211 | 0.098 | 0.218 | 0.556 | 0.353 | 0.328 | 0.899 | 0.269 | NA |
| Other Medication | NA | 0.165 | 0.681 | 0.093 | 0.672 | 0.385 | 0.716 | 0.757 | 0.224 | 0.368 | 0.27 | NA |
| Antibiotic Taken Past 3 Years | NA | 0.773 | 0.722 | 0.89 | 0.512 | 0.834 | 0.851 | 0.544 | 0.092 | 1 | 0.085 | NA |
| Hospitalization Past 5 Years | NA | 0.391 | 0.626 | 0.218 | 0.923 | 0.419 | 0.012 | 0.323 | 0.107 | 0.747 | 0.619 | NA |
| Life Expectancy | NA | 0.763 | 0.809 | 0.335 | 0.504 | 0.115 | 0.972 | 0.817 | 0.075 | 0.119 | 0.408 | NA |
| Now Feeling Physically Miserable | NA | 0.07 | 0.71 | 0.703 | 0.441 | 0.9 | 0.848 | 0.3 | 0.951 | 0.449 | 0.29 | NA |
| Usually Feeling Physically Miserable | NA | 0.749 | 0.933 | 0.58 | 0.262 | 0.164 | 0.477 | 0.217 | 0.532 | 0.286 | 0.517 | NA |
| Now Feeling Mentally Miserable | NA | 0.711 | 0.28 | 0.416 | 0.859 | 0.028 | 0.747 | 0.216 | 0.317 | 0.9 | 0.383 | NA |
| Usually Feeling Mentally Miserable | NA | 0.388 | 0.301 | 0.912 | 0.781 | 0.103 | 0.717 | 0.185 | 0.597 | 0.324 | 0.707 | NA |
| Year | NA | 0.42 | 0 | 0.822 | 0 | 0.057 | 0 | 0.153 | 0.021 | 0.003 | 0.485 | NA |
| Course of COVID-19 | NA | NA | 0.549 | 0.014 | 0.543 | 0.002 | 0.43 | 0.283 | 0.029 | 0.079 | 0.118 | NA |
| Months since COVID Infection | NA | 0.549 | NA | 0.111 | 0.01 | 0 | 0 | 0.509 | 0.039 | 0 | 0.725 | NA |

**Table S12.**
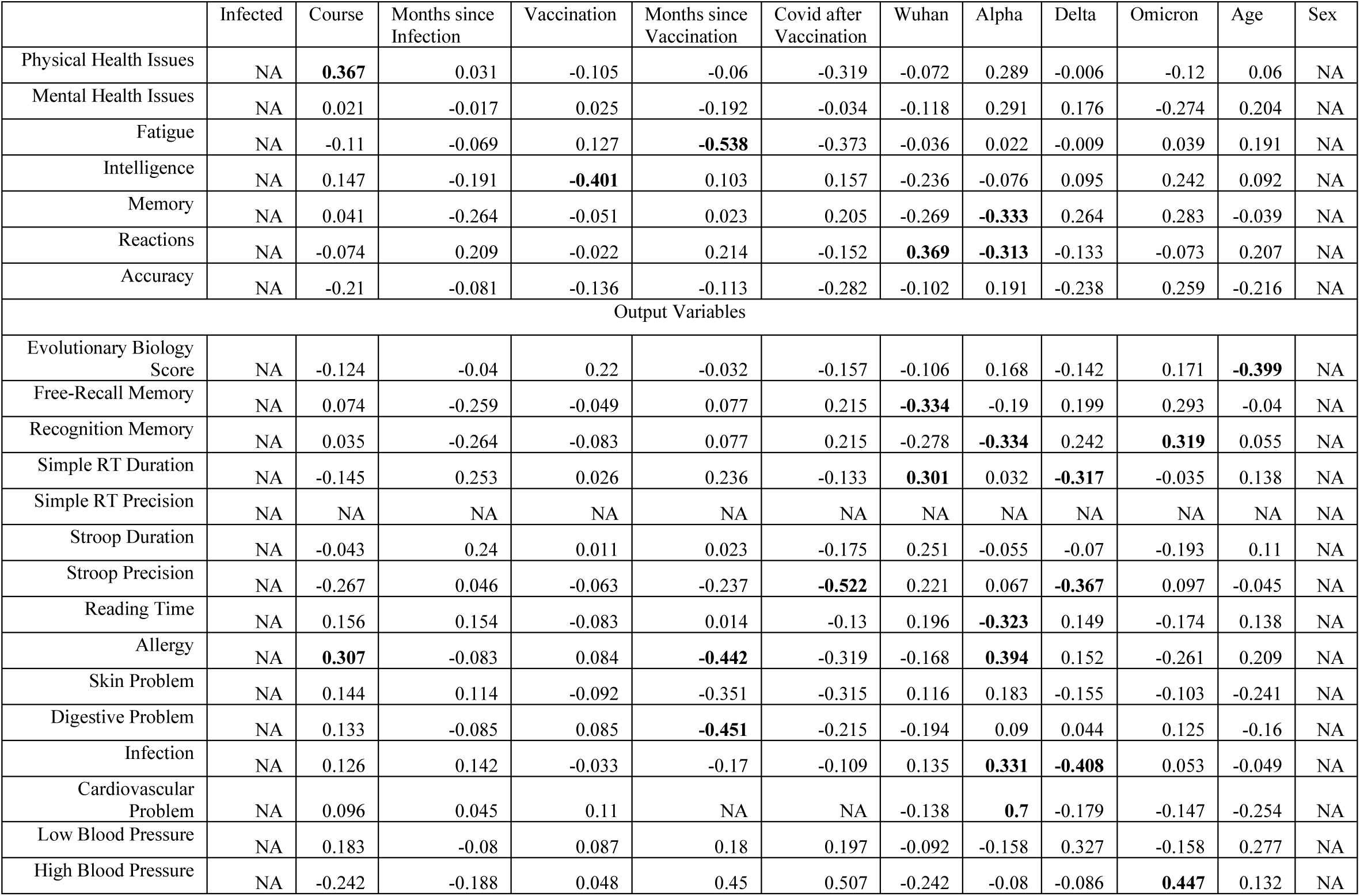
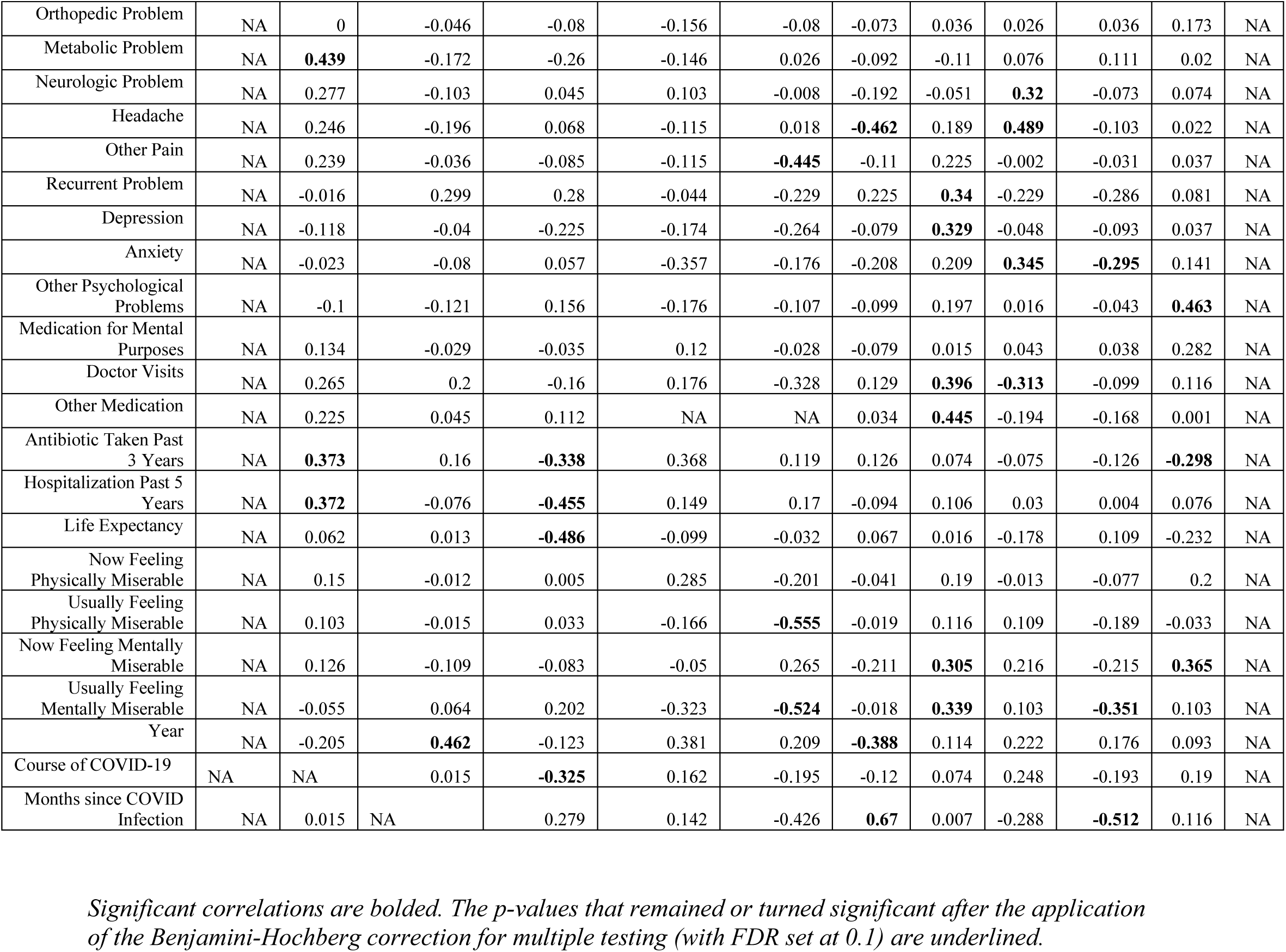
Correlations between health and performance-related variables and COVID-related variables controlled for age, sex, and survey year-Men beyond 24 months post-infection.

**Table S13.** P-values of the correlations between health and performance-related variables and COVID-related variables controlled for age, sex, and survey year-Men beyond 24 months post-infection.

|  | Infected | Course | Months since Infection | Vaccination | Months since Vaccination | Covid after Vaccination | Wuhan | Alpha | Delta | Omicron | Age | Sex |
| --- | --- | --- | --- | --- | --- | --- | --- | --- | --- | --- | --- | --- |
| Physical Health Issues | NA | 0.014 | 0.835 | 0.483 | 0.787 | 0.149 | 0.63 | 0.054 | 0.966 | 0.424 | 0.683 | NA |
| Mental Health Issues | NA | 0.889 | 0.911 | 0.869 | 0.385 | 0.877 | 0.429 | 0.052 | 0.238 | 0.067 | 0.162 | NA |
| Fatigue | NA | 0.463 | 0.647 | 0.395 | 0.015 | 0.092 | 0.812 | 0.881 | 0.95 | 0.797 | 0.19 | NA |
| Intelligence | NA | 0.325 | 0.201 | 0.007 | 0.642 | 0.477 | 0.115 | 0.612 | 0.524 | 0.105 | 0.529 | NA |
| Memory | NA | 0.784 | 0.078 | 0.734 | 0.916 | 0.353 | 0.072 | 0.026 | 0.078 | 0.058 | 0.792 | NA |
| Reactions | NA | 0.619 | 0.162 | 0.882 | 0.332 | 0.493 | 0.014 | 0.037 | 0.374 | 0.626 | 0.157 | NA |
| Accuracy | NA | 0.16 | 0.59 | 0.363 | 0.608 | 0.201 | 0.496 | 0.202 | 0.112 | 0.084 | 0.139 | NA |
| Output Variables |  |  |  |  |  |  |  |  |  |  |  |  |
| Evolutionary Biology Score | NA | 0.406 | 0.79 | 0.142 | 0.884 | 0.476 | 0.478 | 0.262 | 0.342 | 0.254 | 0.006 | NA |
| Free-Recall Memory | NA | 0.638 | 0.101 | 0.757 | 0.742 | 0.358 | 0.034 | 0.228 | 0.206 | 0.063 | 0.795 | NA |
| Recognition Memory | NA | 0.822 | 0.094 | 0.599 | 0.742 | 0.358 | 0.078 | 0.034 | 0.124 | 0.043 | 0.718 | NA |
| Simple RT Duration | NA | 0.331 | 0.091 | 0.861 | 0.286 | 0.548 | 0.044 | 0.833 | 0.034 | 0.816 | 0.346 | NA |
| Simple RT Precision | NA | NA | NA | NA | NA | NA | NA | NA | NA | NA | NA | NA |
| Stroop Duration | NA | 0.772 | 0.109 | 0.941 | 0.917 | 0.429 | 0.093 | 0.713 | 0.642 | 0.197 | 0.453 | NA |
| Stroop Precision | NA | 0.074 | 0.76 | 0.672 | 0.284 | 0.018 | 0.14 | 0.656 | 0.014 | 0.515 | 0.758 | NA |
| Reading Time | NA | 0.297 | 0.304 | 0.58 | 0.948 | 0.555 | 0.191 | 0.031 | 0.321 | 0.245 | 0.345 | NA |
| Allergy | NA | 0.04 | 0.578 | 0.574 | 0.045 | 0.149 | 0.262 | 0.009 | 0.311 | 0.081 | 0.153 | NA |
| Skin Problem | NA | 0.337 | 0.448 | 0.537 | 0.112 | 0.154 | 0.439 | 0.221 | 0.299 | 0.49 | 0.099 | NA |
| Digestive Problem | NA | 0.373 | 0.569 | 0.57 | 0.041 | 0.329 | 0.194 | 0.549 | 0.768 | 0.404 | 0.274 | NA |
| Infection | NA | 0.398 | 0.344 | 0.824 | 0.443 | 0.621 | 0.367 | 0.027 | 0.006 | 0.724 | 0.735 | NA |
| Cardiovascular Problem | NA | 0.522 | 0.765 | 0.463 | NA | NA | 0.357 | 0 | 0.232 | 0.326 | 0.082 | NA |
| Low Blood Pressure | NA | 0.288 | 0.641 | 0.614 | 0.5 | 0.459 | 0.593 | 0.359 | 0.058 | 0.359 | 0.097 | NA |
| High Blood Pressure | NA | 0.209 | 0.33 | 0.802 | 0.119 | 0.079 | 0.208 | 0.677 | 0.656 | 0.02 | 0.475 | NA |
| Orthopedic Problem | NA | 1 | 0.757 | 0.591 | 0.479 | 0.718 | 0.624 | 0.812 | 0.864 | 0.812 | 0.237 | NA |
| Metabolic Problem | NA | 0.003 | 0.251 | 0.083 | 0.509 | 0.907 | 0.539 | 0.461 | 0.613 | 0.459 | 0.889 | NA |
| Neurologic Problem | NA | 0.071 | 0.503 | 0.769 | 0.64 | 0.973 | 0.212 | 0.738 | 0.037 | 0.636 | 0.622 | NA |
| Headache | NA | 0.109 | 0.203 | 0.658 | 0.602 | 0.936 | 0.003 | 0.218 | 0.001 | 0.501 | 0.885 | NA |
| Other Pain | NA | 0.12 | 0.814 | 0.581 | 0.603 | 0.044 | 0.473 | 0.143 | 0.99 | 0.84 | 0.806 | NA |
| Recurrent Problem | NA | 0.915 | 0.051 | 0.069 | 0.842 | 0.299 | 0.143 | 0.027 | 0.135 | 0.063 | 0.59 | NA |
| Depression | NA | 0.432 | 0.792 | 0.133 | 0.431 | 0.233 | 0.597 | 0.028 | 0.749 | 0.535 | 0.798 | NA |
| Anxiety | NA | 0.877 | 0.592 | 0.706 | 0.106 | 0.426 | 0.165 | 0.163 | 0.021 | 0.049 | 0.333 | NA |
| Other Psychological Problems | NA | 0.515 | 0.429 | 0.311 | 0.426 | 0.627 | 0.517 | 0.2 | 0.919 | 0.779 | 0.002 | NA |
| Medication for Mental Purposes | NA | 0.396 | 0.852 | 0.826 | 0.607 | 0.903 | 0.618 | 0.922 | 0.787 | 0.811 | 0.066 | NA |
| Doctor Visits | NA | 0.077 | 0.181 | 0.285 | 0.426 | 0.138 | 0.388 | 0.008 | 0.037 | 0.51 | 0.428 | NA |
| Other Medication | NA | 0.153 | 0.776 | 0.479 | NA | NA | 0.827 | 0.005 | 0.218 | 0.287 | 0.994 | NA |
| Antibiotic Taken Past 3 Years | NA | 0.013 | 0.286 | 0.024 | 0.096 | 0.591 | 0.401 | 0.623 | 0.615 | 0.401 | 0.041 | NA |
| Hospitalization Past 5 Years | NA | 0.013 | 0.613 | 0.002 | 0.501 | 0.442 | 0.529 | 0.479 | 0.841 | 0.98 | 0.603 | NA |
| Life Expectancy | NA | 0.679 | 0.929 | 0.001 | 0.656 | 0.883 | 0.653 | 0.913 | 0.234 | 0.466 | 0.112 | NA |
| Now Feeling Physically Miserable | NA | 0.315 | 0.938 | 0.971 | 0.197 | 0.362 | 0.784 | 0.204 | 0.931 | 0.607 | 0.171 | NA |
| Usually Feeling Physically Miserable | NA | 0.493 | 0.922 | 0.823 | 0.453 | 0.012 | 0.897 | 0.44 | 0.465 | 0.206 | 0.82 | NA |
| Now Feeling Mentally Miserable | NA | 0.399 | 0.467 | 0.578 | 0.822 | 0.23 | 0.159 | 0.041 | 0.149 | 0.151 | 0.012 | NA |
| Usually Feeling Mentally Miserable | NA | 0.712 | 0.667 | 0.178 | 0.143 | 0.018 | 0.906 | 0.024 | 0.493 | 0.019 | 0.48 | NA |
| Year | NA | 0.161 | 0.002 | 0.401 | 0.07 | 0.32 | 0.008 | 0.436 | 0.129 | 0.229 | 0.513 | NA |
| Course of COVID-19 | NA | NA | 0.922 | 0.03 | 0.463 | 0.378 | 0.424 | 0.62 | 0.097 | 0.198 | 0.193 | NA |
| Months since COVID Infection | NA | 0.922 | NA | 0.062 | 0.52 | 0.054 | 0 | 0.964 | 0.054 | 0.001 | 0.426 | NA |

*Significant correlations are bolded. The p-values that remained or turned significant after the application of the Benjamini-Hochberg correction for multiple testing (with FDR set at 0.1) are underlined.*

**Table S14.** Correlations between health and performance-related variables and COVID-related variables controlled for age, sex, and survey year-all subjects with at least 36 months elapsed since infection.

|  | Infected | Course | Months since Infection | Vaccination | Months since Vaccination | Covid after Vaccination | Wuhan | Alpha | Delta | Omicron | Age | Sex |
| --- | --- | --- | --- | --- | --- | --- | --- | --- | --- | --- | --- | --- |
| Physical Health Issues | NA | **0.329** | -0.028 | -0.081 | 0.088 | NA | 0.041 | 0.016 | -0.045 | NA | -0.101 | 0 |
| Mental Health Issues | NA | 0.088 | 0.037 | -0.086 | 0.019 | NA | -0.126 | 0.086 | -0.061 | NA | -0.033 | 0.039 |
| Fatigue | NA | 0.073 | 0.009 | 0.192 | -0.184 | NA | 0.144 | -0.148 | 0.017 | NA | 0.085 | **0.393** |
| Intelligence | NA | -0.041 | -0.072 | -0.192 | 0.024 | NA | 0.106 | -0.013 | -0.012 | NA | -0.038 | -0.157 |
| Memory | NA | -0.125 | 0.038 | **-0.218** | -0.031 | NA | -0.055 | -0.136 | 0.168 | NA | -0.022 | **0.318** |
| Reactions | NA | 0.093 | 0.093 | **-0.233** | 0.011 | NA | 0.108 | -0.18 | 0.035 | NA | -0.027 | 0.038 |
| Accuracy | NA | 0.04 | 0.125 | **-0.217** | 0.027 | NA | 0.085 | -0.035 | -0.142 | NA | -0.162 | -0.153 |
| Evolutionary Biology Score | NA | 0.098 | 0.015 | -0.023 | 0.077 | NA | -0.045 | 0.107 | -0.128 | NA | **-0.288** | -0.03 |
| Free-Recall Memory | NA | -0.098 | -0.025 | -0.179 | 0.007 | NA | -0.107 | -0.12 | 0.2 | NA | -0.009 | **0.423** |
| Recognition Memory | NA | -0.111 | 0.032 | -0.184 | -0.018 | NA | 0 | **-0.217** | 0.193 | NA | 0.009 | **0.421** |
| Simple RT Duration | NA | 0.143 | **0.235** | **-0.246** | 0.211 | NA | 0.11 | -0.141 | -0.17 | NA | 0.005 | -0.06 |
| Simple RT Precision | NA | -0.06 | 0.084 | -0.068 | NA | NA | -0.134 | 0.097 | 0.038 | NA | 0.009 | -0.107 |
| Stroop Duration | NA | -0.012 | -0.053 | -0.003 | -0.005 | NA | -0.019 | -0.044 | 0.14 | NA | -0.087 | -0.01 |
| Stroop Precision | NA | 0.039 | **0.341** | -0.112 | 0.066 | NA | **0.391** | **-0.369** | **-0.389** | NA | -0.014 | -0.184 |
| Reading Time | NA | -0.044 | 0.104 | -0.033 | 0.051 | NA | 0.041 | -0.134 | 0.033 | NA | 0.072 | 0.186 |
| Allergy | NA | **0.216** | **-0.251** | 0.02 | -0.122 | NA | -0.089 | **0.205** | 0.062 | NA | -0.045 | 0.035 |
| Skin Problem | NA | 0.17 | 0.013 | -0.107 | -0.065 | NA | -0.015 | -0.002 | 0.104 | NA | -0.155 | -0.159 |
| Digestive Problem | NA | 0.041 | -0.03 | -0.004 | 0.125 | NA | 0.068 | 0.011 | -0.018 | NA | 0.099 | 0.055 |
| Infection | NA | **0.197** | 0.085 | -0.187 | -0.176 | NA | 0.155 | -0.107 | 0.105 | NA | -0.022 | -0.013 |
| Cardiovascular Problem | NA | -0.001 | 0.032 | 0.067 | -0.262 | NA | -0.031 | 0.089 | -0.037 | NA | -0.171 | -0.092 |
| Low Blood Pressure | NA | -0.126 | 0.014 | -0.196 | 0.281 | NA | 0.004 | 0.064 | -0.21 | NA | -0.2 | **0.329** |
| High Blood Pressure | NA | -0.149 | -0.018 | NA | -0.2 | NA | -0.164 | **0.304** | -0.113 | NA | -0.043 | 0.234 |
| Orthopedic Problem | NA | -0.004 | 0.016 | 0 | 0.045 | NA | 0.046 | 0.052 | **-0.203** | NA | 0.037 | 0.171 |
| Metabolic Problem | NA | **0.267** | -0.01 | -0.071 | 0.289 | NA | 0.032 | -0.081 | -0.071 | NA | 0.122 | 0.151 |
| Neurologic Problem | NA | **0.224** | 0.098 | -0.099 | -0.056 | NA | 0.177 | -0.08 | -0.083 | NA | -0.068 | **0.216** |
| Headache | NA | 0.147 | 0.092 | 0.093 | 0.016 | NA | -0.047 | -0.04 | 0.063 | NA | **-0.22** | **0.262** |
| Other Pain | NA | 0.11 | 0.03 | **-0.228** | 0.116 | NA | 0.167 | -0.089 | -0.066 | NA | **-0.255** | 0.006 |
| Recurrent Problem | NA | 0.016 | -0.05 | 0.058 | -0.045 | NA | 0.108 | 0.045 | -0.009 | NA | -0.115 | -0.124 |
| Depression | NA | 0.086 | 0.13 | 0.026 | -0.01 | NA | -0.045 | 0.003 | 0.013 | NA | 0.024 | -0.032 |
| Anxiety | NA | 0.07 | 0.044 | -0.073 | -0.049 | NA | 0.061 | 0.035 | -0.039 | NA | -0.018 | 0.131 |
| Other Psychological Problems | NA | 0.193 | -0.075 | -0.124 | 0.101 | NA | 0.02 | -0.001 | 0.058 | NA | -0.053 | 0.067 |
| Medication for Mental Purposes | NA | NA | NA | NA | NA | NA | NA | NA | NA | NA | NA | NA |
| Doctor Visits | NA | 0.157 | -0.107 | -0.123 | -0.221 | NA | 0.054 | 0.008 | **0.217** | NA | 0.006 | **-0.252** |
| Other Medication | NA | 0.198 | -0.04 | 0.135 | 0.098 | NA | -0.108 | 0.082 | -0.075 | NA | -0.025 | 0.005 |
| Antibiotic Taken<br>Past 3 Years | NA | 0.151 | -0.034 | **0.196** | -0.073 | NA | 0.055 | -0.021 | **0.211** | NA | -0.185 | -0.008 |
| Hospitalization<br>Past 5 Years | NA | 0.032 | -0.119 | 0.063 | -0.148 | NA | 0.061 | -0.019 | -0.035 | NA | -0.062 | **-0.248** |
| Life Expectancy | NA | 0.036 | -0.001 | 0.007 | 0.022 | NA | -0.006 | -0.043 | 0.14 | NA | **-0.261** | -0.128 |
| Now Feeling<br>Physically<br>Miserable | NA | **0.214** | 0.059 | -0.192 | 0.102 | NA | 0.027 | -0.021 | -0.123 | NA | -0.12 | -0.077 |
| Usually Feeling<br>Physically<br>Miserable | NA | 0.081 | -0.065 | 0.022 | 0.059 | NA | -0.135 | 0.124 | -0.076 | NA | -0.046 | 0.144 |
| Now Feeling<br>Mentally<br>Miserable | NA | 0.087 | 0.105 | -0.156 | 0.076 | NA | -0.093 | -0.025 | -0.08 | NA | 0.044 | 0.007 |
| Usually Feeling<br>Mentally<br>Miserable | NA | 0.083 | -0.012 | -0.024 | -0.041 | NA | -0.158 | **0.194** | -0.086 | NA | -0.031 | 0.022 |
| Year | NA | NA | NA | NA | NA | NA | NA | NA | NA | NA | NA | NA |
| Course of COVID-19 | NA | NA | 0.036 | -0.088 | 0.041 | NA | 0.135 | -0.055 | -0.127 | NA | 0.112 | -0.036 |
| Months since<br>COVID Infection | NA | 0.036 | NA | -0.019 | 0.158 | NA | **0.342** | **-0.526** | **-0.198** | NA | -0.039 | -0.042 |

*Significant correlations are bolded. The p-values that remained or turned significant after the application of the Benjamini-Hochberg correction for multiple testing (with FDR set at 0.1) are underlined.*

**Table S15.** P-values of the correlations between health and performance-related variables and COVID-related variables controlled for age, sex, and survey year-all subjects with at least 36 months elapsed since infection.

|  | Infected | Course | Months since<br>Infection | Vaccination | Months since<br>Vaccination | Covid after<br>Vaccination | Wuhan | Alpha | Delta | Omicron | Age | Sex |
| --- | --- | --- | --- | --- | --- | --- | --- | --- | --- | --- | --- | --- |
| Physical Health<br>Issues | NA | 0.001 | 0.778 | 0.413 | 0.556 | NA | 0.679 | 0.873 | 0.648 | NA | 0.303 | 0.998 |
| Mental Health<br>Issues | NA | 0.372 | 0.705 | 0.384 | 0.898 | NA | 0.201 | 0.384 | 0.54 | NA | 0.734 | 0.689 |
| Fatigue | NA | 0.459 | 0.929 | 0.052 | 0.22 | NA | 0.146 | 0.132 | 0.867 | NA | 0.384 | 0 |
| Intelligence | NA | 0.677 | 0.467 | 0.051 | 0.874 | NA | 0.283 | 0.893 | 0.904 | NA | 0.693 | 0.108 |
| Memory | NA | 0.206 | 0.698 | 0.027 | 0.836 | NA | 0.577 | 0.168 | 0.088 | NA | 0.824 | 0.001 |
| Reactions | NA | 0.346 | 0.348 | 0.018 | 0.94 | NA | 0.273 | 0.068 | 0.719 | NA | 0.785 | 0.698 |
| Accuracy | NA | 0.683 | 0.205 | 0.028 | 0.858 | NA | 0.39 | 0.72 | 0.15 | NA | 0.097 | 0.117 |
| Evolutionary<br>Biology Score | NA | 0.325 | 0.878 | 0.819 | 0.607 | NA | 0.654 | 0.282 | 0.198 | NA | 0.003 | 0.761 |
| Free-Recall<br>Memory | NA | 0.352 | 0.81 | 0.092 | 0.962 | NA | 0.312 | 0.256 | 0.058 | NA | 0.934 | 0 |
| Recognition<br>Memory | NA | 0.277 | 0.755 | 0.071 | 0.905 | NA | 0.997 | 0.033 | 0.058 | NA | 0.929 | 0 |
| Simple RT<br>Duration | NA | 0.149 | 0.017 | 0.013 | 0.158 | NA | 0.264 | 0.154 | 0.085 | NA | 0.958 | 0.539 |
| Simple RT<br>Precision | NA | 0.545 | 0.392 | 0.494 | 25 | NA | 0.175 | 0.327 | 0.701 | NA | 0.925 | 0.271 |
| Stroop Duration | NA | 0.905 | 0.592 | 0.978 | 0.973 | NA | 0.848 | 0.657 | 0.156 | NA | 0.371 | 0.92 |
| Stroop Precision | NA | 0.696 | 0.001 | 0.263 | 0.661 | NA | 0 | 0 | 0 | NA | 0.884 | 0.062 |
| Reading Time | NA | 0.659 | 0.294 | 0.738 | 0.735 | NA | 0.675 | 0.176 | 0.736 | NA | 0.463 | 0.057 |
| Allergy | NA | 0.028 | 0.011 | 0.84 | 0.414 | NA | 0.369 | 0.037 | 0.529 | NA | 0.646 | 0.717 |
| Skin Problem | NA | 0.084 | 0.898 | 0.28 | 0.664 | NA | 0.878 | 0.986 | 0.29 | NA | 0.112 | 0.103 |
| Digestive Problem | NA | 0.676 | 0.763 | 0.967 | 0.403 | NA | 0.493 | 0.91 | 0.855 | NA | 0.31 | 0.571 |
| Infection | NA | 0.046 | 0.389 | 0.058 | 0.24 | NA | 0.115 | 0.277 | 0.288 | NA | 0.824 | 0.896 |
| Cardiovascular Problem | NA | 0.988 | 0.749 | 0.495 | 0.081 | NA | 0.751 | 0.369 | 0.706 | NA | 0.08 | 0.347 |
| Low Blood Pressure | NA | 0.241 | 0.895 | 0.067 | 0.083 | NA | 0.973 | 0.553 | 0.05 | NA | 0.059 | 0.002 |
| High Blood Pressure | NA | 0.331 | 0.908 | 2 | 0.453 | NA | 0.286 | 0.047 | 0.462 | NA | 0.772 | 0.118 |
| Orthopedic Problem | NA | 0.97 | 0.869 | 0.998 | 0.766 | NA | 0.641 | 0.601 | 0.039 | NA | 0.701 | 0.08 |
| Metabolic Problem | NA | 0.007 | 0.915 | 0.474 | 0.053 | NA | 0.748 | 0.412 | 0.473 | NA | 0.212 | 0.122 |
| Neurologic Problem | NA | 0.023 | 0.318 | 0.317 | 0.707 | NA | 0.073 | 0.417 | 0.402 | NA | 0.489 | 0.027 |
| Headache | NA | 0.136 | 0.349 | 0.346 | 0.916 | NA | 0.634 | 0.684 | 0.523 | NA | 0.024 | 0.007 |
| Other Pain | NA | 0.266 | 0.759 | 0.021 | 0.44 | NA | 0.091 | 0.364 | 0.505 | NA | 0.009 | 0.947 |
| Recurrent Problem | NA | 0.875 | 0.615 | 0.557 | 0.762 | NA | 0.272 | 0.651 | 0.924 | NA | 0.238 | 0.204 |
| Depression | NA | 0.386 | 0.187 | 0.791 | 0.947 | NA | 0.651 | 0.976 | 0.895 | NA | 0.802 | 0.742 |
| Anxiety | NA | 0.481 | 0.659 | 0.46 | 0.743 | NA | 0.534 | 0.723 | 0.69 | NA | 0.851 | 0.179 |
| Other Psychological Problems | NA | 0.053 | 0.455 | 0.215 | 0.499 | NA | 0.84 | 0.992 | 0.562 | NA | 0.589 | 0.499 |
| Medication for Mental Purposes | NA | NA | NA | NA | NA | NA | NA | NA | NA | NA | NA | NA |
| Doctor Visits | NA | 0.112 | 0.28 | 0.213 | 0.14 | NA | 0.585 | 0.933 | 0.028 | NA | 0.952 | 0.01 |
| Other Medication | NA | 0.062 | 0.705 | 0.201 | 0.525 | NA | 0.308 | 0.439 | 0.476 | NA | 0.808 | 0.963 |
| Antibiotic Taken Past 3 Years | NA | 0.126 | 0.727 | 0.047 | 0.628 | NA | 0.577 | 0.832 | 0.033 | NA | 0.058 | 0.932 |
| Hospitalization Past 5 Years | NA | 0.744 | 0.229 | 0.523 | 0.323 | NA | 0.534 | 0.85 | 0.722 | NA | 0.523 | 0.011 |
| Life Expectancy | NA | 0.717 | 0.991 | 0.945 | 0.883 | NA | 0.952 | 0.664 | 0.155 | NA | 0.007 | 0.189 |
| Now Feeling Physically Miserable | NA | 0.03 | 0.549 | 0.051 | 0.496 | NA | 0.786 | 0.834 | 0.211 | NA | 0.22 | 0.431 |
| Usually Feeling Physically Miserable | NA | 0.414 | 0.512 | 0.821 | 0.693 | NA | 0.17 | 0.208 | 0.439 | NA | 0.637 | 0.14 |
| Now Feeling Mentally Miserable | NA | 0.379 | 0.289 | 0.113 | 0.611 | NA | 0.348 | 0.796 | 0.415 | NA | 0.655 | 0.939 |
| Usually Feeling Mentally Miserable | NA | 0.402 | 0.903 | 0.808 | 0.783 | NA | 0.108 | 0.049 | 0.383 | NA | 0.75 | 0.824 |
| Year | NA | NA | NA | NA | NA | NA | NA | NA | NA | NA | NA | NA |
| Course of COVID-19 | NA | NA | 0.716 | 0.373 | 0.786 | NA | 0.171 | 0.577 | 0.196 | NA | 0.252 | 0.709 |
| Months since COVID Infection | NA | 0.716 | NA | 0.848 | 0.29 | NA | 0.001 | 0 | 0.045 | NA | 0.688 | 0.666 |

*Significant correlations are bolded. The p-values that remained or turned significant after the application of the Benjamini-Hochberg correction for multiple testing (with FDR set at 0.1) are underlined.*

**Table S16.** Correlations between health and performance-related variables and COVID-related variables controlled for age, sex, and survey year-Women with at least 36 months elapsed since infection.

|  | Infected | Course | Months since Infection | Vaccination | Months since Vaccination | Covid after Vaccination | Wuhan | Alpha | Delta | Omicron | Age | Sex |
| --- | --- | --- | --- | --- | --- | --- | --- | --- | --- | --- | --- | --- |
| Physical Health Issues | NA | **0.373** | 0.077 | -0.1 | 0.072 | NA | 0.197 | -0.135 | -0.064 | NA | -0.094 | NA |
| Mental Health Issues | NA | 0.124 | 0.213 | -0.087 | 0.213 | NA | -0.047 | -0.02 | -0.087 | NA | -0.073 | NA |
| Fatigue | NA | 0.143 | 0.083 | **0.228** | -0.107 | NA | 0.204 | **-0.23** | 0.01 | NA | 0.062 | NA |
| Intelligence | NA | -0.103 | -0.079 | -0.205 | -0.058 | NA | 0.15 | -0.036 | -0.013 | NA | -0.158 | NA |
| Memory | NA | -0.149 | 0.038 | **-0.244** | -0.084 | NA | -0.129 | -0.108 | <u>0.202</u> | NA | -0.117 | NA |
| Reactions | NA | 0.125 | 0.054 | **-0.264** | 0.013 | NA | 0.027 | -0.104 | 0.044 | NA | -0.085 | NA |
| Accuracy | NA | 0.032 | 0.144 | -0.221 | -0.001 | NA | 0.167 | -0.118 | -0.153 | NA | -0.186 | NA |
| Evolutionary Biology Score | NA | 0.137 | 0.039 | -0.028 | 0.09 | NA | 0.001 | 0.073 | -0.15 | NA | -0.24 | NA |
| Free-Recall Memory | NA | -0.115 | -0.009 | -0.191 | -0.04 | NA | -0.166 | -0.123 | 0.253 | NA | -0.112 | NA |
| Recognition Memory | NA | -0.113 | 0.025 | -0.2 | -0.076 | NA | -0.092 | -0.19 | **0.245** | NA | -0.111 | NA |
| Simple RT Duration | NA | 0.161 | **0.271** | **-0.261** | 0.226 | NA | 0.102 | -0.134 | **-0.203** | NA | -0.012 | NA |
| Simple RT Precision | NA | -0.066 | 0.098 | -0.067 | NA | NA | -0.146 | 0.109 | 0.038 | NA | 0.005 | NA |
| Stroop Duration | NA | 0.043 | -0.077 | 0.001 | 0.054 | NA | -0.055 | -0.012 | 0.152 | NA | -0.112 | NA |
| Stroop Precision | NA | 0.036 | **0.386** | -0.111 | 0.074 | NA | **0.427** | **-0.418** | **-0.379** | NA | -0.023 | NA |
| Reading Time | NA | 0.012 | 0.068 | -0.033 | 0.103 | NA | -0.04 | -0.049 | 0.02 | NA | 0.055 | NA |
| Allergy | NA | 0.204 | -0.114 | 0.028 | 0.005 | NA | 0.038 | 0.101 | 0.07 | NA | -0.105 | NA |
| Skin Problem | NA | **0.242** | 0.054 | -0.13 | 0.129 | NA | 0.025 | -0.058 | 0.124 | NA | -0.05 | NA |
| Digestive Problem | NA | 0.051 | -0.1 | -0.018 | 0.266 | NA | 0.121 | -0.042 | -0.012 | NA | **0.227** | NA |
| Infection | NA | **0.228** | 0.156 | -0.206 | -0.173 | NA | **0.264** | **-0.223** | 0.115 | NA | -0.002 | NA |
| Cardiovascular Problem | NA | -0.051 | 0.167 | 0.072 | -0.332 | NA | 0.154 | -0.113 | -0.04 | NA | -0.062 | NA |
| Low Blood Pressure | NA | -0.184 | 0.071 | -0.24 | 0.223 | NA | -0.047 | 0.169 | **-0.249** | NA | **-0.367** | NA |
| High Blood Pressure | NA | -0.179 | -0.021 | NA | -0.4 | NA | -0.204 | **0.423** | -0.117 | NA | -0.042 | NA |
| Orthopedic Problem | NA | -0.015 | 0.01 | 0.003 | 0.069 | NA | 0.073 | 0.052 | **-0.222** | NA | 0.013 | NA |
| Metabolic Problem | NA | **0.244** | 0.028 | -0.073 | **0.364** | NA | 0.021 | -0.08 | -0.076 | NA | 0.096 | NA |
| Neurologic Problem | NA | **0.248** | 0.121 | -0.1 | -0.038 | NA | 0.197 | -0.092 | -0.083 | NA | -0.088 | NA |
| Headache | NA | 0.171 | 0.189 | 0.098 | -0.011 | NA | 0.079 | -0.197 | 0.063 | NA | **-0.285** | NA |
| Other Pain | NA | 0.111 | 0.093 | **-0.256** | 0.141 | NA | **0.291** | -0.212 | -0.074 | NA | **-0.255** | NA |
| Recurrent Problem | NA | 0.062 | -0.069 | 0.054 | 0.067 | NA | 0.211 | -0.043 | 0 | NA | -0.012 | NA |
| Depression | NA | 0.171 | **0.24** | 0.027 | 0.12 | NA | 0.058 | -0.134 | 0.017 | NA | 0.078 | NA |
| Anxiety | NA | 0.152 | 0.163 | -0.08 | 0.087 | NA | 0.164 | -0.05 | -0.05 | NA | -0.028 | NA |
| Other Psychological Problems | NA | **0.265** | 0.035 | -0.137 | 0.338 | NA | 0.1 | -0.073 | 0.051 | NA | -0.192 | NA |
| Medication for Mental Purposes | NA | NA | NA | NA | NA | NA | NA | NA | NA | NA | NA | NA |
| Doctor Visits | NA | 0.117 | -0.147 | -0.148 | **-0.372** | NA | 0.165 | -0.097 | **0.245** | NA | 0.051 | NA |
| Other Medication | NA | 0.143 | 0.092 | 0.155 | 0.128 | NA | -0.029 | -0.023 | -0.086 | NA | -0.007 | NA |
| Antibiotic Taken Past 3 Years | NA | 0.122 | -0.092 | 0.213 | -0.271 | NA | 0.069 | -0.032 | **0.24** | NA | -0.066 | NA |
| Hospitalization Past 5 Years | NA | -0.003 | -0.01 | 0.089 | -0.079 | NA | 0.191 | -0.14 | -0.05 | NA | -0.071 | NA |
| Life Expectancy | NA | 0.085 | -0.03 | 0.001 | 0.064 | NA | -0.013 | -0.054 | 0.169 | NA | -0.192 | NA |
| Now Feeling Physically Miserable | NA | **0.224** | 0.182 | -0.218 | 0.122 | NA | 0.105 | -0.102 | -0.146 | NA | **-0.231** | NA |
| Usually Feeling Physically Miserable | NA | 0.093 | -0.065 | 0.026 | 0.016 | NA | -0.128 | 0.115 | -0.085 | NA | -0.014 | NA |
| Now Feeling Mentally Miserable | NA | 0.026 | **0.302** | -0.175 | 0.243 | NA | 0.003 | -0.168 | -0.1 | NA | -0.05 | NA |
| Usually Feeling Mentally Miserable | NA | 0.175 | 0.118 | -0.028 | 0.111 | NA | -0.069 | 0.102 | -0.105 | NA | -0.018 | NA |
| Year | NA | NA | NA | NA | NA | NA | NA | NA | NA | NA | NA | NA |
| Course of COVID-19 | NA | NA | 0.079 | -0.095 | -0.017 | NA | 0.203 | -0.113 | -0.14 | NA | 0.081 | NA |
| Months since COVID Infection | NA | 0.079 | NA | -0.03 | 0.088 | NA | **0.289** | **-0.514** | **-0.225** | NA | 0.062 | NA |

*Significant correlations are bolded. The p-values that remained or turned significant after the application of the Benjamini-Hochberg correction for multiple testing (with FDR set at 0.1) are underlined.*

**Table S17.** P-values of the correlations between health and performance-related variables and COVID-related variables controlled for age, sex, and survey year-women with at least 36 months elapsed since infection.

|  | Infected | Course | Months since Infection | Vaccination | Months since Vaccination | Covid after Vaccination | Wuhan | Alpha | Delta | Omicron | Age | Sex |
| --- | --- | --- | --- | --- | --- | --- | --- | --- | --- | --- | --- | --- |
| Physical Health Issues | NA | 0.001 | 0.499 | 0.378 | 0.678 | NA | 0.082 | 0.234 | 0.574 | NA | 0.401 | NA |
| Mental Health Issues | NA | 0.272 | 0.059 | 0.442 | 0.218 | NA | 0.68 | 0.862 | 0.444 | NA | 0.515 | NA |
| Fatigue | NA | 0.207 | 0.463 | 0.044 | 0.536 | NA | 0.072 | 0.042 | 0.931 | NA | 0.581 | NA |
| Intelligence | NA | 0.364 | 0.484 | 0.07 | 0.738 | NA | 0.186 | 0.752 | 0.907 | NA | 0.157 | NA |
| Memory | NA | 0.188 | 0.739 | 0.031 | 0.627 | NA | 0.256 | 0.339 | 0.074 | NA | 0.296 | NA |
| Reactions | NA | 0.267 | 0.632 | 0.02 | 0.938 | NA | 0.815 | 0.356 | 0.694 | NA | 0.448 | NA |
| Accuracy | NA | 0.778 | 0.203 | 0.051 | 0.995 | NA | 0.139 | 0.298 | 0.175 | NA | 0.095 | NA |
| Evolutionary Biology Score | NA | 0.231 | 0.734 | 0.806 | 0.601 | NA | 0.994 | 0.523 | 0.19 | NA | 0.034 | NA |
| Free-Recall Memory | NA | 0.346 | 0.94 | 0.119 | 0.822 | NA | 0.175 | 0.314 | 0.039 | NA | 0.353 | NA |
| Recognition Memory | NA | 0.338 | 0.835 | 0.09 | 0.661 | NA | 0.436 | 0.109 | 0.039 | NA | 0.339 | NA |
| Simple RT Duration | NA | 0.154 | 0.016 | 0.021 | 0.191 | NA | 0.365 | 0.238 | 0.072 | NA | 0.914 | NA |
| Simple RT Precision | NA | 0.557 | 0.389 | 0.552 | 19 | NA | 0.196 | 0.337 | 0.739 | NA | 0.966 | NA |
| Stroop Duration | NA | 0.701 | 0.494 | 0.99 | 0.755 | NA | 0.626 | 0.913 | 0.18 | NA | 0.316 | NA |
| Stroop Precision | NA | 0.752 | 0.001 | 0.336 | 0.667 | NA | 0 | 0 | 0.001 | NA | 0.842 | NA |
| Reading Time | NA | 0.916 | 0.549 | 0.771 | 0.549 | NA | 0.721 | 0.667 | 0.857 | NA | 0.622 | NA |
| Allergy | NA | 0.072 | 0.314 | 0.805 | 0.978 | NA | 0.736 | 0.371 | 0.538 | NA | 0.346 | NA |
| Skin Problem | NA | 0.033 | 0.633 | 0.252 | 0.456 | NA | 0.822 | 0.607 | 0.272 | NA | 0.656 | NA |
| Digestive Problem | NA | 0.652 | 0.377 | 0.876 | 0.123 | NA | 0.283 | 0.71 | 0.919 | NA | 0.041 | NA |
| Infection | NA | 0.044 | 0.169 | 0.069 | 0.316 | NA | 0.02 | 0.049 | 0.307 | NA | 0.986 | NA |
| Cardiovascular Problem | NA | 0.65 | 0.14 | 0.524 | 0.055 | NA | 0.173 | 0.317 | 0.722 | NA | 0.577 | NA |
| Low Blood Pressure | NA | 0.14 | 0.568 | 0.054 | 0.229 | NA | 0.708 | 0.174 | 0.045 | NA | 0.003 | NA |
| High Blood Pressure | NA | 0.371 | 0.918 | 2 | 0.259 | NA | 0.308 | 0.035 | 0.559 | NA | 0.829 | NA |
| Orthopedic Problem | NA | 0.892 | 0.933 | 0.98 | 0.688 | NA | 0.521 | 0.644 | 0.049 | NA | 0.905 | NA |
| Metabolic Problem | NA | 0.031 | 0.805 | 0.522 | 0.035 | NA | 0.854 | 0.479 | 0.502 | NA | 0.391 | NA |
| Neurologic Problem | NA | 0.028 | 0.285 | 0.377 | 0.824 | NA | 0.082 | 0.415 | 0.461 | NA | 0.429 | NA |
| Headache | NA | 0.13 | 0.095 | 0.384 | 0.95 | NA | 0.486 | 0.082 | 0.576 | NA | 0.011 | NA |
| Other Pain | NA | 0.325 | 0.411 | 0.024 | 0.414 | NA | 0.01 | 0.061 | 0.513 | NA | 0.022 | NA |
| Recurrent Problem | NA | 0.584 | 0.544 | 0.636 | 0.7 | NA | 0.063 | 0.705 | 0.997 | NA | 0.914 | NA |
| Depression | NA | 0.132 | 0.034 | 0.808 | 0.485 | NA | 0.608 | 0.237 | 0.879 | NA | 0.484 | NA |
| Anxiety | NA | 0.178 | 0.149 | 0.479 | 0.615 | NA | 0.146 | 0.656 | 0.661 | NA | 0.801 | NA |
| Other Psychological Problems | NA | 0.019 | 0.755 | 0.226 | 0.05 | NA | 0.379 | 0.518 | 0.655 | NA | 0.084 | NA |
| Medication for Mental Purposes | NA | NA | NA | NA | NA | NA | NA | NA | NA | NA | NA | NA |
| Doctor Visits | NA | 0.302 | 0.194 | 0.191 | 0.031 | NA | 0.144 | 0.393 | 0.031 | NA | 0.644 | NA |
| Other Medication | NA | 0.243 | 0.451 | 0.205 | 0.474 | NA | 0.811 | 0.85 | 0.48 | NA | 0.956 | NA |
| Antibiotic Taken Past 3 Years | NA | 0.28 | 0.416 | 0.06 | 0.116 | NA | 0.54 | 0.776 | 0.034 | NA | 0.552 | NA |
| Hospitalization Past 5 Years | NA | 0.981 | 0.931 | 0.431 | 0.645 | NA | 0.092 | 0.215 | 0.659 | NA | 0.523 | NA |
| Life Expectancy | NA | 0.454 | 0.792 | 0.993 | 0.709 | NA | 0.907 | 0.632 | 0.135 | NA | 0.086 | NA |
| Now Feeling Physically Miserable | NA | 0.047 | 0.108 | 0.054 | 0.478 | NA | 0.352 | 0.369 | 0.196 | NA | 0.038 | NA |
| Usually Feeling Physically Miserable | NA | 0.412 | 0.568 | 0.817 | 0.928 | NA | 0.259 | 0.309 | 0.452 | NA | 0.898 | NA |
| Now Feeling Mentally Miserable | NA | 0.82 | 0.008 | 0.121 | 0.159 | NA | 0.975 | 0.137 | 0.375 | NA | 0.655 | NA |
| Usually Feeling Mentally Miserable | NA | 0.122 | 0.298 | 0.807 | 0.522 | NA | 0.541 | 0.367 | 0.354 | NA | 0.875 | NA |
| Year | NA | NA | NA | NA | NA | NA | NA | NA | NA | NA | NA | NA |
| Course of COVID-19 | NA | NA | 0.484 | 0.399 | 0.922 | NA | 0.073 | 0.32 | 0.217 | NA | 0.466 | NA |
| Months since COVID Infection | NA | 0.484 | NA | 0.794 | 0.611 | NA | 0.011 | 0 | 0.047 | NA | 0.578 | NA |

*Significant correlations are bolded. The p-values that remained or turned significant after the application of the Benjamini-Hochberg correction for multiple testing (with FDR set at 0.1) are underlined.*

**Table S18.** Correlations between health and performance-related variables and COVID-related variables controlled for age, sex, and survey year-men with at least 36 months elapsed since infection.

|  | Infected | Course | Months since Infection | Vaccination | Months since Vaccination | Covid after Vaccination | Wuhan | Alpha | Delta | Omicron | Age | Sex |
| --- | --- | --- | --- | --- | --- | --- | --- | --- | --- | --- | --- | --- |
| Physical Health Issues | NA | 0.232 | -0.328 | NA | -0.055 | NA | <u>-0.426</u> | 0.426 | NA | NA | -0.144 | NA |
| Mental Health Issues | NA | 0.004 | -0.414 | NA | -0.464 | NA | -0.389 | 0.389 | NA | NA | -0.016 | NA |
| Fatigue | NA | -0.341 | -0.159 | NA | -0.706 | NA | -0.017 | 0.017 | NA | NA | 0.094 | NA |
| Intelligence | NA | 0.165 | 0.129 | NA | 0.468 | NA | -0.057 | 0.057 | NA | NA | 0.307 | NA |
| Memory | NA | -0.241 | 0.314 | NA | 0.342 | NA | 0.36 | -0.36 | NA | NA | 0.394 | NA |
| Reactions | NA | -0.056 | 0.35 | NA | 0.196 | NA | **0.534** | **-0.534** | NA | NA | 0.208 | NA |
| Accuracy | NA | 0.065 | 0.06 | NA | -0.342 | NA | -0.359 | 0.359 | NA | NA | -0.24 | NA |
| Evolutionary Biology Score | NA | -0.004 | -0.091 | NA | -0.148 | NA | -0.258 | 0.258 | NA | NA | **-0.504** | NA |
| Free-Recall Memory | NA | -0.259 | 0.178 | NA | 0.274 | NA | 0.221 | -0.221 | NA | NA | 0.289 | NA |
| Recognition Memory | NA | -0.309 | 0.281 | NA | 0.274 | NA | 0.35 | -0.35 | NA | NA | 0.379 | NA |
| Simple RT Duration | NA | -0.107 | 0.159 | NA | 0.071 | NA | 0.205 | -0.205 | NA | NA | 0.112 | NA |
| Simple RT Precision | NA | NA | NA | NA | NA | NA | NA | NA | NA | NA | NA | NA |
| Stroop Duration | NA | -0.218 | 0.207 | NA | -0.189 | NA | 0.288 | -0.288 | NA | NA | -0.048 | NA |
| Stroop Precision | NA | NA | NA | NA | NA | NA | NA | NA | NA | NA | NA | NA |
| Reading Time | NA | -0.203 | 0.371 | NA | 0.189 | NA | **0.479** | **-0.479** | NA | NA | 0.176 | NA |
| Allergy | NA | 0.202 | **-0.666** | NA | -0.508 | NA | **-0.52** | **0.52** | NA | NA | 0.124 | NA |
| Skin Problem | NA | -0.027 | -0.183 | NA | -0.225 | NA | -0.165 | 0.165 | NA | NA | -0.401 | NA |
| Digestive Problem | NA | -0.016 | 0.071 | NA | -0.76 | NA | -0.232 | 0.232 | NA | NA | -0.407 | NA |
| Infection | NA | 0.094 | -0.197 | NA | -0.286 | NA | -0.361 | 0.361 | NA | NA | -0.154 | NA |
| Cardiovascular Problem | NA | 0.2 | **-0.478** | NA | NA | NA | **-0.665** | **0.665** | NA | NA | -0.353 | NA |
| Low Blood Pressure | NA | -0.03 | -0.052 | NA | 0.582 | NA | 0.166 | -0.166 | NA | NA | 0.34 | NA |
| High Blood Pressure | NA | NA | NA | NA | NA | NA | NA | NA | NA | NA | NA | NA |
| Orthopedic Problem | NA | 0.053 | 0.112 | NA | -0.019 | NA | -0.037 | 0.037 | NA | NA | 0.035 | NA |
| Metabolic Problem | NA | 0.414 | -0.2 | NA | NA | NA | 0.092 | -0.092 | NA | NA | 0.275 | NA |
| Neurologic Problem | NA | NA | NA | NA | NA | NA | NA | NA | NA | NA | NA | NA |
| Headache | NA | 0.054 | -0.219 | NA | -0.141 | NA | -0.393 | 0.393 | NA | NA | -0.201 | NA |
| Other Pain | NA | 0.096 | -0.252 | NA | 0.183 | NA | -0.363 | 0.363 | NA | NA | -0.266 | NA |
| Recurrent Problem | NA | -0.096 | -0.084 | NA | -0.292 | NA | -0.407 | 0.407 | NA | NA | **-0.506** | NA |
| Depression | NA | -0.247 | -0.337 | NA | -0.605 | NA | -0.449 | 0.449 | NA | NA | -0.125 | NA |
| Anxiety | NA | -0.282 | -0.39 | NA | -0.499 | NA | -0.331 | 0.331 | NA | NA | -0.072 | NA |
| Other Psychological Problems | NA | -0.13 | -0.401 | NA | -0.299 | NA | -0.212 | 0.212 | NA | NA | 0.276 | NA |
| Medication for Mental Purposes | NA | NA | NA | NA | NA | NA | NA | NA | NA | NA | NA | NA |
| Doctor Visits | NA | 0.351 | -0.095 | NA | 0.183 | NA | -0.407 | 0.407 | NA | NA | -0.122 | NA |
| Other Medication | NA | 0.44 | -0.474 | NA | NA | NA | -0.384 | 0.384 | NA | NA | -0.067 | NA |
| Antibiotic Taken Past 3 Years | NA | 0.36 | 0.054 | NA | 0.299 | NA | -0.002 | 0.002 | NA | NA | **-0.519** | NA |
| Hospitalization Past 5 Years | NA | 0.104 | -0.301 | NA | -0.109 | NA | -0.154 | 0.154 | NA | NA | -0.071 | NA |
| Life Expectancy | NA | -0.079 | 0.026 | NA | -0.327 | NA | 0.065 | -0.065 | NA | NA | **-0.502** | NA |
| Now Feeling Physically Miserable | NA | 0.22 | -0.253 | NA | 0.41 | NA | -0.238 | 0.238 | NA | NA | 0.168 | NA |
| Usually Feeling Physically Miserable | NA | 0.055 | -0.189 | NA | 0.2 | NA | -0.186 | 0.186 | NA | NA | -0.118 | NA |
| Now Feeling Mentally Miserable | NA | 0.273 | **-0.493** | NA | -0.334 | NA | **-0.473** | **0.473** | NA | NA | 0.281 | NA |
| Usually Feeling Mentally Miserable | NA | -0.151 | -0.404 | NA | -0.612 | NA | -0.452 | 0.452 | NA | NA | -0.071 | NA |
| Year | NA | NA | NA | NA | NA | NA | NA | NA | NA | NA | NA | NA |
| Course of COVID-19 | NA | NA | -0.045 | NA | 0.422 | NA | -0.188 | 0.188 | NA | NA | 0.255 | NA |
| Months since COVID Infection | NA | -0.045 | NA | NA | 0.378 | NA | **0.631** | **-0.631** | NA | NA | -0.272 | NA |

*Significant correlations are bolded. The p-values that remained or turned significant after the application of the Benjamini-Hochberg correction for multiple testing (with FDR set at 0.1) are underlined.*

**Table S19.** P-values of the correlations between health and performance-related variables and COVID-related variables controlled for age, sex, and survey year-men with at least 36 months elapsed since infection.

|  | Infected | Course | Months since Infection | Vaccination | Months since Vaccination | Covid after Vaccination | Wuhan | Alpha | Delta | Omicron | Age | Sex |
| --- | --- | --- | --- | --- | --- | --- | --- | --- | --- | --- | --- | --- |
| Physical Health Issues | NA | 0.32 | 0.161 | NA | 0.894 | NA | 0.068 | 0.068 | NA | NA | 0.514 | NA |
| Mental Health Issues | NA | 0.986 | 0.076 | NA | 0.255 | NA | 0.096 | 0.096 | NA | NA | 0.942 | NA |
| Fatigue | NA | 0.144 | 0.497 | NA | 0.084 | NA | 0.943 | 0.943 | NA | NA | 0.671 | NA |
| Intelligence | NA | 0.48 | 0.582 | NA | 0.252 | NA | 0.807 | 0.807 | NA | NA | 0.165 | NA |
| Memory | NA | 0.302 | 0.179 | NA | 0.402 | NA | 0.123 | 0.123 | NA | NA | 0.075 | NA |
| Reactions | NA | 0.81 | 0.134 | NA | 0.63 | NA | 0.022 | 0.022 | NA | NA | 0.346 | NA |
| Accuracy | NA | 0.78 | 0.798 | NA | 0.402 | NA | 0.124 | 0.124 | NA | NA | 0.277 | NA |
| Evolutionary Biology Score | NA | 0.987 | 0.696 | NA | 0.717 | NA | 0.269 | 0.269 | NA | NA | 0.022 | NA |
| Free-Recall Memory | NA | 0.297 | 0.473 | NA | 0.503 | NA | 0.375 | 0.375 | NA | NA | 0.216 | NA |
| Recognition Memory | NA | 0.186 | 0.229 | NA | 0.503 | NA | 0.134 | 0.134 | NA | NA | 0.086 | NA |
| Simple RT Duration | NA | 0.647 | 0.497 | NA | 0.861 | NA | 0.38 | 0.38 | NA | NA | 0.612 | NA |
| Simple RT Precision | NA | NA | NA | NA | NA | NA | NA | NA | NA | NA | NA | NA |
| Stroop Duration | NA | 0.351 | 0.375 | NA | 0.643 | NA | 0.217 | 0.217 | NA | NA | 0.828 | NA |
| Stroop Precision | NA | NA | NA | NA | NA | NA | NA | NA | NA | NA | NA | NA |
| Reading Time | NA | 0.384 | 0.112 | NA | 0.643 | NA | 0.04 | 0.04 | NA | NA | 0.425 | NA |
| Allergy | NA | 0.387 | 0.004 | NA | 0.213 | NA | 0.026 | 0.026 | NA | NA | 0.575 | NA |
| Skin Problem | NA | 0.906 | 0.434 | NA | 0.582 | NA | 0.479 | 0.479 | NA | NA | 0.07 | NA |
| Digestive Problem | NA | 0.946 | 0.762 | NA | 0.063 | NA | 0.321 | 0.321 | NA | NA | 0.066 | NA |
| Infection | NA | 0.686 | 0.398 | NA | 0.484 | NA | 0.122 | 0.122 | NA | NA | 0.486 | NA |
| Cardiovascular Problem | NA | 0.391 | 0.041 | NA | 6 | NA | 0.004 | 0.004 | NA | NA | 0.11 | NA |
| Low Blood Pressure | NA | 0.903 | 0.834 | NA | 0.236 | NA | 0.504 | 0.504 | NA | NA | 0.145 | NA |
| High Blood Pressure | NA | NA | NA | NA | NA | NA | NA | NA | NA | NA | NA | NA |
| Orthopedic Problem | NA | 0.819 | 0.63 | NA | 0.963 | NA | 0.876 | 0.876 | NA | NA | 0.875 | NA |
| Metabolic Problem | NA | 0.077 | 0.392 | NA | 6 | NA | 0.694 | 0.694 | NA | NA | 0.214 | NA |
| Neurologic Problem | NA | NA | NA | NA | NA | NA | NA | NA | NA | NA | NA | NA |
| Headache | NA | 0.817 | 0.348 | NA | 0.73 | NA | 0.093 | 0.093 | NA | NA | 0.363 | NA |
| Other Pain | NA | 0.68 | 0.28 | NA | 0.654 | NA | 0.12 | 0.12 | NA | NA | 0.229 | NA |
| Recurrent Problem | NA | 0.681 | 0.72 | NA | 0.474 | NA | 0.081 | 0.081 | NA | NA | 0.022 | NA |
| Depression | NA | 0.29 | 0.149 | NA | 0.138 | NA | 0.054 | 0.054 | NA | NA | 0.571 | NA |
| Anxiety | NA | 0.228 | 0.095 | NA | 0.222 | NA | 0.156 | 0.156 | NA | NA | 0.746 | NA |
| Other Psychological Problems | NA | 0.601 | 0.107 | NA | 0.464 | NA | 0.394 | 0.394 | NA | NA | 0.237 | NA |
| Medication for Mental Purposes | NA | NA | NA | NA | NA | NA | NA | NA | NA | NA | NA | NA |
| Doctor Visits | NA | 0.132 | 0.684 | NA | 0.654 | NA | 0.081 | 0.081 | NA | NA | 0.581 | NA |
| Other Medication | NA | 0.076 | 0.057 | NA | 6 | NA | 0.122 | 0.122 | NA | NA | 0.775 | NA |
| Antibiotic Taken Past 3 Years | NA | 0.123 | 0.816 | NA | 0.464 | NA | 0.994 | 0.994 | NA | NA | 0.019 | NA |
| Hospitalization Past 5 Years | NA | 0.655 | 0.197 | NA | 0.79 | NA | 0.511 | 0.511 | NA | NA | 0.747 | NA |
| Life Expectancy | NA | 0.737 | 0.912 | NA | 0.423 | NA | 0.779 | 0.779 | NA | NA | 0.023 | NA |
| Now Feeling Physically Miserable | NA | 0.345 | 0.278 | NA | 0.316 | NA | 0.307 | 0.307 | NA | NA | 0.447 | NA |
| Usually Feeling Physically Miserable | NA | 0.814 | 0.419 | NA | 0.623 | NA | 0.426 | 0.426 | NA | NA | 0.594 | NA |
| Now Feeling Mentally Miserable | NA | 0.242 | 0.035 | NA | 0.413 | NA | 0.043 | 0.043 | NA | NA | 0.203 | NA |
| Usually Feeling Mentally Miserable | NA | 0.517 | 0.084 | NA | 0.134 | NA | 0.053 | 0.053 | NA | NA | 0.748 | NA |
| Year | NA | NA | NA | NA | NA | NA | NA | NA | NA | NA | NA | NA |
| Course of COVID-19 | NA | NA | 0.847 | NA | 0.301 | NA | 0.42 | 0.42 | NA | NA | 0.248 | NA |
| Months since COVID Infection | NA | 0.847 | NA | NA | 0.355 | NA | 0.007 | 0.007 | NA | NA | 0.218 | NA |

*Significant correlations are bolded. The p-values that remained or turned significant after the application of the Benjamini-Hochberg correction for multiple testing (with FDR set at 0.1) are underlined.*

**Table S20.** Correlations between health and performance-related variables and COVID-related variables controlled for age, sex, and survey year-All subjects.

|  | Infected | Course | Months since Infection | Vaccination | Months since Vaccination | Covid after Vaccination | Wuhan | Alpha | Delta | Omicron | Age | Sex |
| --- | --- | --- | --- | --- | --- | --- | --- | --- | --- | --- | --- | --- |
| Physical Health Issues | 0.012 | **0.211** | 0.013 | **0.062** | 0.013 | -0.012 | 0.05 | -0.024 | 0.001 | -0.029 | 0.012 | **0.163** |
| Mental Health Issues | -0.049 | 0.066 | -0.045 | 0.048 | -0.015 | -0.025 | -0.051 | 0.018 | -0.023 | 0.052 | 0.036 | **0.162** |
| Fatigue | -0.042 | **0.093** | 0.044 | **0.089** | 0.036 | -0.005 | **0.126** | **-0.103** | -0.062 | -0.007 | -0.005 | **0.097** |
| Intelligence | 0.021 | -0.057 | -0.071 | -0.044 | 0.005 | **0.171** | **-0.099** | -0.076 | 0.018 | **0.172** | -0.021 | **-0.099** |
| Memory | 0.045 | -0.01 | -0.053 | -0.039 | -0.011 | 0.029 | **-0.119** | -0.035 | 0.065 | 0.068 | **-0.055** | **0.111** |
| Reactions | 0.029 | 0.029 | 0.036 | **-0.075** | 0.027 | -0.067 | **0.097** | **-0.095** | -0.025 | -0.024 | -0.009 | 0.02 |
| Accuracy | 0.001 | **-0.082** | -0.025 | -0.015 | 0.051 | **0.194** | -0.023 | -0.073 | 0.01 | 0.075 | -0.053 | **-0.096** |
| Evolutionary Biology Score | -0.014 | 0.009 | 0.028 | 0.025 | 0.065 | **0.117** | 0.001 | 0.052 | 0.011 | -0.047 | **-0.099** | **-0.073** |
| Free-Recall Memory | 0.053 | -0.017 | -0.063 | -0.004 | -0.014 | 0.063 | **-0.095** | -0.043 | 0.052 | 0.057 | -0.052 | **0.108** |
| Recognition Memory | **0.057** | -0.018 | -0.063 | -0.012 | -0.011 | 0.065 | **-0.094** | -0.075 | 0.081 | 0.055 | -0.043 | **0.117** |
| Simple RT Duration | 0.005 | -0.068 | 0.041 | **-0.058** | 0.043 | 0.009 | 0.066 | -0.048 | -0.037 | -0.018 | 0.035 | 0.046 |
| Simple RT Precision | 0.007 | -0.009 | -0.018 | -0.05 | 0.045 | 0.014 | 0.019 | **-0.194** | 0.067 | 0.054 | -0.008 | **-0.074** |
| Stroop Duration | 0.015 | 0.04 | 0.036 | -0.051 | 0.013 | -0.043 | 0.083 | -0.069 | 0.016 | -0.062 | 0.001 | -0.002 |
| Stroop Precision | -0.006 | **-0.095** | -0.019 | -0.011 | 0.044 | **0.12** | 0.041 | -0.042 | -0.07 | 0.043 | -0.016 | -0.008 |
| Reading Time | 0.041 | -0.011 | 0.044 | -0.049 | -0.01 | -0.073 | 0.055 | -0.053 | -0.017 | -0.027 | -0.002 | 0.053 |
| Allergy | 0.017 | **0.128** | 0.009 | **0.067** | 0.019 | 0.022 | 0.041 | 0.001 | 0.022 | -0.045 | -0.006 | **0.07** |
| Skin Problem | 0.016 | **0.164** | 0.002 | 0.016 | 0.026 | **0.128** | 0.013 | -0.055 | 0.032 | 0.001 | -0.03 | -0.018 |
| Digestive Problem | -0.003 | **0.125** | -0.02 | **0.064** | 0.04 | -0.052 | 0.001 | 0.04 | -0.04 | 0.024 | 0.026 | 0.004 |
| Infection | **0.101** | **0.189** | -0.001 | 0.03 | -0.016 | 0.065 | 0.06 | **-0.127** | 0.017 | 0.037 | **-0.055** | **0.055** |
| Cardiovascular Problem | 0.017 | 0.004 | 0.03 | **0.075** | -0.033 | -0.014 | 0.051 | -0.01 | -0.063 | 0.019 | -0.029 | 0.026 |
| Low Blood Pressure | -0.046 | 0.02 | 0.064 | 0.043 | 0.048 | -0.103 | 0.067 | 0.012 | -0.015 | -0.078 | -0.013 | **0.254** |
| High Blood Pressure | -0.06 | 0 | 0.022 | 0.034 | 0.089 | **0.16** | -0.053 | -0.006 | 0.084 | -0.01 | **0.086** | -0.034 |
| Orthopedic Problem | 0.026 | 0.046 | 0.037 | 0.048 | 0.024 | -0.01 | 0.02 | **0.108** | -0.031 | -0.08 | 0.046 | **0.159** |
| Metabolic Problem | 0.04 | 0.043 | 0.027 | -0.017 | -0.018 | -0.002 | 0.003 | -0.01 | 0.064 | -0.065 | 0.021 | **0.096** |
| Neurologic Problem | **0.061** | **0.125** | -0.033 | -0.043 | 0.026 | 0.074 | 0 | -0.058 | 0.015 | 0.044 | 0.012 | **0.079** |
| Headache | -0.005 | **0.13** | 0.015 | **0.109** | -0.025 | 0 | 0.012 | 0.047 | -0.021 | -0.041 | -0.043 | **0.161** |
| Other Pain | -0.02 | **0.141** | 0.022 | 0.005 | 0.004 | 0.01 | 0.072 | -0.015 | -0.004 | -0.056 | -0.028 | **0.089** |
| Recurrent Problem | 0.027 | **0.08** | 0.019 | **0.056** | -0.012 | 0.053 | **0.125** | -0.017 | -0.085 | -0.017 | 0.031 | **0.07** |
| Depression | 0.008 | **0.088** | -0.084 | 0.014 | -0.014 | -0.054 | -0.074 | -0.029 | -0.029 | **0.128** | **0.069** | **0.081** |
| Anxiety | -0.008 | 0.07 | -0.053 | 0.046 | 0.002 | 0.071 | -0.008 | -0.06 | -0.054 | **0.12** | 0.035 | **0.156** |
| Other Psychological Problems | -0.033 | **0.115** | 0.027 | 0.026 | -0.001 | -0.033 | 0.035 | 0.004 | -0.05 | 0.013 | 0.008 | **0.072** |
| Medication for Mental Purposes | -0.022 | **0.155** | -0.074 | 0.045 | 0.003 | -0.04 | -0.078 | -0.026 | 0.036 | 0.077 | 0.035 | **0.118** |
| Doctor Visits | 0.046 | **0.144** | -0.085 | 0.024 | 0.016 | -0.014 | -0.043 | 0.049 | -0.038 | 0.08 | -0.05 | -0.009 |
| Other Medication | -0.052 | **0.186** | -0.024 | **0.118** | 0 | 0.084 | 0.017 | -0.069 | -0.044 | 0.059 | 0.004 | **0.137** |
| Antibiotic Taken Past 3 Years | **0.063** | **0.099** | -0.01 | 0.002 | 0.034 | 0.013 | -0.014 | -0.014 | 0.022 | 0.03 | -0.016 | **0.117** |
| Hospitalization Past 5 Years | 0.019 | **0.125** | 0.017 | -0.005 | 0.001 | -0.082 | 0.05 | 0.002 | 0.022 | -0.071 | -0.028 | **-0.059** |
| Life Expectancy | -0.016 | **0.096** | 0.058 | 0.037 | -0.024 | **-0.128** | 0.011 | -0.013 | 0.06 | -0.063 | -0.021 | -0.038 |
| Now Feeling Physically Miserable | -0.007 | 0.074 | -0.002 | 0.016 | -0.007 | 0.02 | -0.008 | -0.01 | -0.016 | 0.027 | **0.074** | **0.103** |
| Usually Feeling Physically Miserable | -0.039 | 0.05 | -0.061 | 0.029 | -0.047 | -0.022 | -0.07 | -0.009 | 0.036 | 0.038 | 0.029 | **0.17** |
| Now Feeling Mentally Miserable | -0.022 | 0.053 | 0.001 | -0.006 | -0.028 | -0.011 | -0.017 | 0.01 | 0.024 | -0.043 | 0.043 | **0.129** |
| Usually Feeling Mentally Miserable | **-0.088** | 0.034 | -0.068 | **0.071** | -0.015 | -0.097 | -0.072 | **0.116** | -0.037 | 0.027 | 0.021 | **0.149** |
| Year | **0.37** | **0.106** | **0.618** | **-0.096** | **0.71** | 0.09 | **-0.148** | **-0.132** | -0.018 | **0.248** | 0.018 | 0.05 |
| Course of COVID-19 | NA | NA | -0.01 | -0.037 | -0.007 | 0.032 | 0.033 | -0.034 | -0.001 | 0.001 | **0.101** | 0.05 |
| Months since COVID Infection | -0.036 | -0.01 | NA | 0.032 | **0.144** | **-0.405** | **0.47** | 0.032 | **-0.103** | **-0.479** | 0.007 | -0.034 |

*Significant correlations are bolded. The p-values that remained or turned significant after the application of the Benjamini-Hochberg correction for multiple testing (with FDR set at 0.1) are underlined.*

**Table S21.** P-values of the correlations between health and performance-related variables and COVID-related variables controlled for age, sex, and survey year-All subjects.

|  | Infected | Course | Months since Infection | Vaccination | Months since Vaccination | Covid after Vaccination | Wuhan | Alpha | Delta | Omicron | Age | Sex |
| --- | --- | --- | --- | --- | --- | --- | --- | --- | --- | --- | --- | --- |
| Physical Health Issues | 0.655 | 0 | 0.769 | 0.026 | 0.707 | 0.828 | 0.251 | 0.585 | 0.988 | 0.51 | 0.672 | 0 |
| Mental Health Issues | 0.079 | 0.101 | 0.298 | 0.084 | 0.67 | 0.653 | 0.245 | 0.675 | 0.601 | 0.237 | 0.191 | 0 |
| Fatigue | 0.128 | 0.021 | 0.313 | 0.002 | 0.314 | 0.931 | 0.004 | 0.018 | 0.159 | 0.881 | 0.846 | 0.001 |
| Intelligence | 0.451 | 0.161 | 0.106 | 0.122 | 0.887 | 0.002 | 0.024 | 0.083 | 0.676 | 0 | 0.451 | 0 |
| Memory | 0.108 | 0.809 | 0.227 | 0.167 | 0.757 | 0.612 | 0.007 | 0.428 | 0.141 | 0.119 | 0.048 | 0 |
| Reactions | 0.302 | 0.473 | 0.411 | 0.007 | 0.444 | 0.23 | 0.027 | 0.03 | 0.565 | 0.584 | 0.744 | 0.467 |
| Accuracy | 0.973 | 0.041 | 0.566 | 0.587 | 0.145 | 0.001 | 0.599 | 0.093 | 0.812 | 0.088 | 0.057 | 0.001 |
| Evolutionary Biology Score | 0.615 | 0.828 | 0.521 | 0.365 | 0.064 | 0.037 | 0.989 | 0.237 | 0.799 | 0.283 | 0 | 0.009 |
| Free-Recall Memory | 0.071 | 0.68 | 0.166 | 0.892 | 0.693 | 0.279 | 0.038 | 0.348 | 0.252 | 0.213 | 0.076 | 0 |
| Recognition Memory | 0.048 | 0.661 | 0.172 | 0.672 | 0.762 | 0.257 | 0.04 | 0.1 | 0.077 | 0.233 | 0.139 | 0 |
| Simple RT Duration | 0.858 | 0.089 | 0.346 | 0.039 | 0.223 | 0.874 | 0.133 | 0.268 | 0.393 | 0.673 | 0.213 | 0.1 |
| Simple RT Precision | 0.794 | 0.821 | 0.686 | 0.072 | 0.206 | 0.809 | 0.67 | 0 | 0.125 | 0.217 | 0.782 | 0.007 |
| Stroop Duration | 0.587 | 0.318 | 0.409 | 0.067 | 0.718 | 0.442 | 0.057 | 0.115 | 0.718 | 0.159 | 0.972 | 0.933 |
| Stroop Precision | 0.817 | 0.019 | 0.657 | 0.686 | 0.217 | 0.032 | 0.348 | 0.341 | 0.111 | 0.323 | 0.567 | 0.775 |
| Reading Time | 0.144 | 0.778 | 0.313 | 0.082 | 0.771 | 0.19 | 0.212 | 0.226 | 0.693 | 0.535 | 0.941 | 0.056 |
| Allergy | 0.545 | 0.001 | 0.83 | 0.016 | 0.582 | 0.687 | 0.344 | 0.991 | 0.622 | 0.299 | 0.821 | 0.012 |
| Skin Problem | 0.573 | 0 | 0.957 | 0.573 | 0.458 | 0.022 | 0.769 | 0.207 | 0.471 | 0.989 | 0.28 | 0.522 |
| Digestive Problem | 0.903 | 0.002 | 0.645 | 0.023 | 0.254 | 0.354 | 0.98 | 0.358 | 0.36 | 0.588 | 0.35 | 0.875 |
| Infection | 0 | 0 | 0.982 | 0.284 | 0.65 | 0.243 | 0.174 | 0.004 | 0.696 | 0.397 | 0.047 | 0.047 |
| Cardiovascular Problem | 0.548 | 0.915 | 0.489 | 0.007 | 0.343 | 0.798 | 0.239 | 0.82 | 0.151 | 0.664 | 0.294 | 0.351 |
| Low Blood Pressure | 0.138 | 0.653 | 0.183 | 0.167 | 0.216 | 0.096 | 0.165 | 0.804 | 0.751 | 0.106 | 0.685 | 0 |
| High Blood Pressure | 0.134 | 0.998 | 0.712 | 0.413 | 0.078 | 0.041 | 0.381 | 0.924 | 0.166 | 0.871 | 0.032 | 0.402 |
| Orthopedic Problem | 0.354 | 0.248 | 0.395 | 0.085 | 0.497 | 0.852 | 0.653 | 0.013 | 0.476 | 0.069 | 0.094 | 0 |
| Metabolic Problem | 0.157 | 0.287 | 0.54 | 0.553 | 0.614 | 0.969 | 0.951 | 0.824 | 0.145 | 0.138 | 0.442 | 0.001 |
| Neurologic Problem | 0.029 | 0.002 | 0.455 | 0.129 | 0.459 | 0.187 | 0.991 | 0.183 | 0.737 | 0.319 | 0.665 | 0.005 |
| Headache | 0.853 | 0.001 | 0.73 | 0 | 0.479 | 0.993 | 0.79 | 0.286 | 0.625 | 0.347 | 0.122 | 0 |
| Other Pain | 0.484 | 0 | 0.622 | 0.859 | 0.901 | 0.863 | 0.099 | 0.738 | 0.923 | 0.2 | 0.311 | 0.001 |
| Recurrent Problem | 0.33 | 0.048 | 0.67 | 0.045 | 0.741 | 0.341 | 0.004 | 0.692 | 0.053 | 0.698 | 0.273 | 0.012 |
| Depression | 0.775 | 0.029 | 0.054 | 0.623 | 0.691 | 0.329 | 0.091 | 0.508 | 0.501 | 0.003 | 0.013 | 0.003 |
| Anxiety | 0.773 | 0.084 | 0.221 | 0.104 | 0.95 | 0.202 | 0.85 | 0.17 | 0.215 | 0.006 | 0.214 | 0 |
| Other Psychological Problems | 0.233 | 0.005 | 0.543 | 0.355 | 0.976 | 0.561 | 0.42 | 0.929 | 0.252 | 0.761 | 0.762 | 0.01 |
| Medication for Mental Purposes | 0.449 | 0 | 0.104 | 0.126 | 0.939 | 0.491 | 0.086 | 0.576 | 0.426 | 0.09 | 0.224 | 0 |
| Doctor Visits | 0.098 | 0 | 0.052 | 0.397 | 0.643 | 0.8 | 0.33 | 0.265 | 0.39 | 0.068 | 0.074 | 0.741 |
| Other Medication | 0.073 | 0 | 0.592 | 0 | 0.991 | 0.143 | 0.704 | 0.131 | 0.335 | 0.191 | 0.885 | 0 |
| Antibiotic Taken Past 3 Years | 0.024 | 0.014 | 0.825 | 0.95 | 0.338 | 0.812 | 0.753 | 0.747 | 0.61 | 0.486 | 0.572 | 0 |
| Hospitalization Past 5 Years | 0.49 | 0.002 | 0.692 | 0.846 | 0.978 | 0.145 | 0.25 | 0.966 | 0.614 | 0.107 | 0.315 | 0.034 |
| Life Expectancy | 0.579 | 0.018 | 0.186 | 0.195 | 0.507 | 0.022 | 0.803 | 0.764 | 0.171 | 0.153 | 0.465 | 0.177 |
| Now Feeling Physically Miserable | 0.8 | 0.066 | 0.967 | 0.578 | 0.843 | 0.725 | 0.848 | 0.817 | 0.71 | 0.538 | 0.008 | 0 |
| Usually Feeling Physically Miserable | 0.158 | 0.213 | 0.165 | 0.299 | 0.183 | 0.696 | 0.111 | 0.829 | 0.415 | 0.381 | 0.301 | 0 |
| Now Feeling Mentally Miserable | 0.433 | 0.185 | 0.988 | 0.821 | 0.419 | 0.847 | 0.704 | 0.818 | 0.585 | 0.323 | 0.119 | 0 |
| Usually Feeling Mentally Miserable | 0.002 | 0.393 | 0.119 | 0.011 | 0.671 | 0.081 | 0.101 | 0.008 | 0.4 | 0.544 | 0.439 | 0 |
| Year | 0 | 0.008 | 0 | 0.001 | 0 | 0.107 | 0.001 | 0.003 | 0.688 | 0 | 0.51 | 0.073 |
| Course of COVID-19 | NA | NA | 0.822 | 0.364 | 0.895 | 0.562 | 0.45 | 0.444 | 0.983 | 0.974 | 0.012 | 0.211 |
| Months since COVID Infection | 0.411 | 0.822 | NA | 0.476 | 0.01 | 0 | 0 | 0.465 | 0.018 | 0 | 0.877 | 0.431 |

**Table S22.**
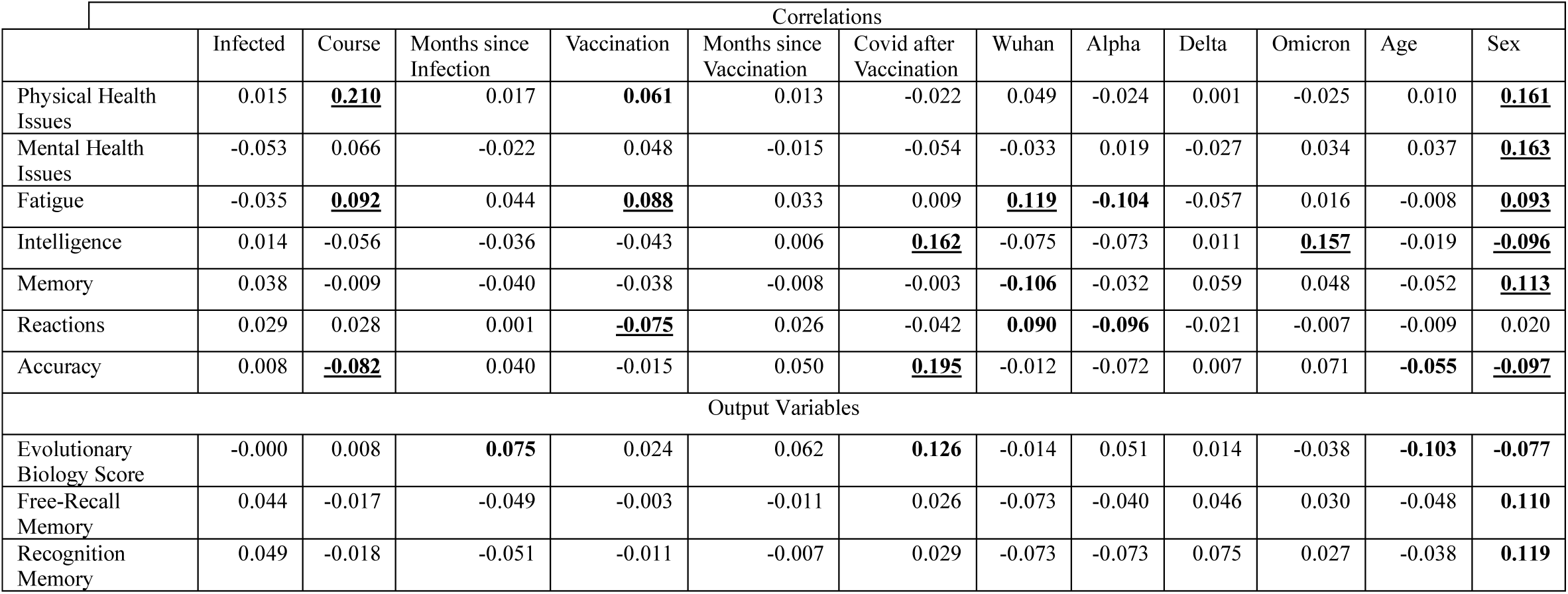
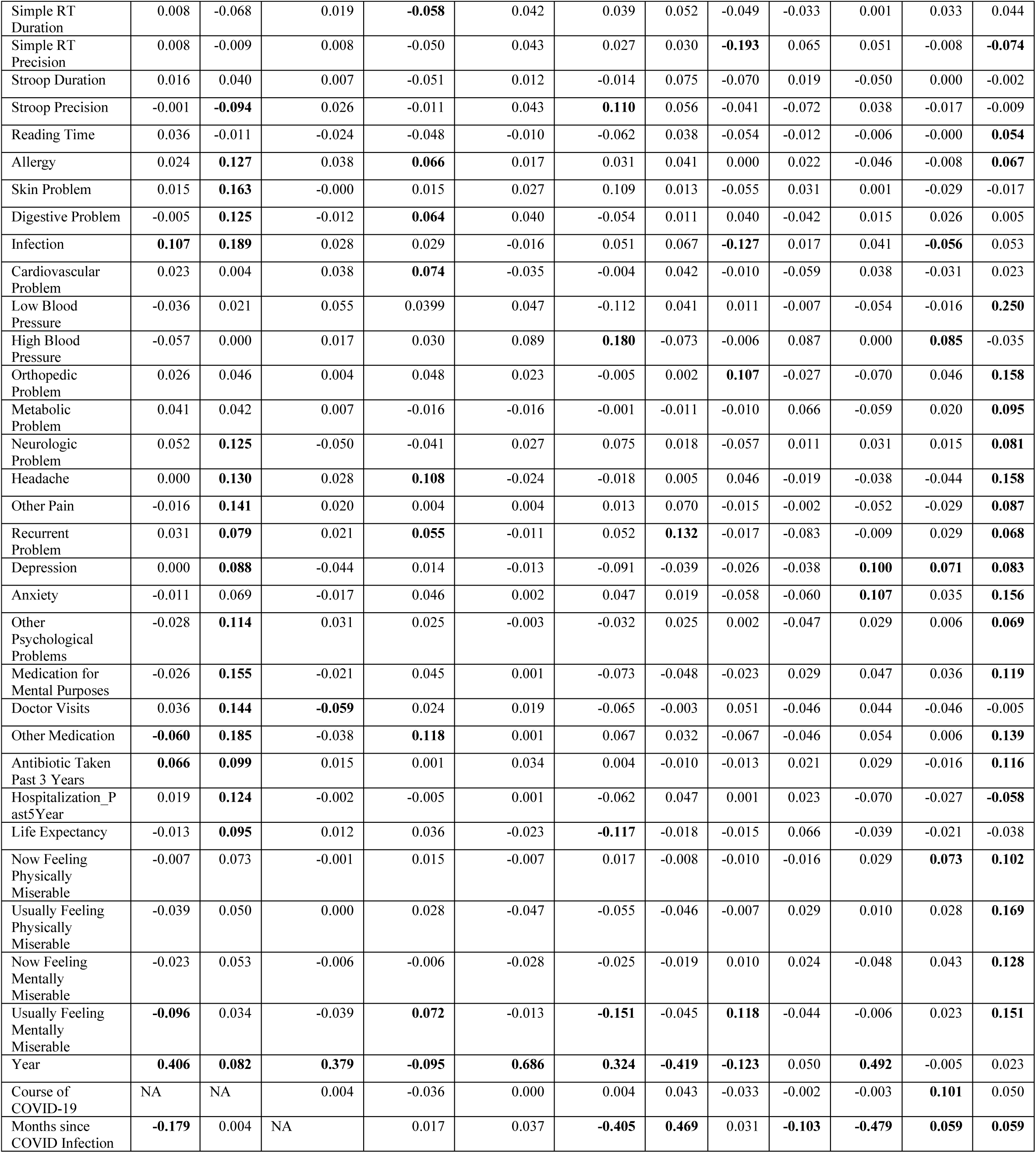
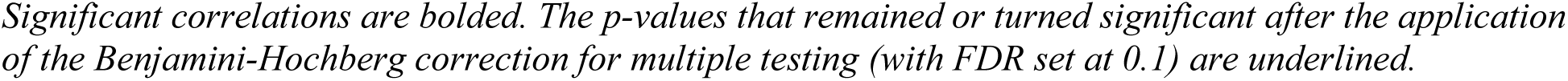
Correlations between health and performance-related variables and COVID-related variables controlled for age, sex, survey year, and time elapsed since infection-All subjects.

**Table S23.**
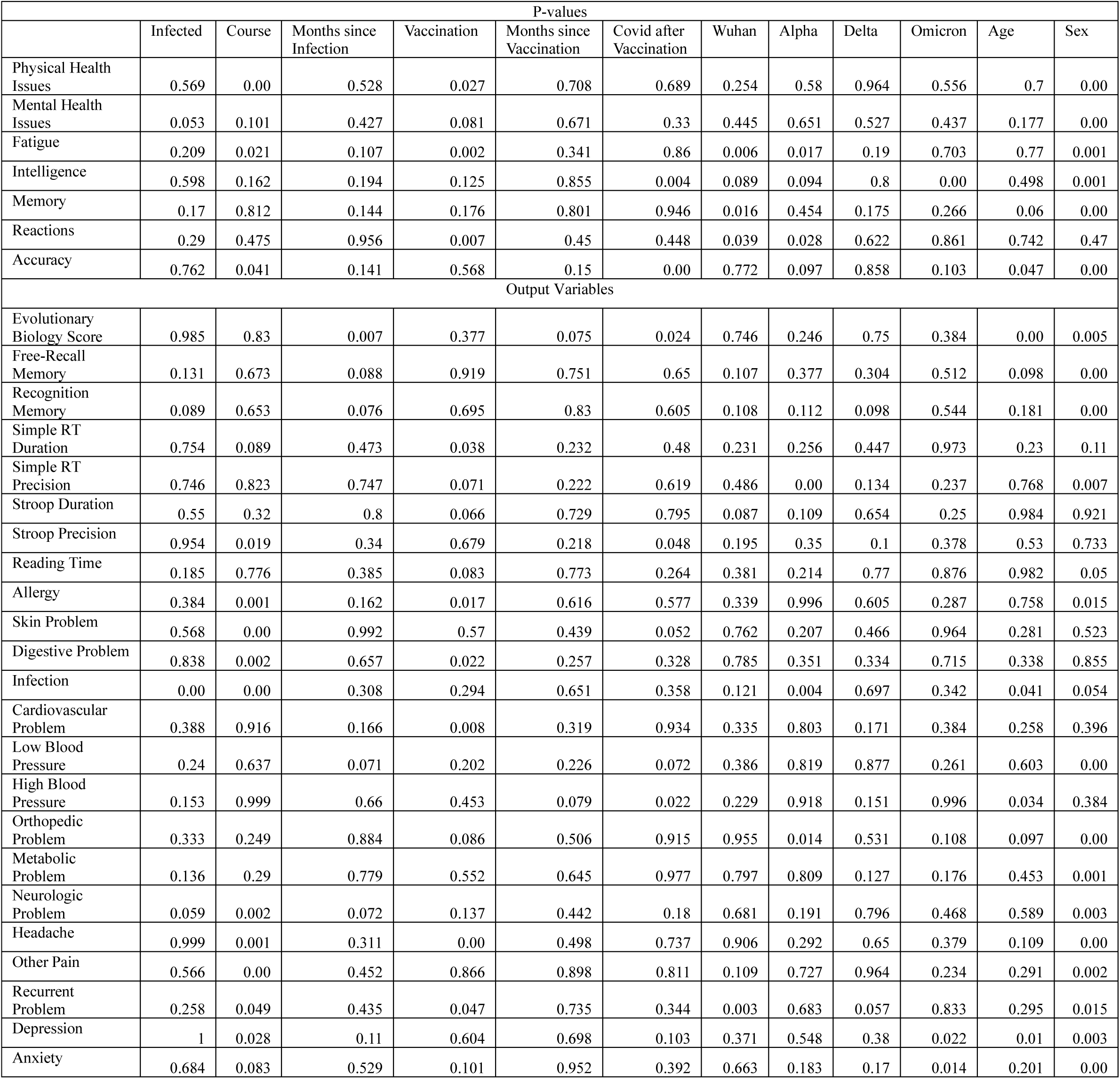
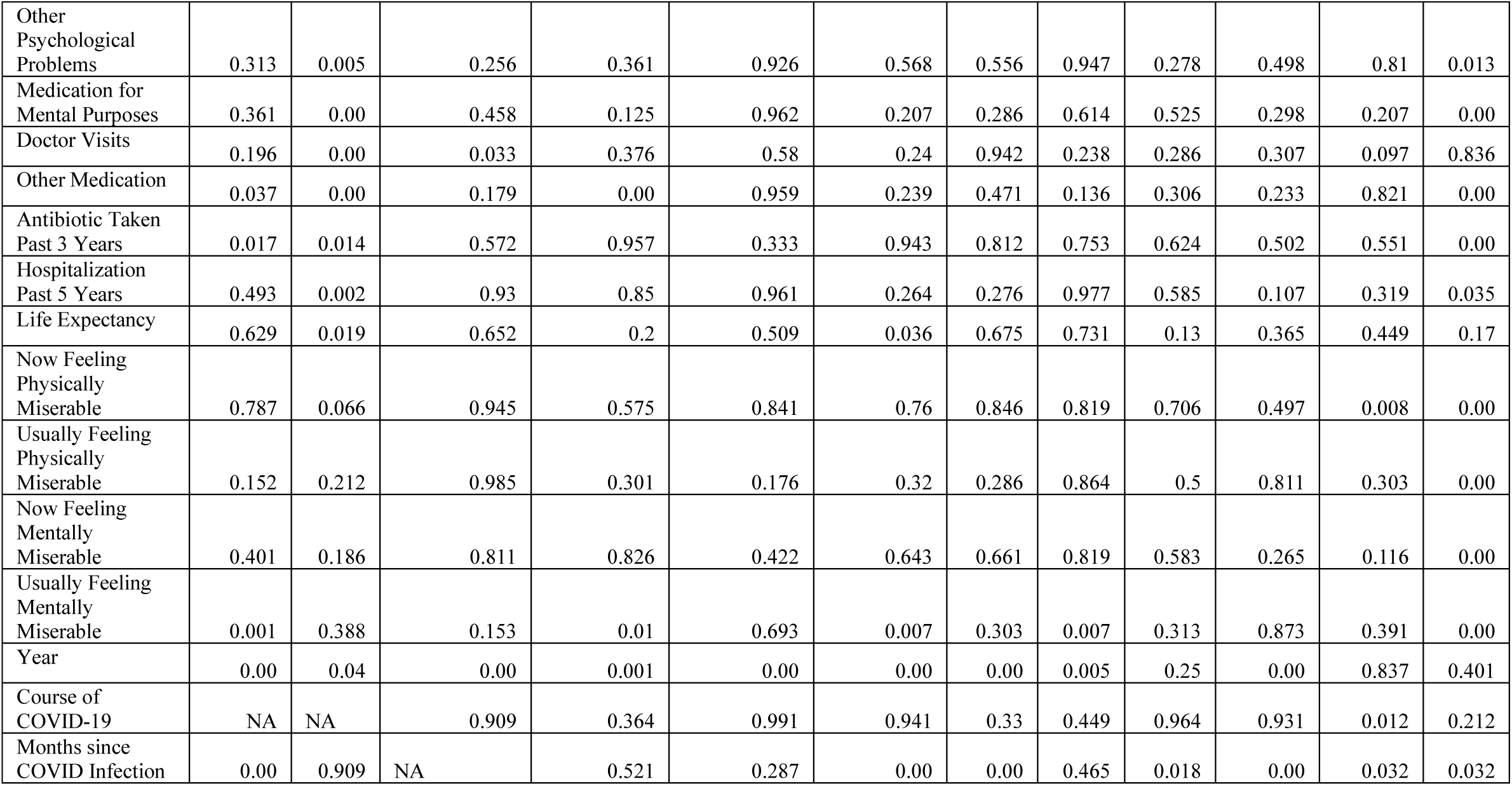
P-values of the correlations between health and performance-related variables and COVID-related variables controlled for age, sex, survey year, and time elapsed since infection-All subjects.

| P-values |  |  |  |  |  |  |  |  |  |  |  |  |
| --- | --- | --- | --- | --- | --- | --- | --- | --- | --- | --- | --- | --- |
|  | Infected | Course | Months since Infection | Vaccination | Months since Vaccination | Covid after Vaccination | Wuhan | Alpha | Delta | Omicron | Age | Sex |
| Physical Health Issues | 0.569 | 0.00 | 0.528 | 0.027 | 0.708 | 0.689 | 0.254 | 0.58 | 0.964 | 0.556 | 0.7 | 0.00 |
| Mental Health Issues | 0.053 | 0.101 | 0.427 | 0.081 | 0.671 | 0.33 | 0.445 | 0.651 | 0.527 | 0.437 | 0.177 | 0.00 |
| Fatigue | 0.209 | 0.021 | 0.107 | 0.002 | 0.341 | 0.86 | 0.006 | 0.017 | 0.19 | 0.703 | 0.77 | 0.001 |
| Intelligence | 0.598 | 0.162 | 0.194 | 0.125 | 0.855 | 0.004 | 0.089 | 0.094 | 0.8 | 0.00 | 0.498 | 0.001 |
| Memory | 0.17 | 0.812 | 0.144 | 0.176 | 0.801 | 0.946 | 0.016 | 0.454 | 0.175 | 0.266 | 0.06 | 0.00 |
| Reactions | 0.29 | 0.475 | 0.956 | 0.007 | 0.45 | 0.448 | 0.039 | 0.028 | 0.622 | 0.861 | 0.742 | 0.47 |
| Accuracy | 0.762 | 0.041 | 0.141 | 0.568 | 0.15 | 0.00 | 0.772 | 0.097 | 0.858 | 0.103 | 0.047 | 0.00 |
| Output Variables |  |  |  |  |  |  |  |  |  |  |  |  |
| Evolutionary Biology Score | 0.985 | 0.83 | 0.007 | 0.377 | 0.075 | 0.024 | 0.746 | 0.246 | 0.75 | 0.384 | 0.00 | 0.005 |
| Free-Recall Memory | 0.131 | 0.673 | 0.088 | 0.919 | 0.751 | 0.65 | 0.107 | 0.377 | 0.304 | 0.512 | 0.098 | 0.00 |
| Recognition Memory | 0.089 | 0.653 | 0.076 | 0.695 | 0.83 | 0.605 | 0.108 | 0.112 | 0.098 | 0.544 | 0.181 | 0.00 |
| Simple RT Duration | 0.754 | 0.089 | 0.473 | 0.038 | 0.232 | 0.48 | 0.231 | 0.256 | 0.447 | 0.973 | 0.23 | 0.11 |
| Simple RT Precision | 0.746 | 0.823 | 0.747 | 0.071 | 0.222 | 0.619 | 0.486 | 0.00 | 0.134 | 0.237 | 0.768 | 0.007 |
| Stroop Duration | 0.55 | 0.32 | 0.8 | 0.066 | 0.729 | 0.795 | 0.087 | 0.109 | 0.654 | 0.25 | 0.984 | 0.921 |
| Stroop Precision | 0.954 | 0.019 | 0.34 | 0.679 | 0.218 | 0.048 | 0.195 | 0.35 | 0.1 | 0.378 | 0.53 | 0.733 |
| Reading Time | 0.185 | 0.776 | 0.385 | 0.083 | 0.773 | 0.264 | 0.381 | 0.214 | 0.77 | 0.876 | 0.982 | 0.05 |
| Allergy | 0.384 | 0.001 | 0.162 | 0.017 | 0.616 | 0.577 | 0.339 | 0.996 | 0.605 | 0.287 | 0.758 | 0.015 |
| Skin Problem | 0.568 | 0.00 | 0.992 | 0.57 | 0.439 | 0.052 | 0.762 | 0.207 | 0.466 | 0.964 | 0.281 | 0.523 |
| Digestive Problem | 0.838 | 0.002 | 0.657 | 0.022 | 0.257 | 0.328 | 0.785 | 0.351 | 0.334 | 0.715 | 0.338 | 0.855 |
| Infection | 0.00 | 0.00 | 0.308 | 0.294 | 0.651 | 0.358 | 0.121 | 0.004 | 0.697 | 0.342 | 0.041 | 0.054 |
| Cardiovascular Problem | 0.388 | 0.916 | 0.166 | 0.008 | 0.319 | 0.934 | 0.335 | 0.803 | 0.171 | 0.384 | 0.258 | 0.396 |
| Low Blood Pressure | 0.24 | 0.637 | 0.071 | 0.202 | 0.226 | 0.072 | 0.386 | 0.819 | 0.877 | 0.261 | 0.603 | 0.00 |
| High Blood Pressure | 0.153 | 0.999 | 0.66 | 0.453 | 0.079 | 0.022 | 0.229 | 0.918 | 0.151 | 0.996 | 0.034 | 0.384 |
| Orthopedic Problem | 0.333 | 0.249 | 0.884 | 0.086 | 0.506 | 0.915 | 0.955 | 0.014 | 0.531 | 0.108 | 0.097 | 0.00 |
| Metabolic Problem | 0.136 | 0.29 | 0.779 | 0.552 | 0.645 | 0.977 | 0.797 | 0.809 | 0.127 | 0.176 | 0.453 | 0.001 |
| Neurologic Problem | 0.059 | 0.002 | 0.072 | 0.137 | 0.442 | 0.18 | 0.681 | 0.191 | 0.796 | 0.468 | 0.589 | 0.003 |
| Headache | 0.999 | 0.001 | 0.311 | 0.00 | 0.498 | 0.737 | 0.906 | 0.292 | 0.65 | 0.379 | 0.109 | 0.00 |
| Other Pain | 0.566 | 0.00 | 0.452 | 0.866 | 0.898 | 0.811 | 0.109 | 0.727 | 0.964 | 0.234 | 0.291 | 0.002 |
| Recurrent Problem | 0.258 | 0.049 | 0.435 | 0.047 | 0.735 | 0.344 | 0.003 | 0.683 | 0.057 | 0.833 | 0.295 | 0.015 |
| Depression | 1 | 0.028 | 0.11 | 0.604 | 0.698 | 0.103 | 0.371 | 0.548 | 0.38 | 0.022 | 0.01 | 0.003 |
| Anxiety | 0.684 | 0.083 | 0.529 | 0.101 | 0.952 | 0.392 | 0.663 | 0.183 | 0.17 | 0.014 | 0.201 | 0.00 |
| Other Psychological Problems | 0.313 | 0.005 | 0.256 | 0.361 | 0.926 | 0.568 | 0.556 | 0.947 | 0.278 | 0.498 | 0.81 | 0.013 |
| Medication for Mental Purposes | 0.361 | 0.00 | 0.458 | 0.125 | 0.962 | 0.207 | 0.286 | 0.614 | 0.525 | 0.298 | 0.207 | 0.00 |
| Doctor Visits | 0.196 | 0.00 | 0.033 | 0.376 | 0.58 | 0.24 | 0.942 | 0.238 | 0.286 | 0.307 | 0.097 | 0.836 |
| Other Medication | 0.037 | 0.00 | 0.179 | 0.00 | 0.959 | 0.239 | 0.471 | 0.136 | 0.306 | 0.233 | 0.821 | 0.00 |
| Antibiotic Taken Past 3 Years | 0.017 | 0.014 | 0.572 | 0.957 | 0.333 | 0.943 | 0.812 | 0.753 | 0.624 | 0.502 | 0.551 | 0.00 |
| Hospitalization Past 5 Years | 0.493 | 0.002 | 0.93 | 0.85 | 0.961 | 0.264 | 0.276 | 0.977 | 0.585 | 0.107 | 0.319 | 0.035 |
| Life Expectancy | 0.629 | 0.019 | 0.652 | 0.2 | 0.509 | 0.036 | 0.675 | 0.731 | 0.13 | 0.365 | 0.449 | 0.17 |
| Now Feeling Physically Miserable | 0.787 | 0.066 | 0.945 | 0.575 | 0.841 | 0.76 | 0.846 | 0.819 | 0.706 | 0.497 | 0.008 | 0.00 |
| Usually Feeling Physically Miserable | 0.152 | 0.212 | 0.985 | 0.301 | 0.176 | 0.32 | 0.286 | 0.864 | 0.5 | 0.811 | 0.303 | 0.00 |
| Now Feeling Mentally Miserable | 0.401 | 0.186 | 0.811 | 0.826 | 0.422 | 0.643 | 0.661 | 0.819 | 0.583 | 0.265 | 0.116 | 0.00 |
| Usually Feeling Mentally Miserable | 0.001 | 0.388 | 0.153 | 0.01 | 0.693 | 0.007 | 0.303 | 0.007 | 0.313 | 0.873 | 0.391 | 0.00 |
| Year | 0.00 | 0.04 | 0.00 | 0.001 | 0.00 | 0.00 | 0.00 | 0.005 | 0.25 | 0.00 | 0.837 | 0.401 |
| Course of COVID-19 | NA | NA | 0.909 | 0.364 | 0.991 | 0.941 | 0.33 | 0.449 | 0.964 | 0.931 | 0.012 | 0.212 |
| Months since COVID Infection | 0.00 | 0.909 | NA | 0.521 | 0.287 | 0.00 | 0.00 | 0.465 | 0.018 | 0.00 | 0.032 | 0.032 |

**Figure S1.**
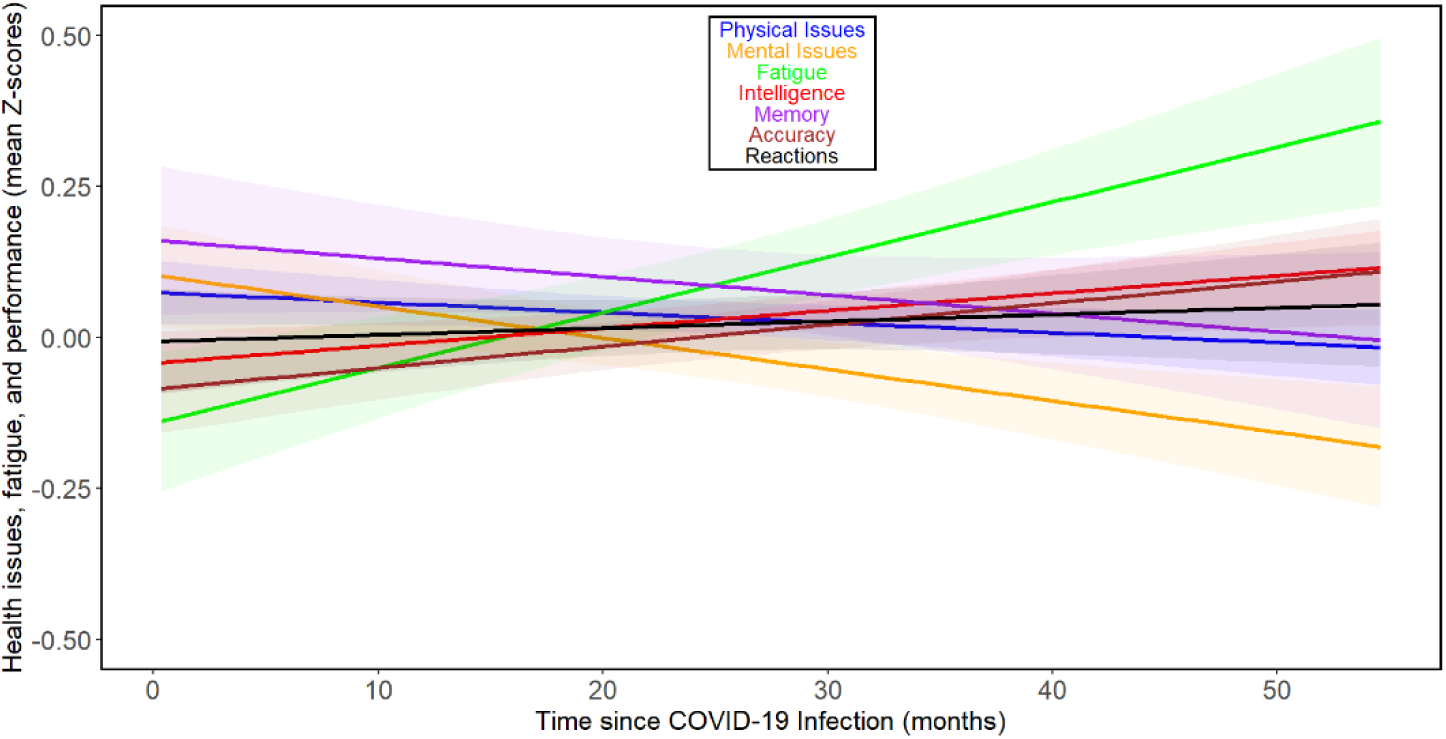
Changes in health and performance-related variables with time since COVID-related variables-Women *Based on the comparison of BIC values, all observed dependencies were best approximated by a first-degree polynomial (a straight line). The shaded areas around the lines indicate the 60% confidence intervals. It should be noted that the slopes of the lines on the graph cannot be directly compared with the (more accurate) Tau values calculated using the non-parametric partial Kendall test, which is less sensitive to the presence of outliers and additionally controlled for the influence of 3 confounding variables (sex, age, and year of survey)*.

**Figure S2.**
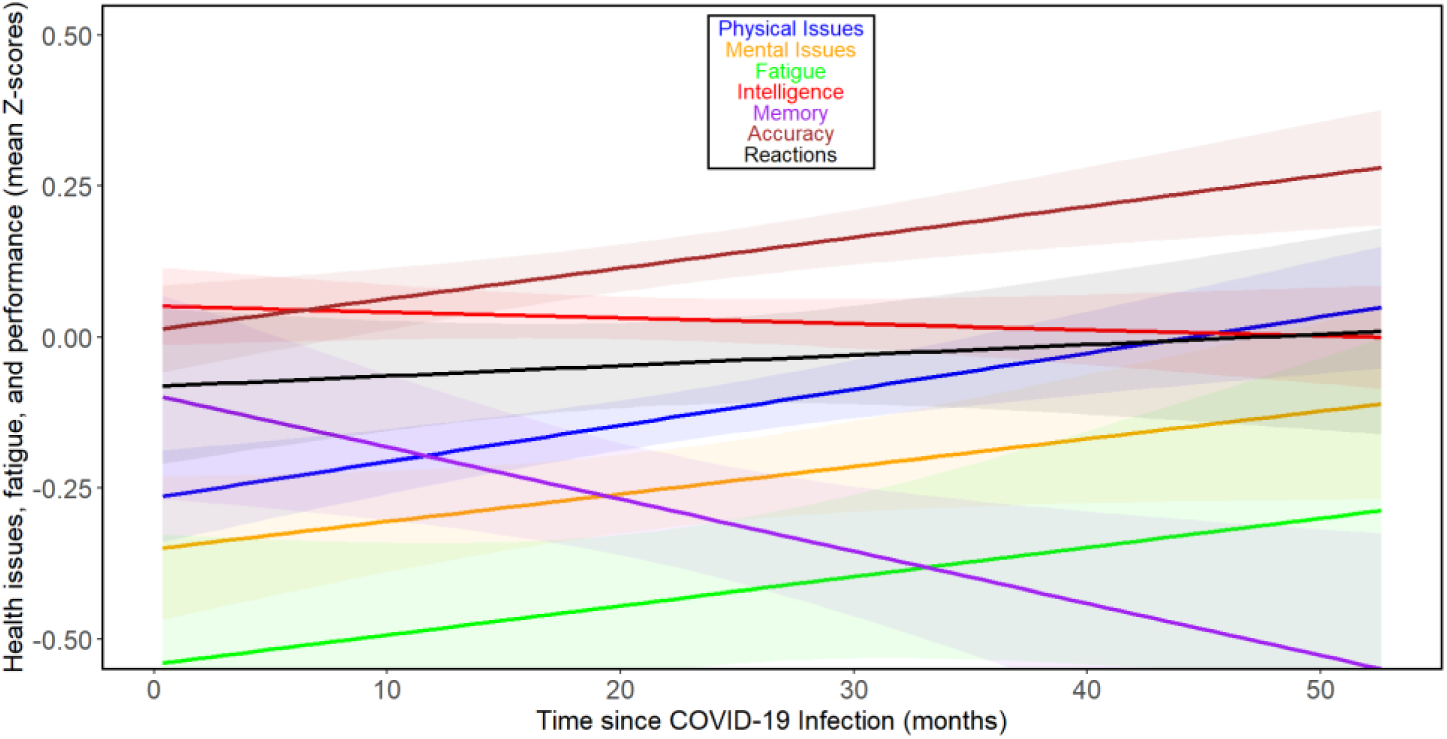
Changes in health and performance-related variables with time since COVID-related variables-men *For legends see Figure S1*.

**Figure S3.**
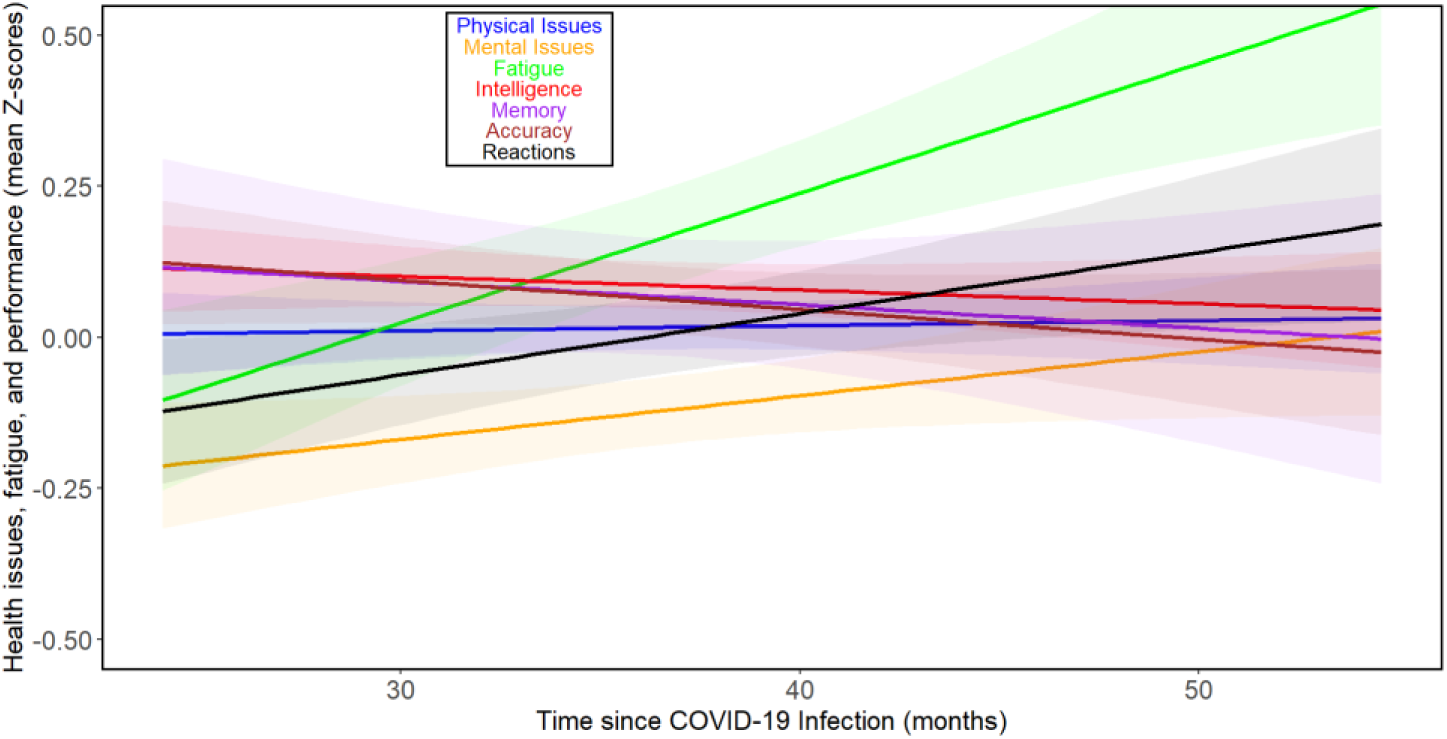
Changes in health and performance-related variables with time since COVID-19 beyond 24 months post-infection-related variables in women *For legends see Figure S1*.

**Figure S4.**
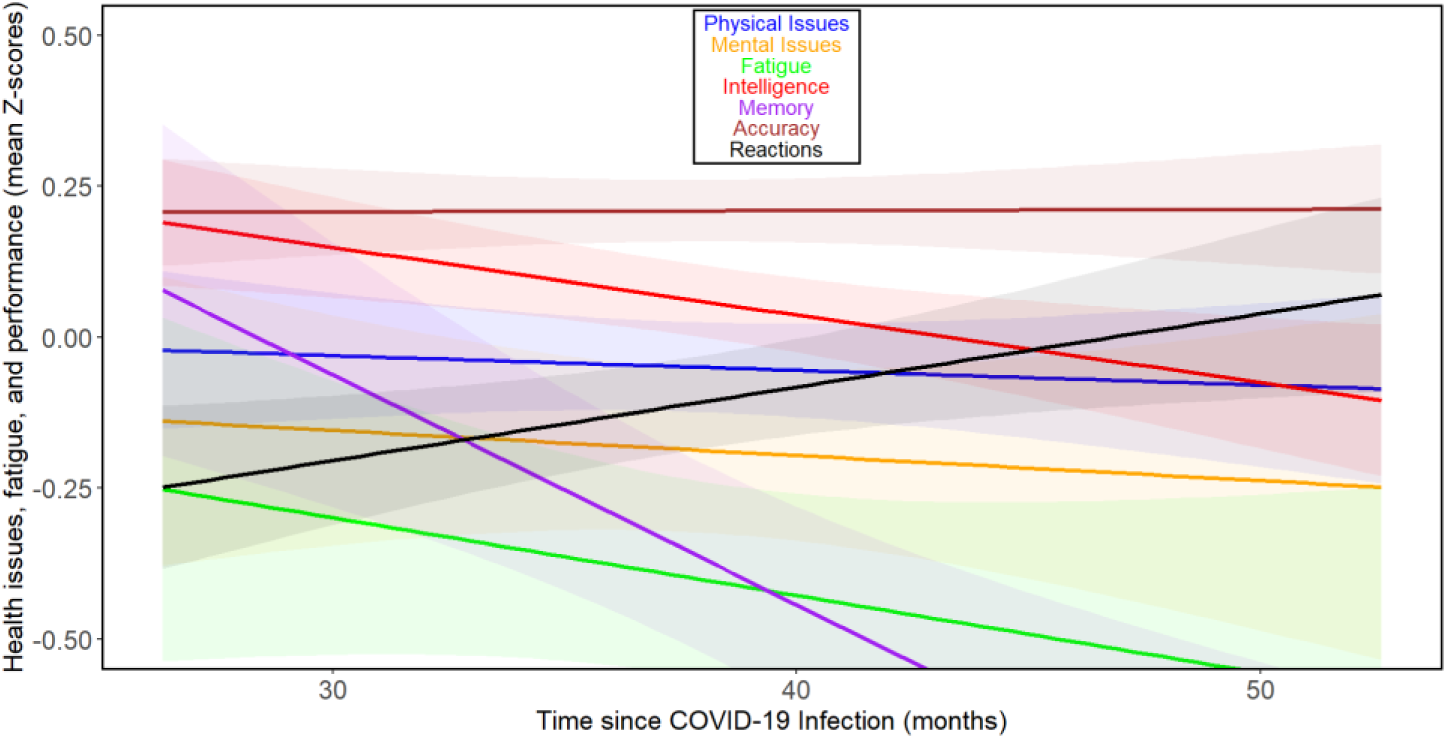
Changes in health and performance-related variables with time since COVID-19 beyond 24 months post-infection-related variables in men *For legends see Figure S1*.

**Figure S5.**
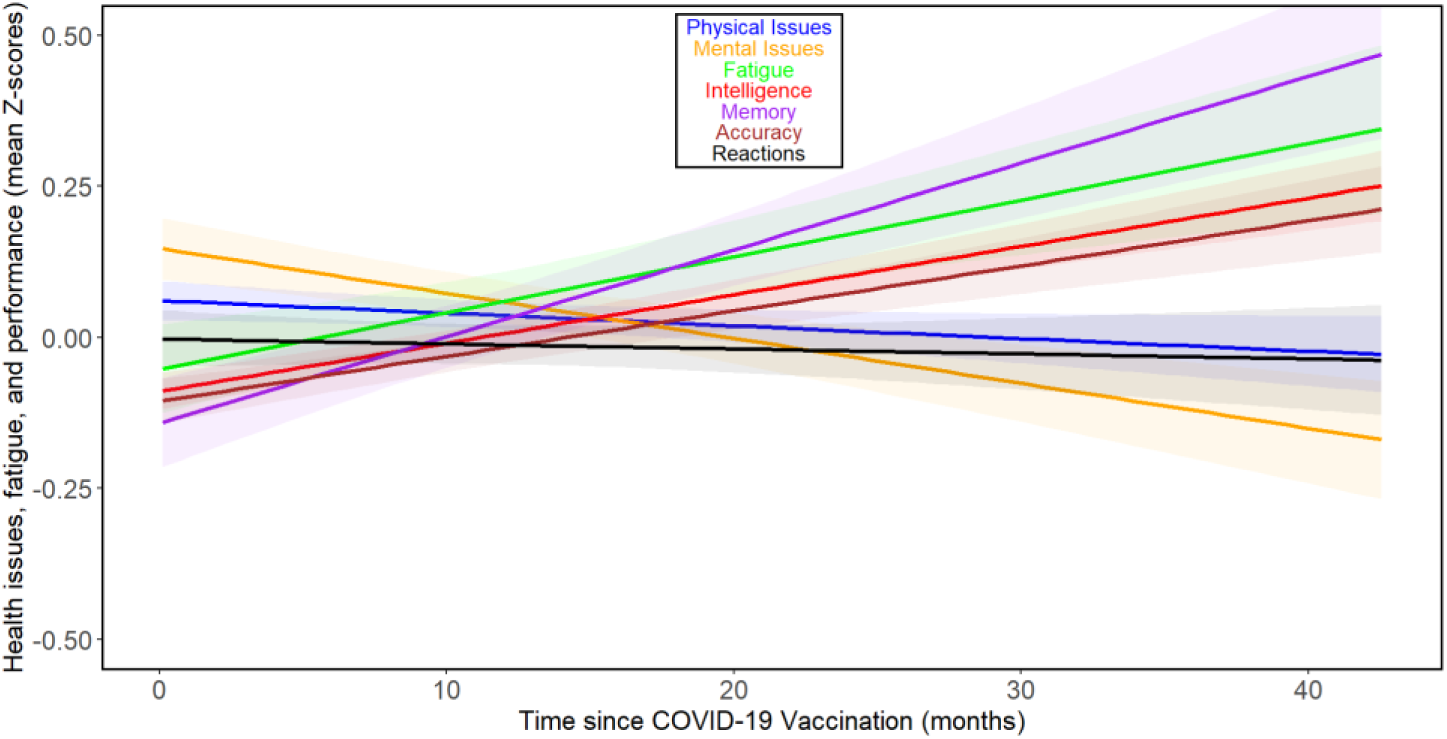
Changes in health and performance-related variables with time since COVID vaccination-related variables in women *For legends see Figure S1*.

**Figure S6.**
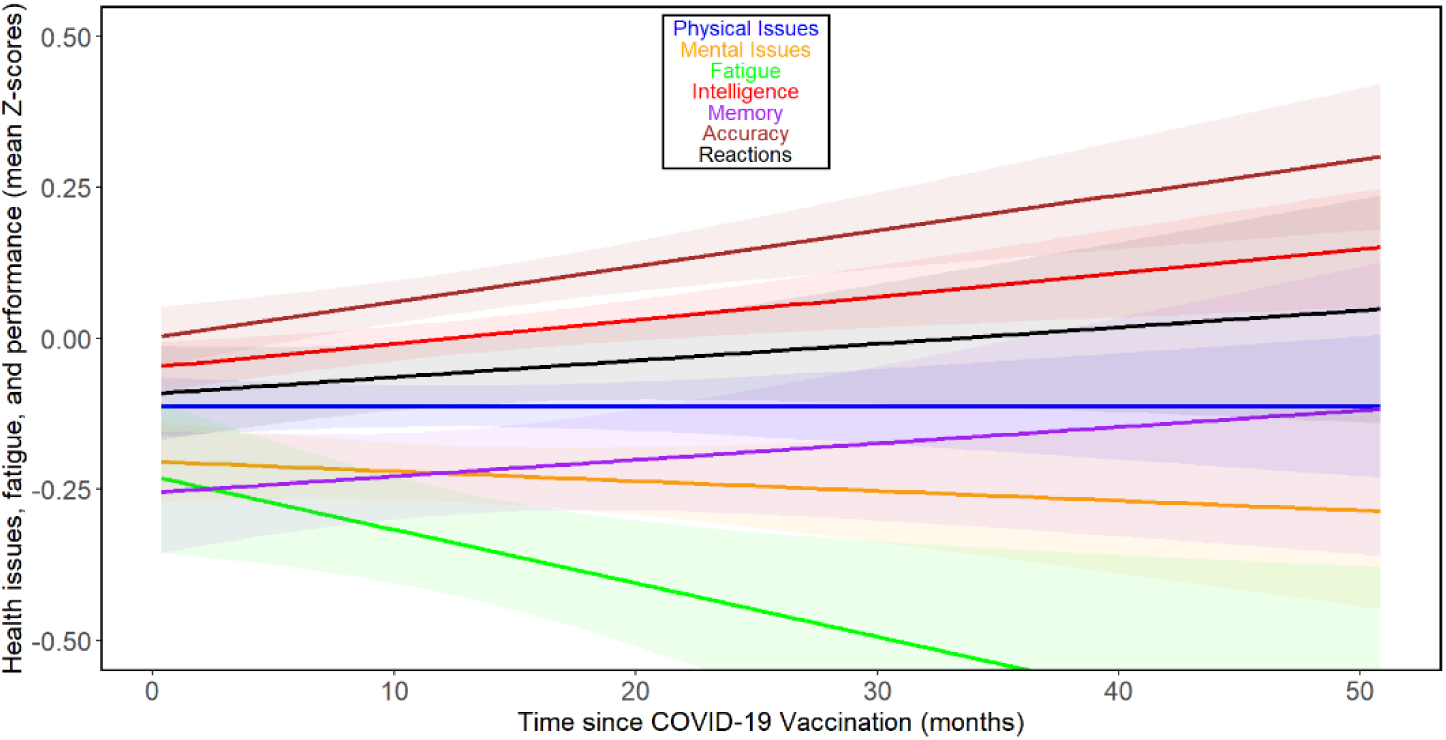
Changes in health and performance-related variables with time since COVID vaccination-related variables in men *For legends see Figure S1*.

## Notes

### Competing Interest Statement

The authors have declared no competing interest.

### Author Declarations

Institutional Review Board of the Faculty of Science at Charles University gave ethical approval for this work (No. 2021/19)

